# The role of socio-economic determinants in SARS-CoV-2 health outcomes: systematic review of population-based studies

**DOI:** 10.1101/2024.06.18.24309062

**Authors:** Jinane Ghattas, Tatjana T. Makovski, Stéphanie Monnier-Besnard, Lisa Cavillot, Monika Ambrožová, Barbora Vašinová, Rodrigo Feteira-Santos, Peter Bezzegh, Felipe Ponce Bollmann, James Cottam, Romana Haneef, Niko Speybroeck, Paulo Jorge Nogueira, Maria João Forjaz, Joël Coste, Laure Carcaillon-Bentata, Brecht Devleesschauwer

## Abstract

**Introduction:** The COVID-19 pandemic has accentuated a health-wealth gradient, reminiscent of patterns observed in previous influenza pandemics. This systematic review, employing a population-based approach, aims to delve into the etiological and prognostic roles of socio-economic factors on COVID-19 outcomes during the pandemic’s initial phase.

**Methods:** Our search spanned PubMed, Embase, WHO COVID-19 Global literature, and PsycINFO databases from January 2020 to April 7, 2021, focusing on English peer-reviewed articles. We examined the impact of socio-economic determinants on SARS-CoV-2 infection, COVID-19 related hospitalization, ICU admission, mechanical ventilation, mortality, and a range of prognostic outcomes including quality of life and mental health.

**Results:** The initial search resulted in 9,701 records after removal of duplicates. Out of hundred articles that met our review criteria, 67 discussed the etiological role of socio-economic factors, 25 addressed the prognostic role, and 8 covered both. Fifty-nine percent of the studies were from the United States of America and the United Kingdom, highlighting an increased risk of infection and severity among their Black, Asian, and Hispanic populations. Lower-income groups, crowded households, and, higher socio-economic deprivation were associated with higher COVID-19 incidence and severity. Results regarding educational status varied across different waves.

**Conclusion:** Populations groups with disadvantaged socio-economic positions and certain ethnic and racial backgrounds face a higher risk of SARS-CoV-2 infection and poorer COVID-19 outcomes. Our findings underscore the need for incorporating social determinants into routine health surveillance and monitoring, suggesting an avenue for targeted interventions.

**What is already known on this topic:** The association between socio-economic factors and outcomes, observed during the COVID-19 pandemic and other influenza epidemics, indicates a significant health-wealth gradient. Higher mortality and worse health outcomes are observed in the most disadvantaged groups of the population

**What this study adds:** This study lays out a comprehensive review of population-based studies, representative of the general population, on the etiological and prognostic role of socio-economic characteristics in COVID-19 outcomes covering studies across 22 countries. Our results show that population-based studies from countries in Asia, Africa, Latin America, and continental Europe were limited in the early phase of the pandemic and highlight the importance of socio-economic characteristics, alongside comorbidities, for predicting SARS-CoV-2 infection and COVID-19 severity.

**How this study might affect research, practice or policy:** This evidence shows a gap in the literature in countries other than the USA and the UK and on the role of socio-economic characteristics in long-term COVID-19 outcomes. Social inequalities need to be at the heart of infectious disease monitoring and surveillance to decrease the gap in adverse outcomes between patients of different socio-economic profiles.

## 1. BACKGROUND

The association between socio-economic (SE) factors and influenza outcomes, evidenced across various influenza epidemics and pandemics, highlights a pronounced health-wealth gradient (1). Notably, during the 2009-2014 influenza pandemic, England witnessed higher mortality rates among its most deprived populations (2). Similarly, the COVID-19 pandemic has revealed analogous patterns, with increased mortality observed among individuals of lower socio-economic status (SES), including immigrants and ethnic minorities (3–5). Such disparities are further exacerbated by unhealthy lifestyle behaviours predominantly found in disadvantaged groups, significantly raising the odds of COVID-19 mortality (6). This underscores the critical need for incorporating health equity into the early stages of pandemic surveillance and preparedness (7, 8). Despite the growing literature on the role of socio-economic (SE) factors in COVID-19 outcomes, there remains a gap in summarizing this interplay from a population-based perspective during the pandemic’s early phase before the introduction of the vaccine.

Using a population-based approach, this systematic review aims to fill this gap by exploring two primary objectives:

1. Assessing the etiological role of socio-economic characteristics in initially SARS-CoV-2 negative cohorts of general population.
2. Investigating the prognostic role of socio-economic characteristics in SARS-CoV-2 positive general population to evaluate case severity and disease progression.

This study is embedded within a larger project supported by the European Union’s (EU) Horizon 2020 research and innovation program (grant agreement No: 101018317), alongside other factors such as multimorbidity and frailty (9–11). In this study, we will only focus on the role of socio-economic factors in COVID-19 severity.

## 2. METHODS

The study was structured following the guidelines of the Preferred Reporting Items for Systematic Review and Meta-Analysis Protocols (12). The protocol was officially documented in the Prospero registry for systematic review protocols under the identification number CRD42021249444 and subsequently published in a peer-reviewed journal where further details on the methodology can be found (9).

### 2.1 Study approach

We used a population-based approach defining populations within specific geographic regions during distinct time frames (13). It aims for findings generalisable to the entire population that is being investigated in the study hypothesis, rather than just to the individuals who were included in the study (14).

Based on the Population, Exposure, Comparator, and Outcomes framework (15), we provide below a comprehensive description of the study objectives:

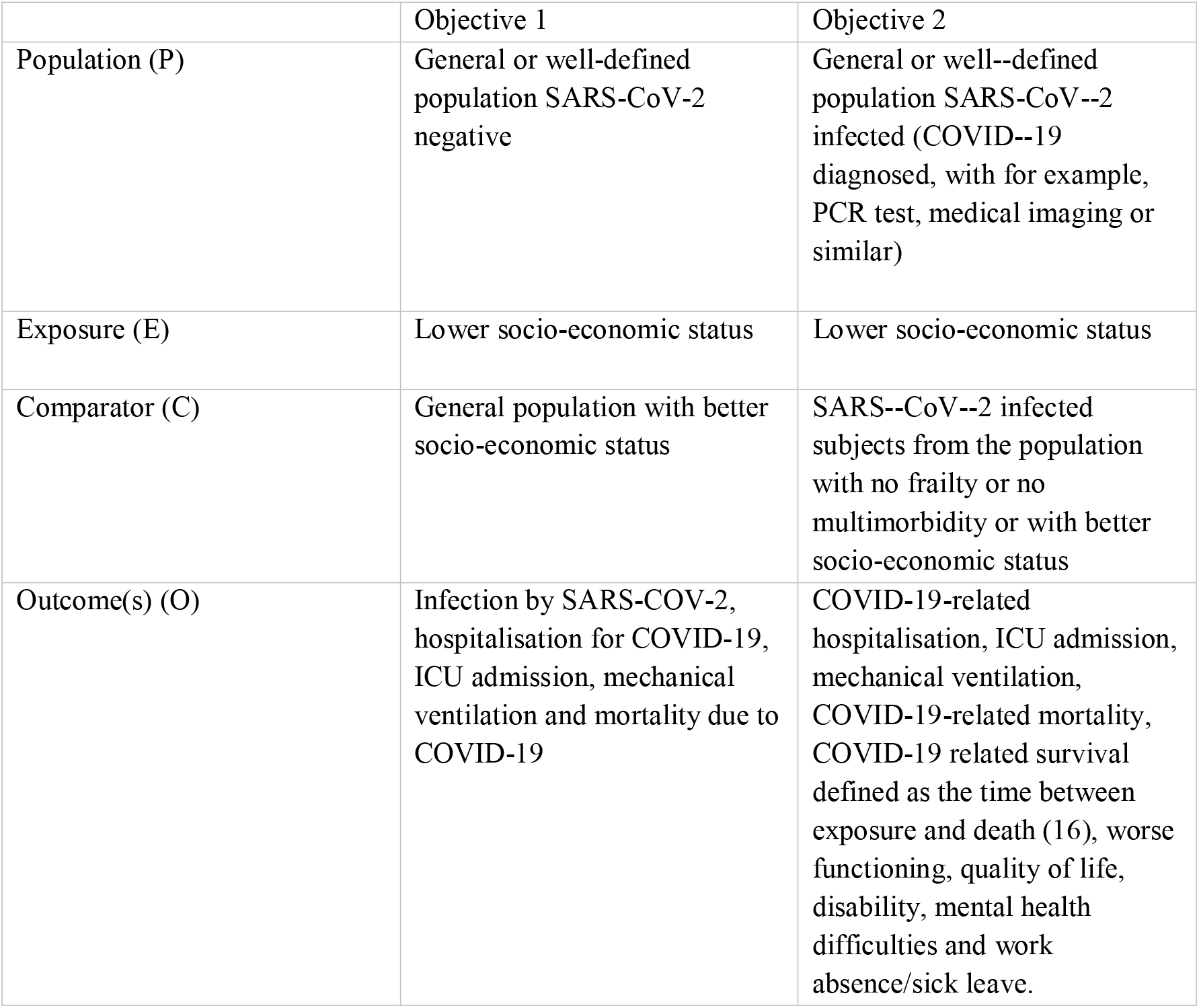

|  | Objective 1 | Objective 2 |
| --- | --- | --- |
| Population (P) | General or well-defined population SARS-CoV-2 negative | General or well-defined population SARS-CoV-2 infected (COVID-19 diagnosed, with for example, PCR test, medical imaging or similar) |
| Exposure (E) | Lower socio-economic status | Lower socio-economic status |
| Comparator (C) | General population with better socio-economic status | SARS-CoV-2 infected subjects from the population with no frailty or no multimorbidity or with better socio-economic status |
| Outcome(s) (O) | Infection by SARS-COV-2, hospitalisation for COVID-19, ICU admission, mechanical ventilation and mortality due to COVID-19 | COVID-19-related hospitalisation, ICU admission, mechanical ventilation, COVID-19-related mortality, COVID-19 related survival defined as the time between exposure and death (16), worse functioning, quality of life, disability, mental health difficulties and work absence/sick leave. |

Observational studies of all types were included, including cohort studies, cross-sectional studies, case-control studies, and ecological studies considering the importance of including contextual factors. Quantitative papers published in English were considered.

The following standard socio-economic indicators were included: income, education, occupation, employment, housing, urban/rural setting, household size, race, ethnicity, nationality and marital status. These indicators were examined at the individual or area level (ecological studies). Acknowledging that socio-economic differences may be observed across all ages, all age groups were considered.

### 2.2 Search strategy and study selection

Databases search included PubMed, Embase, WHO COVID-19 Global literature, and PsycINFO databases, from January 2020 to April 7, 2021. A ’backwards’ snowball search enhanced our literature review. Independent reviewer pairs screened titles, abstracts, and full texts, using Rayyan for data management (17). Pairs of reviewers independently completed the title/abstract screening and full text reading. Any disagreements were resolved via consensus or a third reviewer. Exclusion criteria were applied systematically, focusing on language, originality of research, relevance, and study design.

List 1 details the reasons for excluding studies at the initial screening stage, including language barriers, lack of original research, irrelevance to the review’s scope, non-population-based studies, and focus on specific subpopulations. The enumeration of excluded studies underscores the stringent criteria applied to ensure the review’s focus and quality. Further refinement during the full-text assessment phase led to exclusions based on more specific criteria, such as unclear COVID-19 diagnosis, absence of socio-economic data, outcomes outside the study objectives, and duplication of populations across studies (List 2). This process ensured that only studies directly relevant and methodologically sound were included in the final review.

**List 1:** Number of rejected studies and reasons for systematic review exclusion (title/abstract or records screening phase)

| Criteria for exclusion | # excluded studies |
| --- | --- |
| 1 = Article not in English | 7 |
| 2 = Not an original research (e.g. editorial, protocol, etc.) or no original results | 597 |
| 3 = Unrelated topic (e.g. outside of the review scope) | 8 078 |
| 4 = Not a population-based study | 534 |
| 5 = Subpopulation (medical staff, students, pregnant women, etc.) | 74 |

**List 2:** Number of rejected studies and reasons for systematic review exclusion (full-text reading or reports screening phase)

| Reasons for exclusion | # excluded studies |
| --- | --- |
| 1 = Not a population-based study | 84 |
| 2 = SARS-CoV-2 infection diagnosis not clear* | Not assigned |
| 3 = Study does not contain information on socio-economic characteristics | 15 |
| 4 = Outcome not within the scope of our objectives | 27 |
| 5 = Outcome measurement tool not clear* | 1 |
| 6 = Risk factor measurement tool not clear* | Not assigned |
| 7 = Subpopulation (e.g. pregnant women, health care workers, students etc.) | 2 |
| 8 = Not an original research (e.g. editorial, protocol, review, conference abstract, grey literature, etc.), no original results or not peer-reviewed | 31 |
| 9 = The same or largely the same population already considered in another study for the same outcome | 2 |
| 10 = Clinical trial or intervention study | Not assigned |
| 11 = Qualitative study | Not assigned |
| 12 = Descriptive study, absence of a comparator group and/or no measure of the association of interest | 30 |
| 13 = Other (explain)** | 6 |

\*even after contacting authors
\*\*e.g. full text not found, research question outside of the review scope, case-series, etc.

### 2.3 Data synthesis

A piloted data extraction template guided the independent extraction process, incorporating study details, population characteristics, COVID-19 diagnostics, and outcome measurements. Any disagreements were resolved via consensus or a third reviewer. Data extraction was conducted using Microsoft Excel.

### 2.4 Quality assessment

The Newcastle-Ottawa scale was used to assess the risk of bias in cohort and case-control studies (18).

An adjusted version of the Newcastle-Ottawa scale was used to assess the quality of cross-sectional studies. A higher score indicates higher study quality with a maximum of 9 points for cohort and case-control studies and 10 points for cross-sectional studies (Supplemental file 4).

### 2.5 Patient and public involvement

Patients and public were not directly involved in this systematic review.

## 3. RESULTS

Our database search identified 19,108 records. After deduplication, we screened the titles/abstracts of 9,701 records. 411 records were included in the full-text screening phase of which 276 records focused on the etiological and prognostic role of socio-economic factors in SARS-CoV-2 infection and COVID-19 severity. In this phase, 198 records were excluded and 78 studies were finally included in the narrative review. The backward “snowballing’ resulted in additional 22 articles that were extracted (Figure 1). In sum, a total of 100 articles were reviewed.

**Fig. 1.**
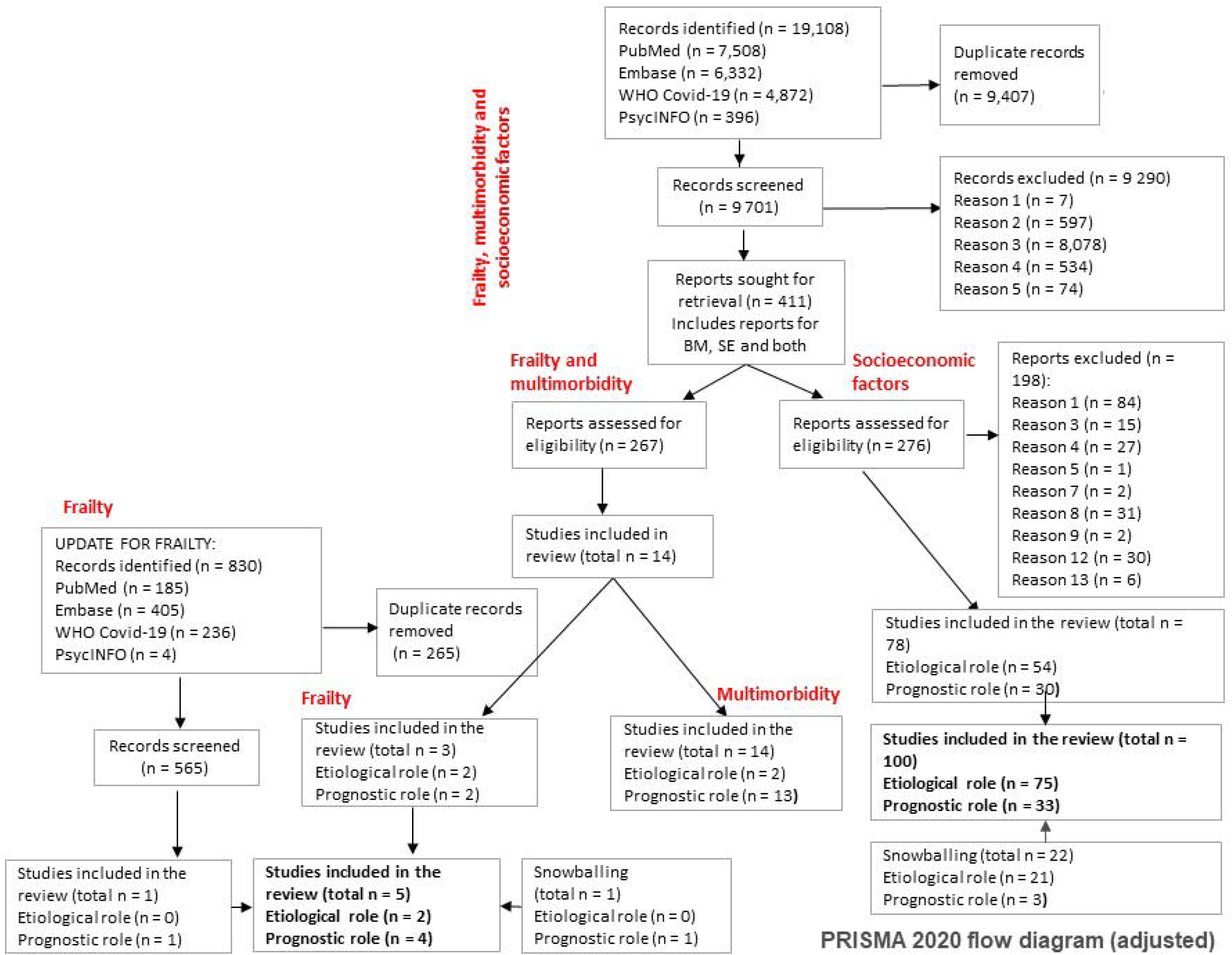
PRISMA 2020 flow diagram for new systematic reviews (adjusted)

### Study characteristics

Out of hundred articles included in this study, 67 studies reported the etiological role of socio-economic determinants in short-term COVID-19 outcomes (5, 19–84), 25 studies focused on the prognostic role (85–109) and eight articles covered both (110–117). Fifty-four studies were based on individual-level data while 46 studies included aggregate-level data. Cross-sectional study design was the most frequent design (52 studies), followed by cohort studies (45 studies) and case-control studies (3 studies).

Most of the studies were published in high-income countries (USA, n = 39; UK, n = 20) (Figure 2). 64 studies were set in the general population, 21 in hospital settings, seven in nursing homes, three in primary care settings, two in emergency or ICU setting, and three included outcomes from the general population and hospitalised patients.

**Fig. 2.**
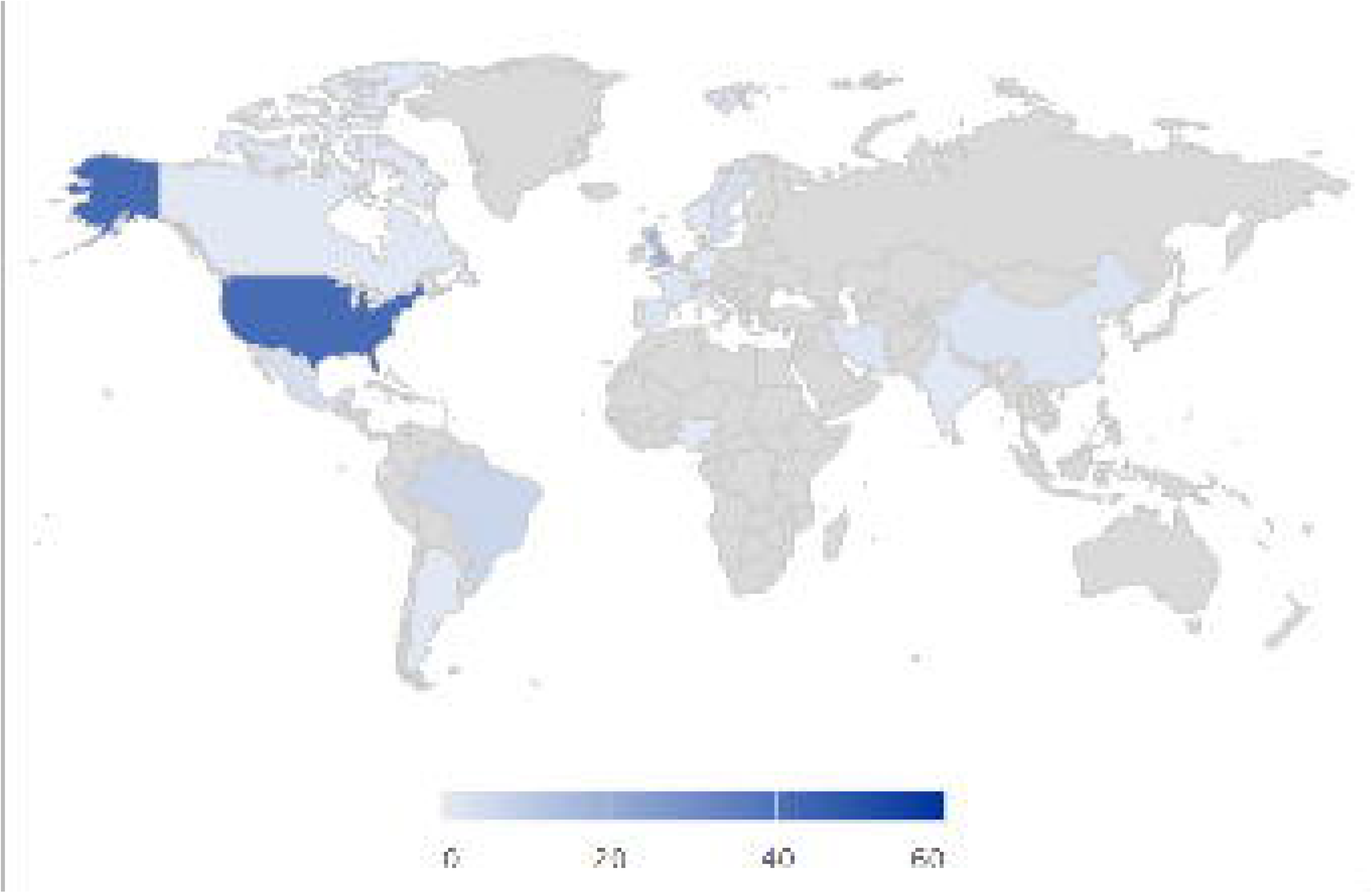
Geographical distribution of population-based studies

During the early phase of the pandemic, studies reported only short-term outcomes such as SARS-CoV-2 infection, hospitalisation, ICU admission, mechanical ventilation, mortality, survival and severe COVID-19 outcome. No studies reported long-term outcomes such as functioning, quality of life, disability, mental health difficulties or work absence/sick leave. COVID-19 diagnosis was usually confirmed by an RT-PCR test or International Classification of Diseases (118). Self-reporting, administrative data, and population-based surveys were the main sources from which the information on socio-economic characteristics was obtained. The sample size in each study varied significantly and is reported in tables 1 and 3 of the supplemental file 2.

Regarding quality assessment, individual-level cohort studies scored between 6 and 9 (out of 10) and studies conducted at the ecological level received a score of either 6 or 7. Quality assessment for individual-level cross-sectional studies ranged between 6 and 8 while ecological cross-sectional studies varied from 4 to 10. Two case-control studies received a score of seven, while one study received a score of six (Supplemental file 3).

### Etiological role of socio-economic characteristics

#### Studies reporting on infections

Fifty-five studies reported on the etiological role of socio-economic determinants in SARS-2-CoV infections (19, 21, 22, 28, 29, 33–36, 38, 39, 41–46, 48–50, 52, 53, 56–65, 67, 69–74, 76, 78–84, 110–112, 114–117, 119). Seventeen studies were performed at an individual-level (19, 21, 22, 29, 33–36, 38, 39, 41, 42, 111, 112, 115, 117, 119) while ecological studies accounted for 37 of the total (43–46, 48–50, 52, 53, 56–65, 67, 69–74, 76, 78–84, 110, 114, 116). One study reported both individual and aggregated data (28). In individual-level studies, the sample size ranged between 378 and 17,636,366 individuals.

The studies with individual-level socio-economic determinants used one or several of the following indicators: race, ethnicity, country of birth, marital status, education, income, employment, housing conditions and tenue, household characteristics, urban or rural place of residence, deprivation score. In ecological studies, the socio-economic determinants included scores reporting on ethnicity, race, education, income, employment, household size and crowding, population density, housing characteristics such as tenure, insurance as well indexes of social vulnerability, development, deprivation, and inequality (Figure 3). Seventeen studies provided unadjusted and adjusted estimates while 36 provided only adjusted measures of associations and 2 reported unadjusted results.

**Fig. 3.**
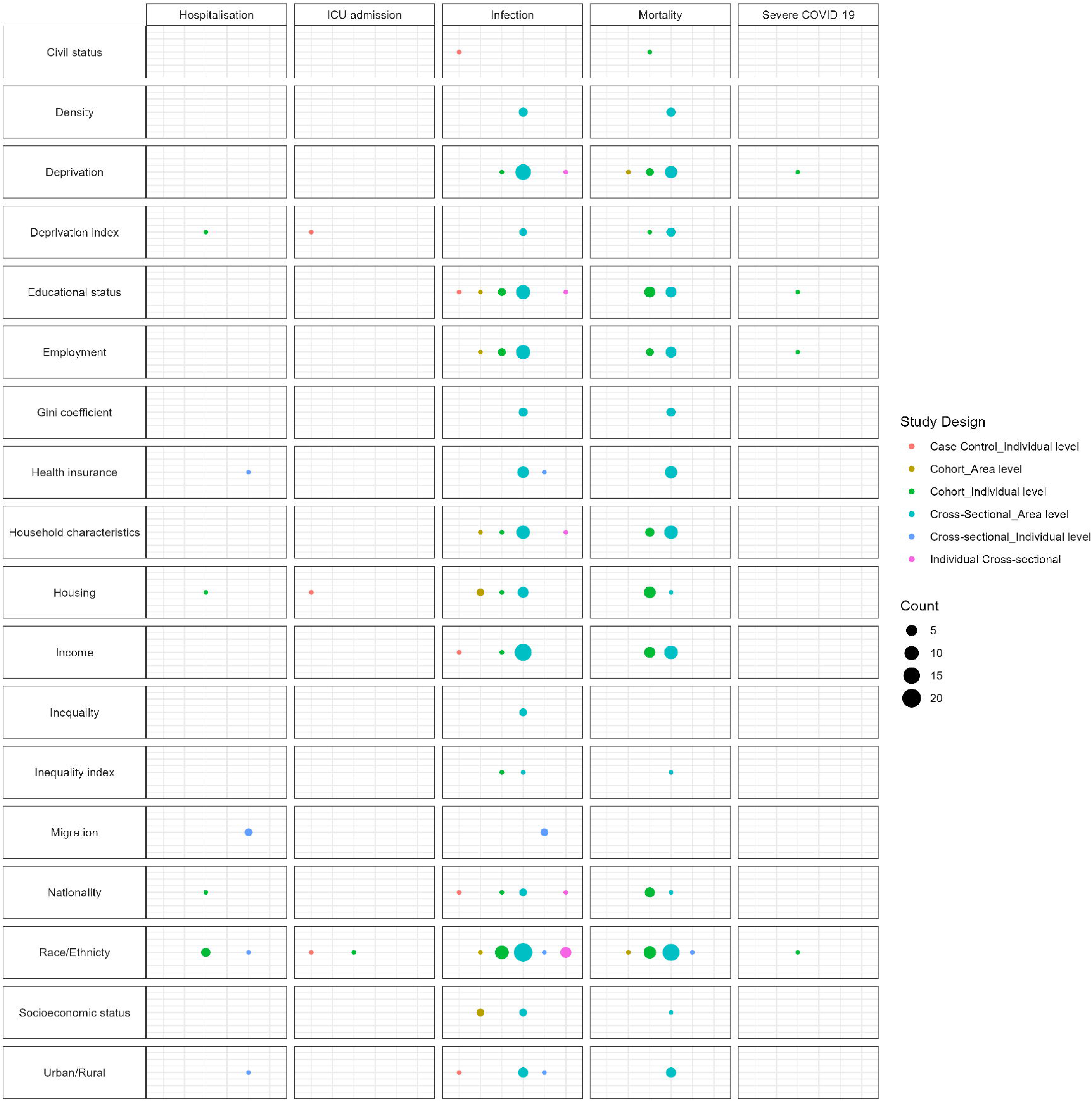
Bubble diagram summarising the etiological role of socio-economic determinants and COVID-19 outcomes per study types

**Fig. 4.**
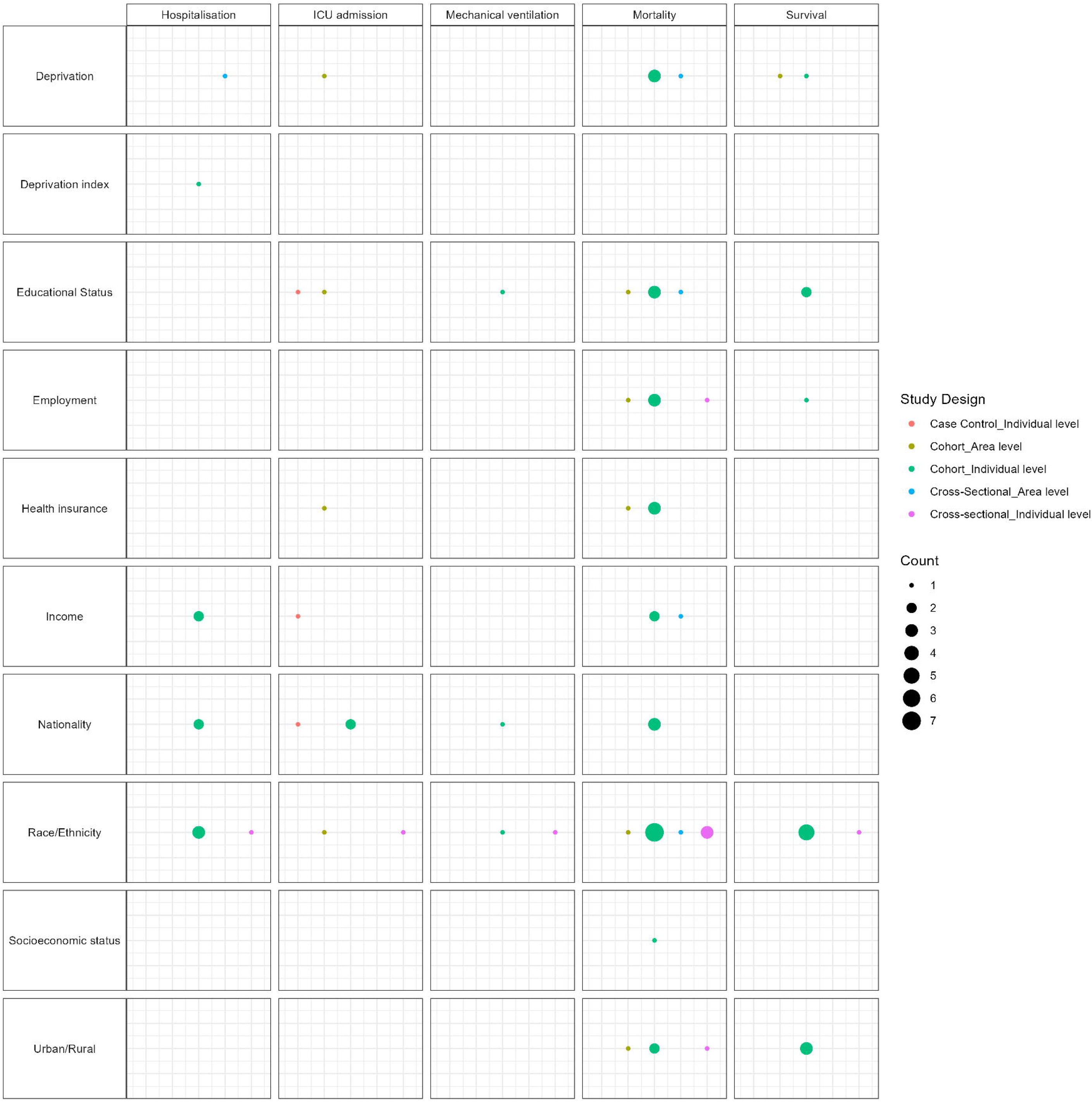
Bubble diagram summarising the prognostic role of socio-economic determinants and COVID-19 outcomes per study types

According to the overall findings, Black, Hispanic, and Asian people in the UK and the USA, as well as migrants in Italy and Sweden were more likely to contract SARS-CoV-2 infection. In addition, socio-economic deprivation scores, social vulnerability, interstate migration, living in a rental property, poor housing conditions, larger household size and a lower socio-economic status (SES) were all positively associated with infection risk. SARS-CoV-2 infections were more pronounced in areas of higher inequalities.

The results on educational status were heterogeneous: some studies reported a higher SARS-CoV-2 infection in populations with lower educational status (22, 28, 36, 38, 59, 63, 67) while other studies showed that higher educational level was associated with higher odds of COVID-19 diagnosis (48, 82, 111). The results from Yang *et al,* documented that patterns of infections varied across the different waves (57) . For instance, tertiary education or above was associated with higher SARS-CoV-2 infection risk during the first wave but it had a protective effect during the second and third wave (57) (Table 1 and 2, Supplemental file 2).

#### Studies reporting on hospitalisations

Five individual-level studies reported on the etiological role of socio-economic characteristics in COVID-19-related hospitalisations (21, 23, 26, 42, 115). Four were cohort studies and one was cross-sectional. Two studies were performed in the UK (23, 42), one in Spain (21), one in Mexico (115) and one in the USA (26). Study samples included ranged from 17,763 to 25,333,329 individuals.

One out of five studies stratified the analysis by sex and adjusted for age, BMI, comorbidities and treatments (23) and another study reported COVID-19 outcomes during two different waves (42). In one of the studies, information on socio-economic determinants was self-reported (42), while in the other four studies, information was extracted from administrative records (21, 23, 26, 115).

Overall, patients identified as White had a lower risk of hospitalisations compared to other ethnic groups independently of sex in the UK and the USA (23, 26, 42). Being Asian, Black, Hispanic, North American native, or from other mixed ethnic groups increased the risk of hospitalisation (26, 42). The study by Mathur *et al.* reported results from a cohort of adults registered with primary care practices in England over two waves (42). The second waves showed an attenuated risk of hospitalisation for Black(42). Patients from Sub Saharan Africa, Latin America and Caribbean were also at higher risk of hospitalisation compared to Spanish patients (21). Indigenous ethnicity in Mexico was not significantly associated with an increased risk of COVID-19 hospitalisation (115).

The risk of being hospitalised for COVID-19 was higher among individuals residing in care homes, experiencing deprivation (23) and with interstate migration status (115). However, the impact of urbanisation on hospitalisation risk was minimal yet statistically significant (115) (Table 1 and 2, Supplemental file 2).

#### Studies reporting on ICU admission

Only one study conducted in the UK looked at the etiological role of socio-economic factors in ICU admission. It included a cohort of adults enrolled in primary care practices in England and spans two periods, from February 2020 to December 2020 (42).

During both waves, the adjusted analysis showed that being South Asian, Black, mixed ethnic group and others were found to have a greater risk of ICU admission when compared to White patients (42) (Table 1 and 2, Supplemental file 2).

#### Studies reporting on mortality

In the thirty-six studies reporting on the etiological role of socio-economic determinants in mortality, 12 studies used individual-level measures(5, 20, 23–27, 30, 32, 37, 42, 113, 120) and 24 used area-level measures(43–45, 48, 50, 51, 53–55, 59, 60, 63, 64, 66–69, 74–77, 79, 80, 84). Individual-level studies included a sample size ranging between 274,712 and 306,800,000 individuals. Studies used one or several of the following indicators: ethnicity, country of birth, country of residence, educational level, income, employment status, civil status, house tenureship, and household size. Deprivation was referred to using the index of multiple deprivation quintiles (IMD) and the Townsend deprivation index. Three studies stratified per sex (23, 24, 37) and one study reported mortality outcomes over several pandemic waves (42). The following socio-economic indicators were used in ecological studies: race, ethnicity, education, income, crowding, density, household size, urban population, employment, insurance, indexes of social vulnerability, deprivation, poverty and Gini coefficient. One study stratified the analysis by sex (54).

Studies from the USA and UK reported worse outcomes in Black, Hispanic and Indian COVID-19 patients (5, 23, 24, 26, 27, 30, 44, 50, 51, 59, 63, 66–69, 77, 113, 120). The study conducted by Mathur et al. revealed different outcomes over multiple waves (42). Specifically, during the first wave in England, Black, Asian, and mixed-race patients were found to be at a greater risk of COVID-19 mortality compared to white patients. However, the results were not statistically significant for Black and mixed-race patients during the second wave. In contrast, the risk remained higher for Asian patients (42). Three studies reported demographic determinants based on country of birth. Being a migrant from low and middle income countries increased the risk of COVID-19 death compared to people born in Sweden (20, 25, 32).

Increased risk of death was also observed in people living in long-term care homes (20, 23, 32), crowded households (20), multigenerational households (37), and in patients with lower educational attainment and lower income (20, 25, 27, 32, 59, 63, 113). Another study came to similar findings, concluding that higher education was a protective factor against COVID-19 mortality (48).

A positive association was observed between deprivation and increased risk of COVID-19 mortality (5, 23, 113). The risk of COVID-19 death was consistently higher for the most disadvantaged compared to the least disadvantaged (48, 50, 54, 55, 67, 68, 75, 84). Additionally, county crowding and increased social vulnerability were linked to an increased risk of COVID-19 mortality (50, 51, 54, 69). Finally, mortality rates were more pronounced in areas with greater SE inequalities (53, 67, 76) (Table 1 and 2, Supplemental file 2).

#### Studies reporting on severe COVID-19

Two studies combined mortality and intensive care unit admissions in a composite score for severe COVID-19 outcomes (31, 40). Both studies were conducted in a healthcare setting in the UK. One study is a case-control study with a sample size of 41,220 patients (31), while the other study is a cohort study with a sample size of 120,075 patients (40).

The study conducted by McKeigue presented unadjusted findings, which indicated that patients residing in care homes and experiencing higher levels of socio-economic deprivation exhibited more unfavourable outcomes in relation to COVID-19 (31). The study by Mutambudzi reported adjusted risk ratios for occupation, socio-economic deprivation, ethnicity and education. Severe COVID-19 cases were seven times higher in healthcare worker compared to non-essential workers, Black and south Asian had a higher risk compared to white (40) (Table 1 and 2, Supplemental file 2).

### Prognostic role of socio-economic characteristics

#### Studies reporting on hospitalisations

Seven studies reported on the prognostic role of socio-economic factors in hospitalisation of infected subjects (86, 89, 95, 99, 105, 114, 116). Five were cohort studies and two were cross-sectional. The studies were conducted exclusively in high-income nations, specifically the UK, USA, Italy, and Norway. One study was conducted in small statistical areas in Utah, whereas the other six were conducted at the individual level, with sample sizes ranging between 1,052 and 418,794. The patients included in the studies were COVID-19 community cases (89, 95, 99, 105, 116), nursing home residents (114) and patients from a large integrated US health system (86, 121). The socio-economic factors considered were the following: ethnicity, race, place of birth, income (both median and average), Townsend deprivation index, and deprivation level. Age, sex and comorbidities were the most used adjustment factors.

The likelihood of hospitalisation in COVID-19 patients was higher in Black, Asian, Hispanic patients (86, 95, 99, 105, 114). Non-Italians were at higher risk of hospitalisations compared to Italians after adjusting for sociodemographic factors and comorbidities in an Italian study (89). Three studies found increased odds of hospitalisation with SE deprivation (86, 99, 116) (Table 3 and 4, Supplemental file 2).

#### Studies reporting on ICU admission

Four individual-level studies (89, 98, 105, 111) and one ecological reported on ICU admission (106) . The studies were completed in Italy, Kuwait, Norway and USA. Regarding study design, we identified three cohort studies, one cross-sectional and one case-control. The socio-economic determinants used included: race, nationality, ethnicity, poverty, education, density, and income. Two studies included hospitalised COVID-19 cases while three included community COVID-19 cases. Only 18.7% of females included in the Kuwait study by Hamadah *et al.* (98). In individual-level studies, the sample size ranged between 1,123 and 518,739. Loomba *et al.* used state-level data to study socio-economic patterns in 205 paediatric COVID-19 positive admissions and included results adjusted for age, gender and comorbidities (106). Overall, patients with higher educational level were at lower risk of being admitted to the ICU for COVID-19 (111). Black and foreign-born populations were at higher risk of severity when compared with White and native-born patients respectively (89, 98, 105, 111) (Table 3 and 4, Supplemental file 2)..

#### Studies reporting on mechanical ventilation

Three studies in Norway, Brazil, and the USA documented the use of mechanical ventilation. Two studies used a cohort design (95, 96), while the other used a cross-sectional approach (105). The first study had a smaller sample size (N= 8,569) (95) than the other two studies (113,314 (96) and 124,780 (105)). Race, ethnicity, birthplace and education were the socio-economic factors considered. Two studies included community COVID-19 cases (95, 105), whilst another included hospitalised COVID-19 patients (96). Compared to patients born in Norway, non-native patients born in Africa, Asia, Latin-America were at higher risk of mechanical ventilation (95). Studies from the USA and Brazil reported lower risk of severe outcomes in White patients and patients with higher educational attainment (96, 105) (Table 3 and 4, Supplemental file 2).

#### Studies reporting on mortality

Twenty studies reported on the prognostic role of socio-economic determinants in mortality. Eighteen were individual-level studies (88, 89, 91, 92, 95–98, 100–105, 112, 113, 115, 117) and two were ecological (108, 110). Studies were performed in the USA, the UK, Brazil, Columbia and Mexico. The sample sizes for the individual-level studies varied from 1,123 to 1,033,218 participants, including both community and hospitalised COVID-19 patients.

The included determinants were ethnicity, race nationality, place of birth, employment, deprivation, education, insurance, urbanisation, and migration. Higher COVID-19 mortality was observed in Black and mixed races COVID-19 patients in Brazil, the USA, the UK (91, 103, 105, 113, 117) and, patients from Africa, Asia and Latin America compared to Norwegian citizens (95). Compared to non-Indigenous, the mortality risk was considerably greater for Indigenous populations at large and those who received ambulatory care in Mexico and Columbia (88, 102). One study from Brazil showed opposing results where Black patients had 25% lower odds of mortality compared to their White counterparts (92). Fabiani *et al.* did not observe any differences between Italian and non-Italian nationals in terms of case fatalities except for non-Italians coming from low Human Development Index (HDI) (89). A lower educational level (91, 96, 113), low socioeconomic status (88), working as a farmer, blue collar, being retired, unemployed or being a prisoner predicted higher mortality risk (104, 112). Both urbanisation and migration were associated with an increased risk of COVID-19-related mortality (115). Living in rural areas showed contrasting results. For instance, Kim *et al.’s* study revealed that patients residing in rural areas had a greater risk of death (97) while the study by Cifuentes *et al.* reported a decreased risk of mortality when living in sparse rural areas (88) .

The two remaining studies included aggregate data on the following determinants: ethnicity, education, median family income, poverty level, race, insurance, employment and urbanisation (108, 110) and included data from county-level and zip codes. The proportion of Black people in a zip code was not associated with death in COVID-19 patients (110). The odds of COVID-19 fatality decreased when the rate of primary physicians increased. After controlling for health factors, healthcare access measures, and county-level demographic characteristics, the association between race or ethnicity and COVID-19 death was not significant (108) (Table 3 and 4, Supplemental file 2).

#### Studies reporting on survival

Seven cohort studies (85, 90, 93, 94, 107, 109, 114) and one cross-sectional (87) performed in China, Brazil, Mexico, the UK and the USA reported survival outcomes. Seven studies were performed on an individual-level whereas one was ecological (107). The sample size ranged between 9,990 and 412,017 individuals including confirmed COVID-19 cases, nursing home residents and hospitalised adults due to COVID-19. The studies reported the following socio-economic determinants: ethnicity, race, area of residence, education, health insurance, occupation, socio-economic strata, deprivation and poverty level. Higher mortality was observed in Black, Asian and Indigenous populations in Mexico and Brazil (85, 87, 93, 94), rural zones, and COVID-19 patients with lower educational level (93, 94). Poverty level also increased the risk of COVID-19 death (90, 107). Mehta *et al.* reported higher mortality risk, among nursing home residents in the US, in Asian populations however, the risk of mortality for Black people was similar to white patients (114) (Table 3 and 4, Supplemental file 2).

## 4. DISCUSSION

This systematic review summarised the etiological and prognostic role of socio-economic determinants in various COVID-19 outcomes during the early phase of the pandemic. During the early stages of the pandemic, most the studies were performed in high-income countries and focusing on short-term outcomes. Although the selected studies included a comparable proportion of males and females, their sample size consisted mainly of white populations. Studies adjusted for age, sex, and comorbidities among other adjustment factors. Socio-economic characteristics were important features, alongside comorbidities, for predicting SARS-CoV-2 infection and COVID-19 severity. Black, Hispanic and Asian populations were at higher risk of SARS-CoV-2 infection, COVID-19 related hospitalisation, ICU admission and death, in Brazil, the USA and the UK. The infection and severity risk was also consistently higher with higher socio-economic deprivation. Higher levels of education were associated with higher odds of COVID-19 diagnosis but with lower odds of worsening outcomes. During the first wave of the pandemic, higher education was also associated with higher incidence, but this trend reversed itself in the subsequent waves. Our results provide strong evidence that inequality seems to be proportionally more important for a severe course of the disease.

The positive association between socio-economic disparity and worse COVID-19 outcomes has been also documented in many publications published after our review. We found additional studies completed in Brazil, Canada, France, Indonesia, Italy, Norway, Switzerland, and the US reporting on the role of socioeconomic factors in COVID-19 incidence and severity (122–131). Population-based studies from Brazil showed that higher risk of COVID-19 mortality was observed among Indigenous, Black and mixed-race patients and in more deprived areas (124–126). Lower income, lower educational level, crowded housing, larger household size, poverty and high density area were also predictors for SARS-CoV-2 infection (128) and COVID-19 mortality (123, 126, 129). According to Pereira *et al*., COVID-19 incidence and mortality did not exhibit similar results: SARS-CoV-2 infection were higher in less deprived areas (125). COVID-19 impact was not only commanded by race and ethnicity but was highly impacted by patients’ socio-ethnic and socioracial profile. Vulnerable patients suffering from worsening COVID-19 outcomes need higher medical attention and financial resources adding constraints for their access to treatment (125). Results from Switzerland and France, until April 2021, show different results as patients from disadvantaged socio-economic position were more likely to test positive to SARS-CoV-2 infection but less likely to get tested (127, 130). A possible explanation for these findings is structural barriers to healthcare access and deprived populations’ lower ability to benefit from protective measures (127, 130). This could also be potentially explained by the “inverse equity hypothesis”: new public health interventions, such as SARS-CoV-2 testing or other related preventive measures, are more utilised by people from higher socio-economic position which leads to increasing inequity in terms of coverage, morbidity and mortality (132). Furthermore, there is a large gap in the geographical distribution of the included articles where 60% of the articles were performed in the USA and the UK. There is a need for population-based studies on the role of socio-economic determinants in Africa, Asia, continental Europe and Latin America.

Given the positive association between area-level social determinants of health and COVID-19 mortality, it remains essential to put in place targeted strategies to address the population that are disproportionally affected by the pandemic such as essential workers who did not have access to telework and were less likely to receive benefits or paid sick leave (131). Our results, covering the heterogeneity of outcomes over different phases, particularly in terms of education, are also supported by a population-based study conducted in Rome, demonstrating that the relationship between education level and infection rates varied across different waves (122). During the initial stage of the pandemic, individuals with lower and medium levels of education had a reduced risk of infection or lower testing rates, which later increased. Likewise, in the last semester of 2020, there was strong evidence that lower education was associated with an increased risk of death within 30 days of infection onset (122).

Two other systematic reviews focusing on the population from the US had shown a disproportionate burden of COVID-19-related infections and mortality experienced by Black and Hispanic populations and in areas with higher deprivation (133, 134). While these findings are consistent with our review, they are limited to the US and do not specifically address population-based studies, proving that no previous systematic review has examined the relationship between socio-economic characteristics and the severity of COVID-19 in the general population. Our review, therefore managed to fill this gap (133, 134).

Our findings show substantial social inequalities in SARS-CoV-2 infection and COVID-19 outcomes. Collecting socioeconomic data in a routine and timely manner is essential to identify better the mechanisms driving such inequalities and design preventive strategies. Addressing socio-economic health disparities is a key focus in public health, but has not been adopted in disease surveillance and pandemic preparedness (7). Countries, should, therefore, establish surveillance systems that include social variables to routinely report health outcomes by social factors routinely (130). Whilst most of the studies reported on infection and mortality outcomes, it remains important to investigate the role of socio-economic characteristics on hospitalisation and other comparable outcomes to understand better the patterns across the whole continuum of care.

Our study has several strengths: first, using a pre-registered protocol, which was peer-reviewed and published thereby guaranteeing a rigorous and reproducible systematic process (9, 135). Second, it includes representative studies allowing the estimation of COVID-19 risk factors at a population level. Third, this review employed a comprehensive search strategy, and fourth, included an extensive number of articles covering a large number of socio-economic determinants and their impact on COVID-19 outcomes during the early phase of the pandemic.

One limitation of our study is that it relies solely on English-language peer-reviewed articles. This may have resulted in excluding population-based studies or grey literature published in other languages. The socio-economic characteristics included were heterogeneous with various definitions and stratifications, resulting in difficult comparison between the reported results. Another drawback is that many included articles considered potential mediators as confounding factors such as comorbidities, which could have caused an over-adjustment bias and decreased the estimates of risk linked to socio-economic determinants. Controlling for comorbidities is unnecessary in this context because they serve as intermediary variables along the causal pathway between socio-economic factors and health outcomes (136). We are also aware that our search strategy covers only articles until April 202. However, our study aims to look at the scientific literature published in the early phase of the pandemic, focusing on a limited number of variants and avoiding the impact of vaccination, which could potentially act as a mediator and mitigate certain outcomes. Hence, our study addresses the direct interaction between socioeconomic characteristics and COVID-19 morbidity and mortality. After extracting 100 articles we also concluded that we reached a saturation of results and that an update was redundant to cover this period.

## 5. CONCLUSION

This review underscores the significant correlation between socio-economic disadvantages, ethnicity, and the severity of COVID-19 outcomes. Particularly, individuals identifying as Black, Asian, or Indigenous are more vulnerable to both infection and adverse prognosis. There is a lack of evidence on long-term COVID-19 outcomes despite growing evidence on the association between deprivation and “long COVID-19” (137). Future research should prioritise these high-risk groups in order to inform and refine intervention strategies, aiming to bridge disparities in health outcomes. Moreover, it’s essential for surveillance systems to broaden their scope, incorporating a wider array of socio-economic factors to capture the full spectrum of determinants impacting health outcomes.

## 6. Authors Contribution

LCB, JCo, BD, PJN and MJF formulated the research questions. All authors conceptualised the search strategy together. JG and TM acted as first reviewers, while LCB, SMB, LC, RH, RFS, FPB, JC, MA, BV, PB, PJN shared the role of the second reviewer. JCo acted as a third party and resolved all disagreements. JG authored the initial draft of the manuscript. All co-authors reviewed and approved the manuscript.

## Data Availability

All data produced are available online

