## Supplemental file 1 for "The role of socio-economic determinants in SARS-CoV-2 health outcomes: systematic review of population-based studies"

**Pubmed strategy**

| #1 | Covid-19(1) | (((((((((((((((((((((("Betacoronavirus"[MeSH Terms] OR "Coronavirus Infections"[MeSH Terms]) OR "COVID-19"[Supplementary Concept]) OR "Coronavirus"[MeSH Terms]) OR "Severe Acute Respiratory Syndrome Coronavirus 2"[Supplementary Concept]) OR "2019nCoV"[All Fields]) OR "betacoronavirus*"[All Fields]) OR "corona virus*"[All Fields]) OR "coronavirus*"[All Fields]) OR "coronovirus*"[All Fields]) OR "CoV"[All Fields]) OR "CoV2"[All Fields]) OR "COVID"[All Fields]) OR (("COVID-19"[Supplementary Concept] OR "COVID-19"[All Fields]) OR "covid19"[All Fields])) OR ((((((("COVID-19"[All Fields] OR "covid 2019"[All Fields]) OR "Severe Acute Respiratory Syndrome Coronavirus 2"[Supplementary Concept]) OR "Severe Acute Respiratory Syndrome Coronavirus 2"[All Fields]) OR "2019 ncov"[All Fields]) OR "SARS CoV 2"[All Fields]) OR "2019nCoV"[All Fields]) OR (("wuhan"[All Fields] AND ("Coronavirus"[MeSH Terms] OR "Coronavirus"[All Fields])) AND (2019/12/1:2019/12/31[Date - Publication] OR 2020/1/1:2020/12/31[Date - Publication])))) OR "HCoV-19"[All Fields]) OR "nCoV"[All Fields]) OR "SARS CoV 2"[All Fields]) OR "SARS2"[All Fields]) OR "SARSCoV"[All Fields]) OR (((("sars virus"[MeSH Terms] OR ("sars"[All Fields] AND "virus"[All Fields])) OR "sars virus"[All Fields]) OR ("sars"[All Fields] AND "CoV"[All Fields])) OR "sars cov"[All Fields])) OR (("Severe Acute Respiratory Syndrome Coronavirus 2"[Supplementary Concept] OR "Severe Acute Respiratory Syndrome Coronavirus 2"[All Fields]) OR "SARS CoV 2"[All Fields])) OR "severe acute respiratory syndrome cov*"[All Fields]) AND (2019/11/17:3000/12/31[Date - Entry] OR 2019/11/17:3000/12/31[Date - Publication]) OR "COVID-19"[MeSH Terms] OR "SARS-Cov-2"[MeSH Terms]  OR "SARS CoV-2" OR "SARS-CoV-2" OR SARSCoV2 OR "CoV-2" OR "covid 19" OR covid2019 OR "covid-2019" OR "novel CoV" OR "corona pandemic*" OR "wuhan virus*" OR "CoV 2" OR ((wuhan OR hubei OR huanan) AND ("severe acute respiratory" OR pneumonia*) AND outbreak*) |
| --- | --- | --- |
| #2 | Frailty | frailty OR frail OR frailty[MeSH Terms] |
| #3 | Multimorbidity(2) | multimorbidity OR "multi-morbidity" OR "multi morbidity" OR multimorbidities OR "multi-morbidities" OR "multi morbidities" OR multimorbid OR "multi-morbid" OR "multi morbid" OR comorbidity OR "co-morbidity" OR "co morbidity" OR comorbidities OR "co-morbidities" OR "co morbidities" OR comorbid OR "co morbid" OR "multiple chronic conditions" OR "multiple chronic diseases" OR "multiple conditions" OR "multiple diseases" OR "multiple disorders" OR polymorbid* OR "poly-morbid*" OR "poly morbid*" OR polypath* OR pluripath* OR multipath* OR "multi path*" OR "multi-path*" OR "multiple pathologies" OR "disease cluster" OR "disease clusters" OR "disease pattern" OR "disease patterns" OR "concurrent chronic diseases" OR "multiple chronic disorders" OR multimorbidity[MeSH Terms] OR comorbidity[MeSH Terms] |
| #4 | Socioeconomic characteristics | "socio economic" OR "socio-economic" OR socioeconomics OR "socio-economics" OR "socio economics" OR socioeconomic OR "social difference" OR "social differences" OR "social inequality" OR "social inequalities" OR "socioeconomic inequality" OR "socioeconomic inequalities" OR "social disparity" OR "social disparities" OR education OR literacy OR "socioprofessional" OR "socio-professional" OR "socio professional" OR "social conditions" OR "social class" OR "social classes" OR "social class"[MeSH Terms] OR "socioeconomic factors"[MeSH Terms] OR health status disparities[MeSH Terms] OR income OR poverty OR deprivation OR rural OR urban OR "housing deprivation" OR homeless OR houseless OR homelessness OR ethnic* OR race OR emigrant OR immigrant OR migrant OR "minority group" OR "minority groups" OR disadvantaged OR "marital status" OR ((characteristics OR factors OR status) AND (economic OR social OR educational)) OR ((composition OR characteristics OR size) AND (family OR household)) |
| #5 | Study design | "cross-sectional" OR "cross sectional" OR "case-control" OR "case control" OR cohort OR longitudinal OR "ecological study" OR "ecological studies" OR "ecological design" OR "ecological designs" OR observational OR "observational study" OR "observational studies" OR "observational design" OR "observational designs" OR "prospective study" OR "prospective studies" OR "prospective design" OR "prospective designs" OR "retrospective study" OR "retrospective studies" OR "retrospective design" OR "retrospective designs" OR "prospective observational study" OR "prospective observational studies" OR "retrospective observational study" OR "retrospective observational studies" OR case-control studies[MeSH Terms] OR cohort studies[MeSH Terms] OR cross-sectional studies[MeSH Terms] |
| #6 |  | #2 OR #3 OR #4 |
| #7 |  | #1 AND #5 AND #6 |

**COVID-19 Global literature on coronavirus disease strategy**

((tw:("socio economic" OR "socio-economic" OR socioeconomics OR "socio-economics" OR "socio economics" OR socioeconomic OR "social difference" OR "social differences" OR "social inequality" OR "social inequalities" OR "socioeconomic inequality" OR "socioeconomic inequalities" OR "social disparity" OR "social disparities" OR education OR literacy OR "socioprofessional" OR "socio-professional" OR "socio professional" OR "social conditions" OR "social class" OR "social classes" OR "health status disparity" OR "health status disparities" OR income OR poverty OR deprivation OR rural OR urban OR "housing deprivation" OR homeless OR houseless OR homelessness OR ethnic$ OR

race OR emigrant OR immigrant OR migrant OR "minority group" OR "minority groups" OR disadvantaged OR "marital status" OR ((characteristics OR factors OR status) AND (economic OR social OR educational)) OR ((composition OR characteristics OR size) AND (family OR household)))) OR (tw:(multimorbidity OR "multi-morbidity" OR "multimorbidity" OR multimorbidities OR "multi-morbidities" OR "multi morbidities" OR multimorbid OR "multi-morbid" OR "multi morbid" OR comorbidity OR "co-morbidity" OR "co morbidity" OR comorbidities OR "co-morbidities" OR "co morbidities" OR comorbid OR "co morbid" OR "multiple chronic conditions" OR "multiple chronic diseases" OR "multiple conditions" OR "multiple diseases" OR "multiple disorders" OR polymorbid$ OR "poly-morbidity" OR "poly-morbidities" OR "poly morbidity" OR "poly morbidities" OR polypath$ OR pluripath$ OR multipath$ OR "multi pathology" OR "multi pathologies" OR "multi-pathology" OR "multi-pathologies" OR "multiple pathologies" OR "disease cluster" OR "disease clusters" OR "disease pattern" OR "disease patterns" OR "concurrent chronic diseases" OR "multiple chronic disorders" )) OR (tw:(frailty OR frail ))) AND tw:("cross-sectional" OR "cross sectional" OR "case-control" OR "case control" OR cohort OR longitudinal OR "ecological study" OR "ecological studies" OR "ecological design" OR "ecological designs" OR observational OR "observational study" OR "observational studies" OR "observational design" OR "observational designs" OR "prospective study" OR "prospective studies" OR "prospective design" OR "prospective designs" OR "retrospective study" OR "retrospective studies" OR "retrospective design" OR "retrospective designs" OR "prospective observational study" OR "prospective observational studies" OR "retrospective observational study" OR "retrospective observational studies" )

**PsycINFO strategy**

S1

"cross-sectional" OR "cross sectional" OR "case-control" OR "case control" OR cohort OR longitudinal OR "ecological study" OR "ecological studies" OR "ecological design" OR "ecological designs" OR observational OR "observational study" OR "observational studies" OR "observational design" OR "observational designs" OR "prospective study" OR "prospective studies" OR "prospective design" OR "prospective designs" OR "retrospective study" OR "retrospective studies" OR "retrospective design" OR "retrospective designs" OR "prospective observational study" OR "prospective observational studies" OR "retrospective observational study" OR "retrospective observational studies" OR

DE("Longitudinal Studies" OR "Followup studies" OR "Retrospective studies" OR "Cohort analysis")

S2

"socio economic" OR "socio-economic" OR socioeconomics OR "socio-economics" OR "socio economics" OR socioeconomic OR "social difference" OR "social differences" OR "social inequality" OR "social inequalities" OR "socioeconomic inequality" OR "socioeconomic inequalities" OR "social disparity" OR "social disparities" OR education OR literacy OR "socioprofessional" OR "socio-professional" OR "socio professional" OR "social conditions" OR "social class" OR "social classes" OR "health status disparity" OR "health status disparities" OR income OR poverty OR deprivation OR rural OR urban OR "housing deprivation" OR homeless OR houseless OR homelessness OR ethnic* OR

race OR emigrant OR immigrant OR migrant OR "minority group" OR "minority groups" OR disadvantaged OR "marital status" OR ((characteristics OR factors OR status) AND (economic OR social OR educational)) OR ((composition OR characteristics OR size) AND (family OR household)) OR DE ("Disadvantaged" OR "Economic Disadvantage" OR "Economic Inequality" OR "Economic Resources" OR "Family Socioeconomic Level" OR "Income (Economic)" OR "Lower Class" OR "Poverty" OR "Rural Health" OR "Socioeconomic Status" OR "Socioeconomic Factors" OR "Social Class" OR "Urban Health" OR "Racial and Ethnic Differences" OR "Racial Disparities" OR "Health Disparities")

S3

multimorbidity OR "multi-morbidity" OR "multi morbidity" OR multimorbidities OR "multi-morbidities" OR "multi morbidities" OR multimorbid OR "multi-morbid" OR "multimorbid" OR comorbidity OR "co-morbidity" OR "co morbidity" OR comorbidities OR "co-morbidities" OR "co morbidities" OR comorbid OR "co morbid" OR "multiple chronic conditions" OR "multiple chronic diseases" OR "multiple conditions" OR "multiple diseases" OR "multiple disorders" OR polymorbid* OR "poly-morbid" OR "poly-morbidity" OR "poly-morbidities" OR "poly morbid" OR "poly morbidity" OR "poly morbidities" OR polypath* OR pluripath* OR multipath* OR "multi pathology" OR "multi pathologies" OR "multi-pathology" OR "multi-pathologies" OR "multiple pathologies" OR "disease cluster" OR "disease clusters" OR "disease pattern" OR "disease patterns" OR "concurrent chronic diseases" OR "multiple chronic disorders" OR Comorbidity [DE]

S4 Frailty OR frail

S5

"covid-19" OR "2019-ncov" OR "sars cov 2" OR "cov-19" OR cov19 OR "cov 19" OR 2019nCoV OR betacoronavirus* OR "corona virus" OR coronavirus* OR coronovirus* OR CoV OR CoV2 OR COVID OR covid19 OR "covid 2019" OR "Severe Acute Respiratory Syndrome Coronavirus 2" OR "2019 ncov" OR (wuhan AND Coronavirus) OR "HCoV-19" OR nCoV OR SARS2 OR SARSCoV OR (sars AND virus) OR "sars virus" OR (sars AND CoV) OR "sars cov" OR "severe acute respiratory syndrome cov" OR "SARS CoV-2" OR "SARS-CoV-2" OR SARSCoV2 OR "CoV-2" OR "covid 19" OR covid2019 OR "covid-2019" OR "novel CoV" OR "corona pandemic" OR "wuhan virus" OR "CoV 2" OR ((wuhan OR hubei OR huanan) AND ("severe acute respiratory" OR pneumonia*) AND outbreak*) OR COVID-19 [DE]

S6

(S2 OR S3 OR S4) AND S5 AND S1

**Embase strategy**

((comorbid* OR 'comorbidity'/exp OR comorbidity OR 'multiple chronic conditions'/exp OR 'multiple chronic conditions' OR multimorbid* OR 'multi morbid' OR 'multi morbidity' OR 'multi morbidities' OR 'co morbid' OR 'co morbidity' OR 'co morbidities' OR 'disease pattern' OR 'disease patterns' OR 'disease cluster' OR 'disease clusters' OR 'multiple conditions' OR 'multiple diseases' OR 'multiple disorders' OR polymorbid* OR 'poly morbid' OR 'poly morbidity' OR 'polymorbidities' OR polypath* OR 'multiple pathology' OR 'multiple pathologies') OR ('frail elderly'/exp OR 'frail elderly' OR 'frailty'/exp OR 'frailty' OR frail*) OR (('socioeconomics'/exp OR 'socioeconomics' OR 'social status'/exp OR 'social status' OR 'social stratification'/exp OR 'social stratification' OR 'social class'/exp

OR 'social class') OR ('urban area'/exp OR 'urban area' OR 'rural area'/exp OR 'rural area') OR 'population group'/exp OR ('minority group'/exp OR 'minority group' OR 'homelessness'/exp OR 'homelessness' OR 'migrant'/exp OR 'migrant' OR 'race'/exp OR 'race' OR ethnic* OR 'minority groups') OR ('economic status'/exp OR 'economic status' OR 'income'/exp OR 'income' OR 'income group'/exp OR 'income group') OR ('social discrimination'/exp OR 'social discrimination' OR 'socioeconomic inequality'/exp OR 'socioeconomic inequality' OR 'health disparity'/exp OR 'health disparity' OR 'poverty'/exp OR 'poverty' OR 'income inequality'/exp OR 'income inequality') OR ('educational status'/exp OR 'educational status' OR 'literacy'/exp OR 'literacy' OR 'education* level') OR

('family size'/exp OR 'family size' OR 'household characteristics' OR 'household size' OR 'household'/exp))) AND ('longitudinal study'/exp OR 'longitudinal study' OR 'cross-sectional study'/exp OR 'cross-sectional study' OR longitudinal OR 'cross sectional' OR 'case control study'/exp OR 'case control study' OR 'case control' OR cohort OR 'ecological study' OR 'ecological design' OR 'observational study'/exp OR 'observational study' OR 'retrospective study'/exp OR 'retrospective study' OR 'prospective study'/exp OR 'prospective study' OR 'prospective observational study' OR 'prospective observational studies' OR 'retrospective observational study' OR 'retrospective observational studies' OR 'ecological designs' OR 'prospective studies' OR 'retrospective studies' OR 'observational studies' OR 'ecological studies') AND ('coronavirus disease 2019'/exp OR 'coronavirus disease 2019' OR 'covid 2019' OR 'covid 19' OR covid2019 OR covid19 OR sarscov2 OR 'sars cov 2' OR 'sars cov2' OR 'severe acute respiratory syndrome'/exp OR 'severe acute respiratory syndrome' OR 'wuhan virus' OR 'cov 2' OR cov2 OR coronavirus OR 'corona virus' OR 'novel cov' OR 2019ncov OR '2019 ncov' OR 'corona pandemic' OR betacoronavirus OR cov19 OR 'cov 19' OR 'hcov-19' OR ncov OR sars2) AND [english]/lim AND [2020-2021]/py
