## Supplemental file 2 for "The role of socio-economic determinants in SARS-CoV-2 health outcomes: systematic review of population-based studies"

##### Table 1. Characteristics of studies reporting association on socioeconomic determinants among population representatives samples – Etiological role

| **First author (Country)** | **Study design Study setting Study period** | **Study quality score*** | **Follow-up period** | **Covid-19 diagnostics for population selection** | **Outcome measures** | **Study participants included in the analyses and available socio-demographic characteristics** | **List of socioeconomic determinants considered in the study** | **Socioeconomic determinants (definition, measurement, levels and the source of this information; prevalence where available)** | **Association metrics between socioeconomic determinants and the outcome; ; analyses Adjusted (yes/no/both)** | **Total N (%) of people for which the outcome occurred** |
| --- | --- | --- | --- | --- | --- | --- | --- | --- | --- | --- |
| **Individual-level studies** | | | | | | | | | | |
| Hanson et al.  2020    (USA) | Cross-sectional study  Patients at Indiana University Health or Eskenazi Health hospitals  January 1, 2020 – April 30 2020 | 7 | NA | Nasopharyngeal swab | COVID-19 diagnosis  COVID-19 cases based on zip code  characteristics | N = 3 892 | Race  Ethnicity | Race:   - African American - Non-African American   Ethnicity:   - Hispanic - Non-Hispanic | OR (95% CI)  Adjusted | 3 892 COVID-19 cases |
| Bergman et al. 2021  (Sweden) | Case-control  Community  January 30 2020 – September 27 2020 | 6 | NA | No info on the methods of testing  International Classification of Diseases, 10th Revision, Swedish Version, [ICD-10-SE) Code: U071 | COVID-19 diagnosis without hospitalisation, non-ICU hospitalisation with confirmed COVID-19, ICU hospitalization for confirmed COVID-19 | N = 518 739  Control = 434081 Cases = 84658  Female 49.15% | Country of Birth  Education  Disposable family income in 2018 | Country of Birth   - Sweden N = 410 092 (79.06%) - Not born in Sweden (20.94%)   Education   - Primary N = 83 206 (16.04%) - Secondary N = 178 629 (34.44%) - Post-secondary < 3y N = 58 601 (11.3%) - Post-secondary> 3y N = 93 212 (17.96%) - Missing N = 105 091 (20.26%)   Disposable family income in 2018   - Quintile 1 N = 85 870 (16.55%) - Quintile 2 N = 84 931 (16.37%) - Quintile 3 N = 83 641 (16.12%) - Quintile 4 N = 86 044 (16.6%) - Quintile 5 N = 87 423 (16.85%) - Missing N = 90 830 (17.51%) | OR (95% CI)  Both | 68 575 infections (13.22%)  13 586 Non-ICU hospitalisations (2.62%)  2494 ICU hospitalisations (0.48%) |
| Brandén et al.  2020  (Sweden) | Cohort  Community- All individuals living in Stockholm county  InclUSA ion on March 12 2020 | 9 | Follow-up until May 5 2020 | Data from the cause-of-death register using codes [ICD) version 10 codes U07.1, U07.2, or B34·2) | COVID-19 mortality | N = 274 712  Female 55.8%; Sweden 79.3%; LMIC MENA 2.7%; Other LMIC 4.3%; HIC 13.7%;  % Education > HS 75.3%;  Low income 42.1%; Middle income 35.8%; High income 22.1%;  Living alone 40.3% | m² per individual in household  Housing  Country of birth  Educational level  Individual disposable income | m² per individual in household   - 0 to < 20 N = 7 859 (2.9%) - 20 to <30 N = 19 323 (7%) - 30 to < 40 N = 43 278 (15.8%) - 40 to < 60 N = 89 081 (32.4%) - >=60 N = 114 228 (41.6%) - Missing N = 943 (0.3%)   Housing   - Multi-family housing N = 173 179 (63%)   - Single family housing N = 90 655 (33%)   - Care home N = 10 878 (4%)   Country of birth   - Sweden N = 217 785 (79·3%) - HIC N = 37 517 (13·7%) - LMIC MENA N = 7 550 (2·7%) - LMIC others N = 11 860 (4·3%)   Educational level   - Primary N = 61 988 (22·6%)   - Secondary N = 106 894 (38·9%)   - Post-secondary N = 99 875 (36·4%)   - Missing N = 5 955 (2·2%)   Individual disposable income   - Lowest tertile N = 115 518 (42·1%) - Mid tertile N = 98 421 (35·8%) - Highest tertile N = 60 773 (22·1%) | HR (95% CI)  Adjusted | 1301 deaths (0.47%) |
| Guijarroa et al.  2021  (Spain) | Cohort  All adults registered in the official municipal population registry of the City Council of Alcorcón  February 1 2020 – April 25 2020 | 7 | 2 months | RT-PCR | COVID-19 incidence rates  Cumulative incidence  COVID-19 hospitalisations | N = 152 018  Median age 71; IQR (54-79);  Female 42.5%; | Region of origin | - Spain N = 131 599 (86.57%) - European Union N = 7 658 (5.04%) - Eastern Europe and Russia N = 2 058 (1.35%) - Asia N = 1 439 (0.95%) - North Africa N = 1 952 (1.28%) - Sub Saharan Africa N = 801 (0.53%) - Latin America N = 5 826 (3.83%) - Caribbean N = 657 (0.43%) - USA of America and Canada N = 118 (0.08%) | RR (95% CI)  Adjusted | 1 036 COVID-19 cases (0.68%)  877 COVID-19 hospitalisations (0.58%) |
| Chadeau-Hyam et al. 2020  (UK) | Cohort  Community  March 16 2020 – May 18 2020 | 7 | NA | RT-PCR | COVID-19 tests  COVID-19 cases | N = 4509  Mean age (± SD) 68.64 (±8.88);  Female 51.19%  Black 3.73%;  White 90.74%;  Other race / ethnicity 5.53%;  ≥ High School Education 76.6% | Ethnicity  Education  Type of accomodation  Own or rent accomodation  Number in household  Income  Occupation | Ethnicity   - White N = 4067 (90.20%) - Black N = 167 (3.70%) - Other N =248 (5.50%)   Education   - High N = 1250 (27.72%) - Intermediate N = 2108 (46.75%) - Low N = 1023 (22.69%)   Type of accommodation   - House N = 3826 (84.85%) - Flat N = 599 (13.28%)   Own or rent accommodation   - Own outright Own N = 2057 (45.62%) - Own with a mortgage N = 1535 (34.04%) - Rent N = 760 (16.86%)   Number in household N= 4421 (98.05%)  Number in household  Income   - less than 18 000 N =1197 (26.55%) - 18 000 to 30999 N = 918 (20.36%) - 31 000 to 51 999 N = 834 (18.50%) - greater than 52 000 N = 789 (17.50%)   Occupation   - Employed (other) N = 10497 (232.80%) - Healthcare worker N = 628 (13.93%) - Unemployed N = 784 (17.39%) - Retired N = 1542 (34.20%) | OR ( CI 95%I)  Both | 1325 COVID-19 cases (32.92%) |
| Clift et al.  2020  (UK) | Cohort  Primary care patients  Inclusion on January 24 2020 | 7 | Follow-up until April 30 2020 | RT-PCR | Time to death from COVID-19  Time to hospital admission with confirmed SARS-CoV-2 infection | N=6 083 102  Mean age (± SD) 48.21(±18.57); Female 51.1%;  Black 2.47%; Asian 8.9%; White  64.51%;  Other 3.69%; Caribbean 1.14% | Ethnicity  Accommodation  Townsend deprivation fifth | Ethnicity   - White N=3924110 (64.51%) - Indian N=175909 (2.89%) - Pakistani N=114727 (1.89%) - Bangladeshi N=87491 (1.44%) - Other Asian N=110579 (1.82%) - Caribbean N=69166 (1.14%) - Black African N=150022 (2.47%) - Chinese N=58511 (0.96%) - Other ethnic groups N=224394 (3.69%) - Not recorder N=1168193 (19.20%)   Accommodation   - Not in care home or homeless N=6036288 (99.23%) - Lives in care home or homeless N=35813 (0.59%) - Homeless according to GP records N=11001 (0.18%)   Townsend deprivation fifth   - 1 (most affluent) N=1238575 (20.36%) - 2 N=12228575 (20.10%) - 3 N=1187082 (19.51%) - 4 N=1176829 (19.35%) - 5 (most deprived) N=1231431 (20.24%) - Not recorded N=26504 (0.44%) | HR (95%CI) Adjusted | 10776 hospitalizations (0.18%)  4384 deaths (0.07%) |
| AyoubKhani et al.  2020  (UK) | Cohort  Community (residents of England and Wales)  2 March 2020-15 May 2020 | 7 | 2 March 2020-15 May 2020 | Death registered with an underlying cause, or any mention, of ICD-10 codes U07.1 (COVID-19, virus identified) or U07.2 (COVID-19, virus not identified) | COVID-19 mortality | N = 47 872 412  Age mean 46.5;  Female 51.6%;  Black 3.2%;  Asian 6.2%;  White 86.4%;  Other 4.3%;  Married / Cohabiting 71%;  Divorced / Seperated 13.5% | Ethnicity | - White N = 41 361 764; - Bangladeshi/Pakistani N = 1 436172 - Black N= 1 531 917 - Chinese N= 287 235 - Indian N= 1 244 683 - Mixed N = 957 448 - Other N = 1 053 193 | HR (95% CI)  Adjusted | NA |
| Drefahl et al.  2020  (Sweden) | Cohort  Community  March 12 2020 | 9 | March 13 2020 till May 7 2020 | Underlying cause of death (emergency ICD code U07.1, U07.2, or B34.2) | All deaths reported between March 13, 2020 and May 7, 2020, and association between death and COVID-19 | N = 7 775 054  Female 50.1%;  Country of birth: Sweden 79.5%; HIC 6.7%; LMIC MENA 5%; LMIC other 8.8%  ≥ High School Education 37.2%;  Middle income class 33.4%;  High income class 33.3%;  Married/ Cohabiting 42.5%; Divorced / Separated 12.8%;  Widowed 5.4%;  Never married 39.3% | Civil status  Education  Individual net income  Country of birth  County of residence | Civil status   - Married N= 3302934 (42.5%) - Never married N= 3059130 (39.3%) - Divorced N = 993609 (12.8%) - Widowed N = 419381 (5.4%)   Education   - Primary N = 1307280 (16.8%) - Secondary N = 3439340 (44.2%) - Post-secondary N = 2891664 (37.2%) - Missing N = 136 770 (1.8%)   Individual net income   - Tertile 1 (low) N = 2592086 (33.3%) - Tertile 2 N = 2593056 (44.2%) - Tertile 3 (high) N = 2589912 (33.3%)   Country of birth   - Sweden N = 6184398 (79.5%) - HIC N = 521804 (6.7%) - LMIC MENA N = 388483 (5%) - LMIC other N = 680369 (8.8%)   County of residence   - Stockholm N = 1,757,088 (22.6%) - Other N = 6,017,966 (77.4%) | HR (95% CI)  Both | 3126 deaths (0.04%) |
| Elimian et al. 2020  (Nigeria) | Cohort  Community  February 27 2020 – June 8 2020 | 7 | NA | RT-PCR | COVID-19 positivity | N = 36 496 | Occupation | Occupation   - Pupil/ student N = 2669 (9.7%) - Child N = 399 (1.4%) - Housewife N = 521 (1.9%) - Trader/bUSA iness N = 2441 (8.8%) - Health worker N = 4490 (16.2%) - Animal-related work N = 78 (0.3%) - Farmer N = 56 (2%) - Religious / traditional leader N = 95 (0.4%) - Transporter N = 195 (0.7%) - Other N = 16 205 (58.6%) | OR (95% CI)  Both | 10 517 COVID-19 cases (28.8%) |
| Izurieta et al.  2021  (USA) | Cohort  Medicare fee-for-service beneficiaries  April 1 2020 - May 8 2020 | 7 |  | ICD-10 and BNF chapter with severe COVID-19 | Hospitalisation  Mortality | N = 25 333 329  Female 55.6%;  Black 6.7%;  Hispanic 1.6 %;  Asian2.1 %;  White 85.1;  North American native : 0.5%  Other 4.1% | Ethnicity | - White REF - Black - Hispanic - North American Native - Asian - Other/Unknown | OR (95%CI)  Adjusted | 27,961 Hospitalizations (0.11%)  12 613 Deaths (0.05%) |
| Elliott et al.  2021  (UK) | Cohort  Community  January 31 2020 | 7 | January 31 2020-21 September 2020 | NA  National death registries  ICD-10 codes denoting COVID-19 death:  U07.1 , U07.2 | Number of COVID-19 deaths | N=473550  Females 55.26%; Black 1.64%; White 93.96%; Other 3.85% | Education  Type of accommodation  Own/rent  Number in household  Income  Occupation  Ethnicity | Education   - High N=154566 (32.64%) - Intermediate N=233115 (49.23%) - Low N=77601 (16.39%)   Type of accommodation   - House N=423584 (89.45) - Flat N=45841 (9.68%)   Own/rent   - Own outright N=214909 (45.38%) - Own with a mortgage N=1772248 (37.43%) - Rent N=43205 (9.12%)   Number in household N=469501  Income   - 18000 to 30999 N=87650 (18.51%) - Less than 18 000 N=101563 (21.45%) - 31000 to 51999 N=106461 (22.48%) - Greater than 52000 N=106330 (22.45%)   Occupation   - Employed N=237301 (50.11%) - Health care worker N=28029 (5.92%) - Unemployed N=64637 (13.65%) - Retired N=138142 (29.17%)   Ethnicity   - White N=449945 (95.02%) - Black N=7784 (1.64%) - Other N=18249 (3.85%) | OR (95% CI)  Both | 459 COVID-19 deaths (0.10%) |
| Kolin et al.  2020  (UK) | Cohort  Participants from England  between the ages of40 and 69 recruited from 2006 to 2010  March 16 2020 – May 18 2020 | 7 | NA | RT-PCR | Covid-19 positive status | N = 397 064  Mean age (± SD) 56.4 (±8.1);  Minimum – Maximum age 40 – 69;  Female 54.8;  Black 1.9%;  Asian 2.6%;  White 94.2%; | Race  Townsend deprivation index | Race   - Asian N = 10181 (2.6%) - Black N = 7431 (1.9%) - White N =374090 (94.2%)   Townsend deprivation index  Mean (SD) = -1.4 (3) | RR (95% CI)  Both | 968 COVID-19 cases (0;24%) |
| Bailey et al.  2020  (USA) | Cohort  Network paediatric health system  1 January 2020 – 8 September 2020 | 7 | 8 months | RT-PCR | SARS-CoV-2 infection | N = 135 794  Female 47.38% ;  Black 15% ;  Hispanic 11%;  Asian 3%  White 59%;  Other 9%;  Multiple 9% | Ethnicity | - White N= 79 625 - Black N= 20 189 - Hispanic N= 15 182 - Asian or Pacific Islander N= 4631 \| | OR (95% CI)  Adjusted | 4814 infections (3.55%) |
| Mak et al. 2021  (UK) | Cohort  Community (UK- Biobank)  March 16, 2020 - November 30, 2020 | 7 | NA | RT-PCR  ICD-10 | COVID-19 mortality | N = 410 199  Age range 67.6+/-8.1;  Female 86; Black 55.1%;  Hispanic 1.8%;  White 2.4%;  Other 94.3%;  High income 33% | Ethnicity  Income  Townsend deprivation index | Ethnicity   - White N = 385 357 (94.3%) - Asian N = 9731 (2.4%) - Black N = 7277 (1.8%) - Others N =6441 (1.6%)   Education   - Low N =65 293 (16%) - Intermediate N = 207 004 (51%) - High N = 133 902(33%)   Income   - < £18,000 N = 76 158 (21.7%) - 18,000–30,999 N = 89 104 (25.3%) - £31,000–51,999 N = 92 967 (26.4%) - ≥£52,000 N = 93 379 (26;6%)   Townsend deprivation quintile:   - 1 (least deprived) N = 82 356 (20.1%) - 2 N= 83 744 (20.1%) - 3 N = 82 668 (20.4%) - 4 N = 82 102 (20.2%) - 5 (most deprived) N = 76 158 (19.2%) | OR (95%CI)  Adjusted | 514 COVID-19 deaths (0.13%) |
| Bassett et al. 2020  (USA) | Cross-sectional Community  1 February 2020 to 22 July 2020 | 9 | Not specified | Completed  death certificates | Death counts from New York City | N = 306 800 000  Black 13.27%; Hispanic 0.85%; Asian 18.86% ; White 60.95% ; American/ Indian native 6.37% | Ethnicity | - White N = 186 400 000 - Black N = 40 600 000 - Hispanic N = 2 600 000 - Indian/Alaska N = 19 500 000 - Asian N = 57 700 000 | RR (95% CI)  Adjusted | 122500 deaths (0.04%) |
| McKeique et al.  2020  (UK) | Case-control  ICU admission/ control  Inclusion on June 14 2020 | 7 | NA | ICD-10 and BNF chapter with severe COVID-19 | ICU admission/ control  Severe cases = Mortality OR ICU admission  Severe COVID- | N = 41 220 | Scottish index of Multiple Deprivation  Care Home  Ethnicity | Scottish index of Multiple Deprivation: Indicator based on postal code:   - 1 N = 9680 - 2 N = 8 891 - 3 N = 7 556 - 4 N = 7 331 - 5 (Least deprived) N = 7 733   Care Home Residence N= 4 829  Ethnicity   - White - South Asian N = 172 - Black N = 40 - Other N = 162 | RR (95%CI)  UnAdjusted | 4 272 cases of severe COVID-19 |
| Mehta et al. 2021  (USA) | Cohort  Nursing home residents  April 1, 2020 - September 30, 2020 | 6 | NA | (ICD-10-CM) diagnosis code specific for COVID19 (U07.1) | Risk of diagnosis with SARS-CoV-2, hospitalisation or death within 30 days after diagnosis | N = 482 323  Mean age +/- SD 82.7+/-9.2;  Female 67.8%;  Black 12.6%;  Hispanic 5.1%;  Asian 2%;  White 79.6%;  Other 0.7% | Ethnicity | - Black N = 60 810 - Asian N = 9 819 - Hispanic or Latino N = 24 732 - Other N = 3 124 - White = 383 838 | HR (95%CI)  Adjusted | 131 119 COVID-19 infections (27.18%)  29 204 hospitalisations (6.05%)  26 384 deaths (5.47%) |
| Mendez-Dominquez et al. 2020  (Mexico) | Cross-sectional  Community  February 28, 2020 - April 21, 2020 | 7 | NA | RT-PCR | COVID-19 incidence, confirmation, hospitalisation and lethality (fatal cases) | N = 17 763  Mean age +/- SD 46.43 +/-0.55;  Female 55.31% | Migration  Indigenous ethnicity  Population affiliated to public health institutions  Urbanization  Interstate migrante | Migration. Refers to national and international migratory movements  Urbanization level: Percentage of the population living in communities with urban services and infrastructure. | IRR (95% CI)  OR (95% CI)  Adjusted | 9 501 COVID-19 cases (53.49%)  5 545 COVID-19 hospitalisations (31.22%) |
| Rostila et al. 2021  (Sweden) | Cohort  Community - Adult population aged 21+ living in Stockholm  January 31 2020 – May 4 2020 | 7 | 3 months | Death registry ICD-10 U07.1, U07.2, B3.42 | COVID-19 deaths  All-cause mortality excluding COVID-1 | N = 1778670  Female 49.55%;  ≥ High School Education 46.68%; Foreign born 31% | Country of birth  Income  Education  Employment status  Housing type  Working age in HH | Country of birth   - Sweden N = 1 232 511 - Other Nordic countries N = 58 206 - Europe N =165 988 - Middle East N = 127 125 - Africa N= 63 461 - Rest of the world N = 131 379   Income  Divided into quintiles   - 1 (least) N = 438 120 - 2 N = 437 733 - 3 N = 438 107 - 4 (most) N = 437 802 - NA N = 27 508   Education   - Primary N = 217 338 - Secondary N = 628 857 - Postsecondary N = 831 316 - Missing data N = 80 913   Employment status   - Employed N = 1 322 653 - Unemployed N = 456 617   Housing type   - House or apartment N = 1 167 041 - Special care N = 19 660   Working age in Household   - 0 people N = 284 878 - 1-2 people N = 1 099 933 - >=3 people N = 347 038 | RR (95% CI)  Adjusted | NA |
| Raisi-Estabragh et al. 2020  (UK) | Cohort  In-hospital  March 16 2020 – June 14 2020 | 7 | NA | RT-PCR | COVID-19 positive tests | N = 7099  Mean age (± SD) 69.11 (±8.65);  Minimum – Maximum age (40 – 69);  Female 50.3%;  White 91.5;  BAME (Black, Asian, and Minority ethnic) 7.9% | Ethnicity | Ethnicity   - White N = 6 498 (91.5%) - BAME N = 562 (7.9%) | OR (95% CI)  Both | 1 439 COVID-19 cases (20.27%) |
| Levin et al. 2021  (USA) | Cross-sectional  Emergency Department encounters  April 9 2020 – May 7 2020 | 8 | 1 month | RT-PCR | SARS-CoV-2 positive | N = 729  Median age 13;  Age range 2 -17;  Female 51%;  Black 10%;  Hispanic 34%;  White 46%;  Other 9%;  Not reported 1% | Ethnicity | Ethnicity - White N = 337; - Black or African American N = 73; - Other N = 64; - Hispanic N = 248 | OR (95% CI)  Adjusted | 81 infections (11.11%) |
| Shahbazi et al.  2020  (Iran) | Case-control  Hospitalised patients in Nahavand county  February 24 2020 – April 21 2020 | 7 | NA | RT-PCR | COVID-19 infection | N = 378  Female 64.55%; Single 30.2%; Married/Cohabiting 69.8%;  Living in urban areas 54.76 % | Place of residence  Marital status | Place of residence   - Urban N = 171 (45.24%) - Rural N = 207 (54.76%)   Marital status   - Single N = 114 (30.15%) - Married N = 264 (69.84%) | OR (95% CI)  Both | 126 COVID-19 cases (33.33%) |
| Aung et al.  2021  (USA) | Cross-sectional  World-wide mobile application-based cohort | 6 | Not specified | RT-PCR | Prevalent SARS-CoV-2 infection | N = 36041  Median age 42.98; Age range (35.01 – 53.95);  Female 65.27%;  Black 0.95%;  Hispanic 7.29%;  Asian 5.35%;  White 81.31%;  Other 2.28%  % Education ≥ High School 96.6% | Ethnicity  Education | Ethnicity:   - White N = 29 305 - Black N = 343 - Hispanic N = 2 628 - Asian/Pacific Islander N = 1 929 - Other/multi-racial N = 822   Education:   - Low Level - High level | OR (95% CI)  Adjusted | 484 infections (1.34%) |
| Williamson et al. 2020  (UK) | Cohort  General population registered with a general practice  February 1 2020 – May 6 2020 | 7 | NA | ONS death certificate data using the COVID related ICD-10 codes U071 or U072 | COVID-19 related deaths | N = 17 278 392  Female 50;1%;  Black 2%;  South Asian 5.9%;  White 62.9%;  Other 1.9%;  Mixed race 1%;  Missing 26.4% | Ethnicity  IMD quintile | Ethnicity   - White N = 10 866 411 (62.9%) - Mixed N = 169 697 (1%) - South Asian N= 1 022 130 (5.9%) - Black N = 339 909 (2%) - Other N= 320 132 (1.9%)   IMD quintile: measure of deprivation derived from patient’s postcode   - 1 (least deprived) N= 3 497 154 (20.2%) - 2 N= 3 476 668 (20.1%) - 3 N= 3 483 668 (20.2%) - 4 N= 3 480 459 (20.1%) - 5 N= 3 340 443 (19.3%) | HR (95% CI)  Adjusted | 10 926 COVID-19 deaths (0.06%) |
| Nafilyan et al.  2021  (UK) | Cohort  residents of England aged 65 years  March 2, 2020 - November 30, 2020 | 8 | NA | ICD codes | COVID-19 death | N = 10 078 568  Mean Age (± SD) 75.2 (7.6);  Female 53%;  White 93.9% | Living in household | - Single - Multi-generational household without children - Multi-generational household with children - 3+ elderly adults | HR (95% CI)  Adjusted | 39 419 (0.39%) deaths |
| Raisi-Estabragh et al.  2020  (UK) | Cross-sectional  General population  March 16 2020 until May 18 2020 | 8 | NA | COVID-19 tests results | COVID-19 infection | N = 4 510  Black 3.7%; Asian 3.1%; White 90.2%; Other 2.1% ; Mixed 0.7%; Chinese 0.2% | Ethnicity  Townsend deprivation score  Household size | NA | OR (95% CI)  Both | 1 326 COVID-19 infections (29.4%) |
| Rossi et al. 2020  (Italy) | Cross-sectional  General population in Northern Italy  March 3 2020 – March 26 2020 | 7 | NA | RT-PCR | COVID-19 infection | N = 2 635 tested persons  Female 47.06% | Place of birth | - Italians N = 2 420 (91.84%) - Immigrants N = 215 (8.16%) | OR (95% CI)  Adjusted | 1 564 COVID-19 infections (59.35%) |
| Zelner et al. 2021  (USA) | Cohort  General population  March 8 2020 until July 5 2020 | 7 | NA | RT-PCR | COVID-19 infection | N =49 701 | Race | Race   - Black N = 19 662 (39.56%) - Latino N = 3 657 (7.36%) - Other N = 1 612 (3.24%) - Asian/Pacific Islander N = 1 346 (2.71%) - White N = 23 301 (46.88%) - Native American N = 123 (0.25%) | IRR (95%CI)  Adjusted | 5 815 deaths (11.7%) |
| Ho et al. 2020  (UK) | Cohort  In-hospital setting  March 16 2020 – May 3 2020 | 7 | NA | RT-PCR | In-hospital COVID-19 incidence | N = 235 928  Age range 49-86 yeqrs old | Ethnicity  Deprivation score | Ethnicity   - White - Black - South Asian - Others | RR (p-value)  Both | 397 positive results conducted in hospital (0.17%) |
| Atkins et al. 2020  (UK) | Cohort  In-hospital setting  March 16 2020 – April 26 2020 | 7 | NA | RT-PCR | COVID-19 infection | N = 268 563  Mean age (± SD) 73.1(4.4); Black 1%; Asian 1.6%; White 96.2%; other 1.3%; Education > HS 78% | Ethnicity  Education | Ethnicity   - White N = 256 966 (95.68%) - Black N = 2 726 (1.02%) - South Asian N = 4 200 (1.56%) - Others N = 3 335 (1.24%)   Education   - None N = 57 794 (22%) - School/College N = 85 602 (32.6%) - Professional qualification N = 41 764 (15.9%) - Degree N = 77 558 (29.5%) | RR (95%CI)  Adjusted | 507 COVID-19 infections (0.19%) |
| Mutambudzi et al. 2021  (UK) | Cohort  In-hospital setting  March 16 2020 – July 26 2020 | 7 | NA | Positive test results | Severe COVID-19 | N = 120 075  Female 54.2%; Black 2.7%; Asian 2.6%; White 91.5%; Other 1.5%; Mixed 1% | Occupation  Socioeconomic deprivation  Ethnicity  Education | Occupation   - Non-essential workers N = 84 948 (70.7%) - Healthcare workers N = 10 748 (9%) - Social and education workers N = 13 476 (11.2%) - Other essential workers N = 10 903 (9.1%)   Socioeconomic deprivation   - Quartile 1 N = 28 488 (23.7%) - Quartile 2 N = 28 626 (23.8%) - Quartile 3 N = 31 802 (26.5%) - Quartile 4 (least advantaged) N = 31 159 (25.6%)   Ethnicity   - White British N = 102 485 (85.4%) - White Irish N = 3 205 (2.7%) - White Other N = 4 974 (4.1%) - Mixed N = 1 218 (1%) - South Asian N = 3 075 (2.6%) - Black N = 3 268 (2.7%) - Other N = 1 850 (1.5%)   Education   - College or University degree N = 48 189 (40.1%) - A levels/AS levels or equivalent N = 16 629 (13.8%) - O levels/GCSEs/CSEs or equivalent N = 39 730 (33.1%) - Other N = 10 157 (8.5%) - None of the above N = 5 370 (4.5%) | RR (95%CI)  Adjusted | 271 cases of severe COVID-19 (0.2%) |
| Mathur et al. 2021  (UK) | Cohort  Adults (aged ≥18 years) registered with primary care practices in England   - Wave 1: February 1 2020 – August 3 2020 | 7 | NA | ICD-10 codes U07.1 [confirmed COVID-19] or U07.2 [suspectedCOVID-19] anywhere on the death certificate | COVID-19 infection  COVID-19 hospitalisation  COVID-19 ICU admission  COVID-19 mortality | N = 17 288 532  Female 50%; Black 2&, Asian 5.9%; White 64.3%; Other 1.9%; Mixed 1% | Ethnicity | - White 64.3% - South Asian 5.9% - Black 2% - Mixed 1% - Other 1.9% - Unknown 25% | HR (95%CI)  Both | 71 246 COVID-19 infections (0.4%)  32 473 COVID-19 hospitalisations (0.2%)  3 096 ICU admissions (<0.1%)  11 469 COVID-19 deaths (0.1%) |
|  | - Wave 2: September 1 2020 – December 31 2020 | 7 | NA | ICD-10 codes U07.1 [confirmed COVID-19] or U07.2 [suspectedCOVID-19] anywhere on the death certificate | COVID-19 infection  COVID-19 hospitalisation  COVID-19 ICU admission  COVID-19 mortality | N = 17 636 366  Female 50%; Black 2&, Asian 6%; White 64.2%; Other 1.9%; Mixed 1% | Ethnicity | - White 64.2% - South Asian 6% - Black 2% - Mixed 1% - Other 1.9% - Unknown 24.8% | HR (95%CI)  Both | 506 773 COVID-19 infections (2.9%)  1 855 COVID-19 hospitalisations (<0.1%)  3 351 ICU admissions (<0.1%)  7 366 COVID-19 deaths (<0.1%) |
| **Ecological studies** | | | | | | | | | | |
| Akanbi et al.  2021  (USA) | Cross-sectional  Community- COVID-19 in 70 zip codes in Oakland County  April 3 2020 - May 16 2020 | 7 | 1.5 months | NA | Crude and adjusted incidence of COVID-19 and COVID-19 case fatality rates | 70 Zip codes  Mean age (± SD) 41.41 (±4.24);  Median age 42.14;  Age range 37.92 – 44.10;  Black 13.6%;  Asian 5.6%;  White 77.3%;  Other 3.5% | Ethnicity  Education level  Median family income  Poverty level | Ethnicity: % of Black people per ZIP code  Education level: have a bachelor degree  Median family income: in USA dollars  Poverty level: below poverty | IRR (95% CI)  Both | NA |
| Adhikari et al. 2020  (USA) | Cross-sectional  Community - Inclusion158 counties that experienced at least one death attributed to confirmed COVID-19 as of May 10 2020 | 7 | NA | COVID-19 infection and deaths provided by Centers for Disease Control and Prevention, state health departments and aggregated by USA Facts | Cumulative COVID-19 incident infections and deaths | 158 counties | Ethnicity | Substantially White : 3-17.9%; Less diverse : 18 -29.4%; More diverse : 29.5-44.5%; Substantially non White : >44.5% | RR (95% CI)  Adjusted | NA |
| Alves et al.  2021  (Portugal) | Cross-sectional  Community - Portuguese municipalities  Inclusionat four different points:  - April 1 2020  - May 1 2020  - June 1 2020  - July 1 2020 | 8 | NA | NA  (data from the Portuguese Directorate-General of Health (DGS)) | Cumulative number of COVID-19 cases per municipality | 308 municipalities | Unemployment  Earning  Gini coefficient | Unemployment: Number of persons registered as unemployed at the Portuguese public employment service divided by the number of residents (25–64 years old). It corresponds to three levels: 1st percentile – 2nd percentile – 3rd percentile  Earning : Average monthly earnings by municipality (e) divided in tertiles (1st , 2nd and 3rd)  Gini coefficient: Gini coefficient for the declared gross income, subtracted from the income tax. It corresponds to three terciles (1st, 2nd, 3rd)) | OR (95% CI)  Both | NA |
| Li et al. 2020  (USA) | Cross-sectional  County  April 14 2020 | 7 | NA | NA  Data from Center for Systems Science and Engineering (CSSE) Coronavirus Resource Center at Johns Hopkins University | COVID-19 cases/100,000 people  COVID-19 deaths/100,000 people | 661 counties | Race | - NH White - Black - Hispanic - Asian - Native Hawaiian/Other Pacific native   American Indian/Alaska native | OR (95% CI)  Regression coefficients  Both | NA |
| Temkin Greener et al. 2020  (USA) | Cross-sectional  Residents of Assisted Living (AL) communities  March 2020 - May 29 2020 | 7 | NA | Laboratory confirmed cases | ASL COVID-19 cases and deaths | N = 3994 ALs  Mean age (± SD) 76 (± 11.1);  Female 74.3%;  Black and Hispanic 19.9% | Ethnicity | - Black and Hispanic N=795 - White | OR (95% CI)  Adjusted | 2 542 cases (96.0%) and 675 deaths (86.9%) |
| Abedi et al.  2020  (USA) | Cross-sectional  Community-based –  County level  April 9 2020 | 8 | NA | Laboratory testing samples | COVID-19 infection and mortality rates | N = 102 178 117 | Ethnicity  Median income  High school  Bachelor degree  Poverty | Ethnicity   - Black male: Percent BA (Black Alone) MALE - White male: Percent NHWA (non-Hispanic White Alone) MALE - Asian male: Percent AA (Asian Alone) MALE - Hispanic male: Percent H (Hispanic) MALE - Black female: Percent BA (Black Alone) FEMALE - White female: Percent NHWA (non-Hispanic White Alone) FEMALE - Asian female: Percent AA (Asian Alone) FEMALE - Hispanic female: Percent H (Hispanic) FEMALE   Median income: Median Family income; past 12 months 2018  High school  Bachelor degree: Percent Population 25 years and over Bachelor’s degree or higher  Insurance: Percent Public Coverage Medicaid means tested public coverage alone or in combination   - Poverty: Percent below poverty level Population for whom poverty status is determined | Regression coefficient  Adjusted | COVID-19 infection rate per one million mean (± SD), 912.20 ± 1034.26  COVID-19-related death (mean, 4.13%± 2.70%; median, 3.40; IQR, 2.22–5.61) |
| Biggs et al.  2020  (USA) | Cross- sectional  Louisiana census tracts.  March 9 2020 – August 24 2020 | 8 | NA | Clinical specimen Usinga molecular amplification detection | COVID-19 cumulative incidence | N = 1105 census tracts  Percentage No High School Diploma 16% (SD=8.8);  Percentage Living below poverty line 21.4%(SD=12.8);  Percentage Minority (all persons except non-Hispanic Whites) 45.1%(SD=28.9);  Percentage of single parent households 11.1% (SD = 6.4) | Overall Social Vulnerability Index  Theme 1 Socioeconomic  Theme 2-House Composition & Disability  Theme 3- Minority & Language  Theme 4- Housing & Transportation | Theme 1 Socioeconomic is based on 4 elements:   - Percentage Living Below Poverty Line - Unemployment Rate - Per Capita Income - Percentage with No High School Diploma   Theme 2-House Composition & Disability is based on 4 elements:   - Percentage of persons aged 65 and older - Percentage of persons aged 17 and younger - Percentage with a disability - Percentage of single parent households   Theme 3- Minority & Language is based on 2 elements:   - Percentage Minority (all persons except non-Hispanic Whites) - Percentage of persons who Speak English “Less than Well”   Theme 4- Housing &Transportation is based on 5 elements:   - Percentage of housing in structures with 10 or more units - Percentage of mobile homes - Percentage of occupied housing units with more people than rooms - Percentage of households with no vehicle available - Percentage of persons in institutionalized group quarters | RR (95% CI)  Both | NA |
| Daras et al. 2021  (UK) | Cross-sectional  Residents of 6789 Middle Super Output Areas (MSOAs)  March 1 2020 – May 31 2020 | 8 | NA | Death certificates and COVID-19 reported as underlying cause of death Usingcodes: ICD codes: U07.1 and U07.2 | COVID-19 mortality | 6789 Middle Super Output Areas (MSOAs) (average 8242 people) | Overcrowding (%)  Black, Asian and Minority Ethnic groups (%) | Overcrowding (%):  proportion of overcrowded households in each MSOA based on the number of bedrooms in the household | IRR (95% CI)  Adjusted | NA |
| Chen et al. 2020  (USA) | Cross-sectional  USA county  As of May 5 2020 | 8 | NA | COVID-19 deaths at the county level from USA Facts | COVID-19 mortality rates  Confirmed cases per 100 000  Positive tests per 100 000 | 3142 counties (322 903 3030 population) | % poverty (categories)  Index of Concentration at the Extremes (high income White households versus low income Black households)  % crowding (quintiles)  % percent population of color | % poverty (categories)   - 0-4.9%   N =4 495 932   - 5-9.9%   N =71 157 744   - 10-14.9%   N = 108 820 591   - 15-19.9%   N = 101 961 251   - 20-100%   N = 36 428 205  Index of Concentration at the Extremes (high income White households versus low income Black households)   - Q1: (-0.522,0.114)   N = 61 949 063   - Q2: (0.114,0.159)   N = 64 942 197   - Q3: (0.159,0.205)   N = 65 113 354   - Q4: (0.205,0.283)   N = 64 525 801   - Q5: (0.283,0.536)   N = 66 333 308  % crowding (quintiles)   - Q1: [0%, 1.47%)   N = 65 389 574   - Q2: (1.47%, 2.12%)   N = 64 425 866   - Q3: (2.12%, 3.06%)   N = 63 510 499   - Q4: (3.06%, 4.91%)   N = 65 654 959   - Q5: (4.91%, 49.3%)   N = 63 913 934  % percent population of color   - Q1: (0%, 17.2%)   N = 65 219 939   - Q2 (17.2%, 30.2%)   N = 65 166 967   - Q3(30.2%, 44.3%)   N = 69 376 152   - Q4(44.3%, 61%)   N = 60 922 155   - Q5(61%, 100%)   N = 62 217 817 | RR (95% CI)  Adjusted | 68 656 deaths  1 189 798 cases |
| De Souza et al.  2020  (Brazil) | Cross-sectional  Residents of 2496 Municipalities  Inclusion until May 6 2020 | 7 | NA | NA  Data on cases and deaths were obtained from the  CoVida network panel | COVID-19 incidence rate/100,000 inhabitants  COVID-19 mortality rate/100,000 inhabitants | N=2496 municipalities (179 084 725 inhabitants) | Social vulnerability index (SVI):synthetic indicator based on urban infrastructure, human capital, income and work  Municipal human development index (MDHI): synthetic indicator based on longevity, education, and income  Population size: number of inhabitants per municipalities | MHDI   - Very low (0-0.499) N=18 municipalities 0.72% (489795 inhabitants) - Low (0.500-0.599) N=588 municipalities 23.56% (13623241 inhabitants) - Medium (0.600-0.699) N=830 municipalities 33.25% (31335083 inhabitants) - High (0.700-0.799) N=1016 municipalities 40.71% (100556389 inhabitants) - Very high (0.800-1) N=44 municipalities 1.76% (33080217 inhabitants)   SVI   - Very low (0-0.200) N=350 municipalities 14.02% (20782329 inhabitants) - Low (0.200-0.300) N=703 municipalities 28.17% (79129666 inhabitants) - Medium (0.300-0.400) N=485 municipalities 19.43% (49839712 inhabitants) - High (0.400-0.500) N=534 municipalities 21.39% (17423869 inhabitants) - Very high (0.500-1) N=424 municipalities 16.99% (11909149 inhabitants)   Population size   - ≤10,000 inhabitants N=482 municipalities (2824212 inhabitants) 19.31% - 10,001 to 20,000 N=580 municipalities 23.24% (8549277 inhabitants) - 20,001 to 50,000 31.37% N=783 municipalities (24491464 inhabitants) - 50,001 to 100,000 N=328 municipalities 13.14% (22642923 inhabitants)   >100,000 N=323 12.94% (120576849 inhabitants) | Correlation coefficient (95% CI)  Both | 125186 COVID-19 cases (0.07%)  8452 COVID-19 deaths (0.005%) |
| Demenech et al.  2020  (Brazil) | Cross-sectional  Brazil federative units (FU)s  Inclusion on twelve different days between April 27 2020 and July | 5 | NA | NA  Data extracted from Panel Coronavírus portal4 | COVID-19 incidence rates  COVID-19 mortality rates | N=27 Federative Units | Gini coefficient  Population density | Gini coefficient: measure for estimating the degree of income concentration of the population of each FU. It can vary between 0 and 1, and the higher the value, the greater the concentration of income  Population density (people/km^2^):  Measure calculated by dividing the total population and the territorial area (in km2) of each FU | Correlation coefficient (95% CI)  Adjusted | NA |
| Di Girolamo et al. 2020  (Italy) | Cross-sectional  Residents in Emilia-Romagna  1 March 2020-30 April 2020 | 7 | 1 March 2020-30 April 2020 | NA  Regional mortality suveillance | Counts of all deaths  Counts of deaths attributable to COVID-19 | N=44 27 364 | Index of deprivation: measure of social  and material deprivation based on five census variables (low level of education, unemployment, non-home ownership,  single-parent family, and household crowding)  Household crowding: number of people per 100 square metres of  the House surface  Proportion of foreign resident population: the percentage of the foreign resident population: a  proxy for social and economic disadvantage | Index of deprivation   - Q1 N=891 734 (20.14%) - Q2 N=857 536 (19.37%) - Q3 N=860 557 (19.44%) - Q4 N=872 345 (19.70%) - Q5 N=927538 (20.95%) - Missing N=17654 (0.40%)   Household crowding   - Q1 N=923 539 (20.86%) - Q2 N=869 010 (19.63%) - Q3 N=86 9685 (19.64%) - Q4 N=867693 (19.60%) - Q5 N=884687 (19.98%) - Missing N=12750 (0.29%)   Proportion of foreign resident population   - Q1 N=893029 (20.17%) - Q2 N=855531 (19.32%) - Q3 N=853270 (19.27%) - Q4 N=877973 (19.83%) - Q5 N=939516 (21.22%)   Missing N=8045 (0.18%) | Mortality rate ratio (95%CI)  Adjusted | 3 301 COVID-19 deaths (0.07%) |
| Rodriguez-Villamizar et al.  2021  (Columbia) | Cross-sectional  Colombian municipalities  Inclusion up to July 17 2020 | 7 | NA | RT-PCR | COVID-19 mortality | 772 municipalities | % Urban population  Population density  Poverty index | Socioeconomic determinant: Mean (SD)  % Urban population: 49.26 (24.71)  Population density: 229.98 (899.66)  Poverty index: socioeconomic ecologic measure at the municipality level that ranges from 0 to 100 with higher percentages meaning privation of more indicators and dimensions. 39.85 (17.73) | OR (95% CI)  Both | 182 140 COVID-19 cases  6 288 COVID-19 deaths |
| Sugg et al. 2021  (USA) | Cross-sectional  Nursing homes  Inclusiontill June 30 2020 | 6 | NA | NA  Data on COVID-19 infection data for staffs and residents were collected from the Centers for Medicare and Medicaid Services (CMS) | Cumulative COVID-19 nursing home cases | 13709 nursing homes | % Total population: Asian  % Total population: American Indian  % Total population: African American  Average household size  Per capita income  % Civilian population in labor force 16 years and over:  unemployed  Population per sq. mile | NA | RR (95% CI)  Adjusted | Average of 11.32 confirmed cases per nursing home |
| Yang et al. 2021  (Hong Kong) | Cross-sectional  Tertiary Planning Unit Level  Wave 1&2:  January 18 2020 – April 30 2020  Wave 3:  May 1 2020 – August 31 2020 | 7 | NA | Laboratory confirmed COVID-19 cases | COVID-19 incidence | Wave 1&2:  N = 1038  Wave 3:  N = 3773 | Tertiary education or above (per 10% increase)  Executives and professionals (per 10% increase)  Median monthly household income (per HKD 10,000 increase) | Tertiary education or above (per 10% increase) represents the proportion of people with tertiary education such as the number of residents >14 years who had attained at least a tertiary-level education divided by the total number of residents >14 years in a given TPU. Education is divided into three levels: primary, secondary and tertiary  Executives and professionals (per 10% increase): refers to the number of residents who were employed as managers and administrators, professionals, or associate professionals divided by the total number of employed residents in a given TPU  Median monthly rent (per HKD 10 000 increase) | IRR (95% CI)  Adjusted | 4811 COVID-19 cases  Wave 1&2:  N = 1038 (14%)  Wave 3:  N = 3773 (86%) |
| Figueroa et al. 2020  (USA) | Cross-sectional  Massachusetts cities  January 1 2020 – May 6 2020 | 8 | NA | NA  Confirmed cases available on The Massachusetts Department of Public Health database | COVID-19 cases per 100 000 population | 351 towns | Ethnicity  Median household income  Average household size  Proportion with < HS education  Proportion of non citizens | Ethnicity - Hispanic or Latino; - Black non Latino; - White non Latino; - Other non Latino | Regression coefficients (95% CI)  Both | NA |
| Figueroa et al.  2021  (USA) | Cross-sectional  USA counties  January 1 2020 – September 12 2020 | 8 | NA | NA  Confirmed COVID-19 cases and deaths from the the Johns Hopkins Coronavirus Resource Center data repository and USA facts | COVID-19 cases and deaths per 100,000 | NA | Ethnicity  Median household income  Average household size  Proportion with < HS education  Proportion of non citizens | Ethnicity - Hispanic or Latino; - Black non Latino; - White non Latino; - Other non Latino | Regression coefficients (95% CI)  Both | NA |
| Garcias et al.  2021  (Spain) | Cross-sectional  17 Spanish regions  Inclusion as of May 23 2020 | 8 | NA | NA  Data on COVID-19 mortality and incidence from the Spanish government | Mortality rate of COVID-19 per 1,000, 000 inhabitants  Estimated  cumulative incidence | 17 regions | Gini Index  Urban population  Log GDP per capita | Gini Index is used to measure income inequality within each region: Mean (± SD) = 0.309 (± 0.25)  Proportion of urban people refers to the proportion of population living in municipalities with more than 100,000 inhabitants: Mean (± SD) = 36.85 (± 14.65)  Log GDP per capita is the logarithm of GDP per capita: Mean (± SD) = 10.12 (± 0.19) | Regression coefficients (95% CI)  Adjusted | NA |
| Hawkins D. et al.  2020  USA | Cross-sectional study  Residents of Massachusetts (MA)  January 1 2020- June 10 2020 | 8 | 5 months | Laboratory-confirmed cases of COVID-19 | Cases per 100 000 residents  Tests per 100 000 residents | N = 87 256 | Percentage of residents in poverty  Median income  Percentage of residents who rented  Percentage of residents who were uninsured  Unemployment rate | Percentage of residents in poverty   - First quartile N = 6985 (8.0) - Second quartile N = 8819 (10.1) - Third quartile N = 16 338 (18.7) - Fourth quartile N = 55 114 (63.2)   Median income   - Fourth quartile N = 8024 (9.2%) - Third quartile N = 15 709 (18%) - Second quartile N = 28 530 (32.7%) - First quartile N = 34 993 (40.1%)   Percentage of residents who rented   - First quartile N = 6533 (7.5%) - Second quartile N = 9603 (11%) - Third quartile N = 18 569 (21.3%) - Fourth quartile N = 52 551 (60.2%)   Percentage of residents who were uninsured   - First quartile N = 5402 (6.2%) - Second quartile N = 13 129 (15%) - Third quartile N = 22 907 (26.3%) - Fourth quartile N = 45 818 (52.5%)   Unemployment rate   - First quartile N = 7848 (9%) - Second quartile N = 14 471 (16.6%) - Third quartile N = 22 661 (26%) - Fourth quartile N = 42 276 (48.5%) | RR (95% CI)  Adjusted | 87 256 COVID-19 cases  1 584.8 COVID-19 cases per 100 000 |
| Li et al. 2020  (USA) | Cross-sectional  Nursing homes nationally  May 25 2020 – May 31 2020 | 7 | NA | RT-PCR | Numbers of incident COVID-19 confirmed cases among residents  Staff  Incident COVID-19 related deaths among residents  Confirmed new cases among staff | N = 12 576  ≥ High School 87.5%;  Medium household income 1000 $: 56,4 % | Proportions of racial/ethnic minority residents | - Low proportion group (<2.92%) N = 3 143 - Medium-proportion group (2.92% – 11.11%) N = 3 149 - Medium-high–proportion group (11.11%–30.18%) N = 3 140   High-proportion group (≥30.18%) N = 3 144 | OR (95%CI)  Adjusted | 0.9 ± 4.7 cOVID-19 confirmed cases among residents  0.3 ± 1.5 COVID-19 related deaths among residents |
| Kolin et al.  2020  (UK) | Cohort  Census zone  March 16 2020 – May 18 2020 | 7 | NA | RT-PCR | Covid-19 positive status | N = 397 064  Mean age (± SD) 56.4 (±8.1);  Minimum – Maximum age 40 – 69;  Female 54.8;  Black 1.9%;  Asian 2.6%;  White 94.2%; | Economic activity  Ethnic group  Household composition  Living arrangements  Qualifications  Tenure | Economic activity   - Individuals self employed - Individuals unemployed - Individuals retired   Ethnic group   - White Individuals - Multiple Ethnicities Individuals - Black Individuals   Household composition   - Individuals Lone Parents - Individuals in Other Household Type with Dependent Children   Living arrangements   - Individuals Living Not in Couple - Individuals Separated but Married   Qualifications   - Individuals with No Qualification - Individuals with Level 2 Qualification (i.e. School certificate, intermediate higher diploma …) - Individuals with Level 4 Qualification (i.e. degree, professional qualifications etc…)   Tenure   - Individuals Own Home - Individuals Socially Rent - Individuals Rent from Other | RR (95% CI)  Adjusted | 968 COVID-19 cases (0;24%) |
| Loomba et al. 2021  (USA) | Cross-sectional  Community State level  March 14, 2020 - April 30, 2020 | 6 | NA | NA | COVID-19 cases and mortality | 3332 COVID-19 cases per 1 000 000 patients | Race  Insurance  Average household income  High school grad percent  Per capita health care spending  Public transportation volume  State population density | Race   - White - Black - Native American - Asian - Islander - Other race - Multiple race   Insurance  Insured / uninsured | Regression Coefficient (95% CI)  Both | 1 090 500 COVID-19 cases  63 642 CVOID-19 deaths |
| Hawkins R. et al. 2020  (USA) | Cross-sectional study  USA countries  May 2, 2020 | 8 | X | X | COVID-19 Infection  COVID-19 Mortality | 3127 counties | - Housing vacancy rate - Median income ratio - Poverty rate - % change in employment - % of adults not working - % of adults with high school degree - % population in distressed zip codes - % of population Black   % of uninsured population aged younger than 65 | - Housing vacancy rate - Median income ratio - Poverty rate - % change in employment - % of adults not working - % of adults without high school degree - % population in distressed zipcodes - % of population Black   % of uninsured population aged younger than 65 | RR(95% CI)  Adjusted | 1 089 999 cases and 62 298 deaths in 3127 counties  Case fatality rate 5.7% |
| Karmakar et al.  2021  (USA) | Cross-sectional  USA counties  20 January 2020- July 29 2020 | 7 |  | NA | Infection  Mortality | 3137 counties | Poverty rate  Unemployment rate (%)  Per capita income  Education  Adults without health insurance (%)  Gini income inequality index   - racial/ethnic minorities and language, % | Percentage minority (all persons except White, non-Hispanic) = Any racial/ethnic minority:   - African american - Aispanic or Latinx - American Indian or Alskan native - Asian   Education = Education, % age ≥25 y with no high school degree | IRR (95% CI)  Adjusted | 289 283 COVID-19 cases  147074 COVID-19 deaths |
| Khazanchi et al.  2020  (USA) | Cross-sectional  USA counties  April 19 2020 | 8 | NA | NA  COVID-19 case and death rates compiled by The New York Times from health agency reports | Mortality | 2754 counties | - Social vulnerability index - Socioeconomic status - Household composition and disability - Minority status and language   Housing type and transportation | NA | IRR (95% CI)  Adjusted | 25,978 COVID-19 deaths across 2754 counties |
| Gorges et al. 2021  (USA) | Cross-sectional  Residents of nursing homes  January 1 2020 – September 13 2020 | 7 | NA | Laboratory test  Specific signs or symptoms or patient-specific transmission-based precautions for  COVID-19 infection | COVID-19 deaths among nursing home residents | N = 13 312  Mean age +/- SD 79.5 +/- 6.7;  White 77.9% | Race | Nursing home racial composition measured as the proportion of residents of non-Hispanic White race contained in the database and grouped into 5 racial composition quintiles across the sample.   - 1 : 0-59.73 - 2 :59.75 - 80.99 - 3 : 81-91.77 - 4 : 91.78-97.32   5 : 97.33-100 | RR (95% CI)  Both | 51 606COVID-19–associated deaths  Mean (SD) of 3.9 deaths per facility |
| Gaudart et al.  2021  (France) | Cross-sectional  96 departments / in-hospital patients  March 19 2020 - May 11 2020 | 7 | 2 months | PCR / CT scan | In-hospital COVID-19 incidence  COVID-19 mortality  COVID-19 fatality | 96 adminstrative departments | Economic indicators | - High median standard of living - High rate of social assistance   High poverty and unemployment ratio | SIR=standardised incidence ratio (IC 95%)  UnAdjusted | 16 597 ICU admission  17 062 deaths |
| Khanijahani et al. 2021  (USA) | Cohort  USA counties  Jan 22 2020 - 21 July 2020 | 7 | 6 months | NA  Data from the Center for Systems Science and Engineering (CSSE) at Johns Hopkins University | Confirmed COVID-19 deaths per 100,000 | 73056 tracts /3142 counties | % residing in concentrated disadvantaged (10% increments)  % residing in Black concentration (10% increments) | - % residing in Black concentration (10% increments) = Black-concentrated neighborhoods if25% or more ofthe residents were Black | OR(95% CI)  Adjusted | Average of 20.4 per 100,000 population COVID-19 death |
| Macchia et al.  2021  (Argentina) | Cohort  Community  January 31 – July 7 2020 | 6 | NA | RT-PCR | Incidence rate of COVID-19 | N = 114 052  Female 50.11% | Socioeconomic status (SES) quintiles | It is a score that includes the average income of a family, the type of employment and economic activity, the years of education (both as a categorical or continuous variable), the type of medical insurance (public and private) and the housing conditions. On the basis, of these domains a score was created which was then divided quintiles:   - 1 - 2 - 3 - 4 - 5 - Slums | IRR (95% CI)  UnAdjusted | 39 039 COVID-19 cases |
| Palacio et al. 2021  (USA) | Cross-sectional  Community  Inclusionup to July 24 2020 | 7 | NA | NA | COVID-19 cases | N = 2 834 352 | Quartile income  Tertile of Self-reported Financial Strain | - Quartile income 50194-61897$ - Quartile income 41541-49239$ - Quartile income 14999-40039$ - Tertile of Self-reported Financial Strain (34- 52$)   Tertile of Selfreported Financial Strain(53-100$) | IRR (95%CI)  Adjusted | 97 594 COVID-19 infections |
| Lewis et al. 2020  (USA) | Cross-sectional  General population (community  July 3 2020 – July 9 2020 | 7 | NA | RT-PCR | COVID-19 incidence | 99 Utah small statistical areas | Level of deprivation | - Very low (least deprived) - Low - Average - High - Very high (most deprived) | OR (95%CI)  Both | 28 139 COVID-19 infections |
| Das et al. 2020  (India) | Cross-sectional  General population  May 15 2020 – May 21 2020 | 5 | NA | NA | COVID-19 infection | 155 Electoral wards | - Poor Housing Condition - Lack of Household Amenities and Services   Low Asset Possession |  | Regression coefficients (p-value)  Adjusted |  |
| Li et al. 2020  (USA) | Cros—sectional  Residents of nursing homes (NHs)  Inclusionas of April 4 2020 | 7 | NA | Laboratory confirmed cases | COVID-19 infection  COVID-19 mortality | 215 Nursing homes with at least once case | NHs concentrated by Medicaid residents   - NHs concentrated by racial/ethnic minority residents | NHs concentrated by Medicaid residents defined as NHs in the top quartile group for percentage of Medicaid residents (i.e., ≥78.1%) versus all other NHs in Connecticut.   - NHs concentrated by racial/ethnic minority residents defined as nursing homes in the top quartile group for percentage of racial and ethnic minority residents (i.e., ≥21.7%) versus all other nursing homes in Connecticut. | OR (95%CI)  Adjusted | Mean (range)  1 918.8 infections (87–6,816)  121.1 deaths (1-406) |
| Alipio et al. 2020  (Philippine) | Cross-sectional  General population  Inclusionas of April 7 2020 | 6 | NA | NA | COVID-19 infection | 17 regions | Income | NA | Regression coefficients (p-value  Adjusted | NA |
| Ramirez-Aldana et al. 2020  (Iran) | Cross-sectional  General population  February 419 2020-March 18 2020 | 6 | NA | NA | COVID-19 infection | 31 provinces | Urban population  Literacy | NA | Regression coefficients (p-value)  Adjusted | 100 988 COVID-19 cases  16 597 ICU admissions  17 062 deaths |
| Ginsburgh et al. 2021  (France) | Cross-sectional  Hospital setting  May 13 2020 – September 3 2020 | 6 | NA | NA | COVID-19 infection per 100 000  COVID-19 mortality per 100 000 | 94 French continental departments | Income  Household size  Rural dummy  Gini index | NA | Regression coefficients (p-value)  Adjusted | Mean (range)  COVID-19 infections 1.78.79 per 100 000 (43.03-705.80 per 100 000)  COVID-19 deaths 22.16 per 100 00 (0.66-114.19 per 100 000) |
| Mollalo et al. 2020  (USA) | Cross-sectional  General population / County-level  January 22 2020 – April 9 2020 | 8 | NA | NA | COVID-19 infection | NA | Income inequality  Median household inequality  % of Black females | NA | Regression coefficients (p-value)  Adjusted | NA |
| Ehlert et al. 2020  (Germany) | Cross-sectional  General population  Inclusionuntil June 16 2020 | 10 | NA | NA | COVID-19 infection  COVID-19 mortality | 401 administrative districts | Household income  Unemployment rate  Rural regions  Academics | NA | Regression coefficients (p-value)  Adjusted | COVID-19 infections  COVID-19 deaths |
| Gross et al. 2020  (USA) | Cross-sectional  General population  Inclusionas of April 21 2020 | 7 | NA | NA | COVID-19 mortality | 28 states | Race  Ethnicity | Race   - White - Black   Ethnicity   - White   Latinx | SMR Standardised mortality ratio (95%CI)  UnAdjusted | COVID-19 deaths |
| Karaye et al. 2020  (USA) | Cross-sectional  General population  Inclusionas of May 12 2020 | 8 | NA | NA | COVID-19 infection | 48 states | Overall SVI  Socioeconmic status (SES)  Household composition  and disability  Minority status and language  Housing and transportation | NA | Regression coefficients (p-value)  Adjusted | 1 320 909 COVID-19 infections |
| Khanijahani et al. 2020  (USA) | Cross-sectional  General population  Inclusion until November 2 2020 | 8 | NA | NA | COVID-19 infection  COVID-19 mortality | 3142 USA counties  Mean (SD)  % Black 9.9 (14.7) ;  % Hispanic 9.3 (13.8) ;  % Female 49.9 (2.4) ;  Median age 41.3 (5.4) | % Hispanic  % Black  Median household income  Average household size  Unemployment rate | NA | Regression coefficients (95%CI)  Adjusted | NA |
| Fielding-Miller et al. 2020  (USA) | Cross-sectional  General population  Inclusionas of July 12 2020 | 6 | NA | NA | COVID-19 mortality | 3 024 counties | % Residents uninsured  % Residents in poverty | NA | Regression coefficients (p-value)  Adjusted | NA |
| White et al. 2020  (USA) | Cross-sectional  Nursing facilities  Inclusionas of April 21 2020 | 7 | NA | Laboratory‐confirmed SARS‐CoV‐2 | COVID-19 infection | 341 nursing facilities | % Black | NA | Marginal effect (95%CI)  Adjusted | NA |
| Zhang et al. 2020  (USA) | Cross-sectional  General population  Inclusionas of May 1 2020 | 8 | NA | NA | COVID-19 infection | 1 624 counties | % Population below poverty  % Minority population  % Uninsured population | NA | Regression coefficients (p-value)  Adjusted | NA |

##### Table 2. Association for socioeconomic determinants and outcomes among population representative samples – Etiological role

| **First author (Country)** | **Sample** | | **Outcome** | **N (%) of people for which the outcome occurred (total and per socioeconomic determinants level where available)** | **Association with socioeconomic determinants - Unadjusted** | **Association with socioeconomic determinants - Adjusted** | **Adjustment factors** |
| --- | --- | --- | --- | --- | --- | --- | --- |
| **Individual-level studies** | | | | | | | |
| Bailey et al.  2020  (USA) | N = 135 794  Patients were younger than 25 years prior to March 1, 2020 from 7 children’s health systems | | SARS-CoV-2 Infection | 4814 infections(3.55%) - White N= 2085 (43.31%) - Black N= 1543 (32.05%) - Hispanic N= 1026 (21.31%) - Asian or Pacific Islander N= 160 (3.32%) \| | NA | Socioeconomic determinants = OR (95% CI)  Race / ethnicity   - White - Black 2.66 (2.43 – 2.90) - Hispanic 3.75 (3.39 – 4.15) - Asian or Pacific Islander 2.04 (1.69 – 2.48) | age, chronic conditions, health care Use, testing location, payer (commercial/public) |
| Levin et al. 2021  (USA) | N = 729  Emergency Department encounters  April 9 2020 – May 7 2020 | | SARS-CoV-2 Infection | 81 (11.11%)   - White N = 15 (18.52%) - Black or African American N = 8 (9.88%) - Other N = 3 (0.037%) - Hispanic N =55 (67.9%) | NA | Socioeconomic determinants = OR (95% CI)  Race / ethnicity   - White = Reference - Black or African American = 2 (0.8-4.9) - Hispanic 4.9 (2.6-8.9) | Age, presenting symptoms |
| Bergman et al. 2021  (Sweden) | N = 518 739  Community  January 1 2020 – September 27 2020 | | SARS-CoV-2 Infection | 68 575 infections (13.22%)  Country of Birth   - Sweden N = 51 281 (74.8%)   Education   - Primary N = 11 837 (18.2%) - Secondary N = 27 705 (42.7%) - Post-secondary < 3y   N = 9 011 (13.9%)   - Post-secondary> 3y   N = 16 313(25.1%)  Disposable family income in 2018   - Quintile 1 N = 12 120 (18.3%) - Quintile 2 N = 12 407 (18.7%) - Quintile 3 N = 11 500 (17.3%) - Quintile 4 N = 14 386 (21.7%) - Quintile 5 N = 16 313 (25.1%) | Socioeconomic determinants = OR (95% CI)  Country of Birth   - Sweden 0.72 (0.71 - 0.74) - Not born in Sweden = Reference   Education   - Primary = Reference - Secondary = 1.08 (1.05-1.1) - Post-secondary < 3y = 1.06 (1.03-1.09) - Post-secondary> 3y = 1.23(1.2-1.26)   Disposable family income in 2018   - Quintile 1 = Reference - Quintile 2 = 1.02 (0.99-1.05) - Quintile 3 = 0.94 (0.92-0.97) - Quintile 4 = 1.18(1.15-1.21) - Quintile 5 = 1.3 (1.27-1.34) | Socioeconomic determinants = OR (95% CI)  Country of Birth   - Sweden = 0.71 (0.69-0.72) - Not born in Sweden = Reference   Education   - Primary = Reference - Secondary = 1.11 (1.08 -1.15) - Post-secondary < 3y = 1.09 (1.05-1.13) - Post-secondary> 3y = 1.19 (1.15 – 1.23)   Disposable family income in 2018   - Quintile 1 = Reference - Quintile 2 = 1.3 (1.26 - 1.35) - Quintile 3 = 1.74 (1.69 -1.8) - Quintile 4 = 1.88 (1.82 -1.95) - Quintile 5 = 1.81 (1.75-1.88) | Sex, age, education, family disposable income, Stockholm residence, long-term care facility, country of birth |
| Chadeau-Hyam et al. 2020  (UK) | N = 4 509  Community  March 16 2020 – May 18 2020 | | SARS-CoV-2 Infection | 1325 COVID-19 cases (32.92%) | Socioeconomic determinants = OR (95% CI)  Ethnicity   - White = Reference - Black = 2.14 (1.57-2.93) - Other 1.68 (1.29-2.18)   Education   - High = Reference - Intermediate = 1.25 (1.07-1.46) - Low = 1.40 (1.16-1.68)   Type of accommodation   - House = Reference - Flat = 1.02 (0.85-1.24)   Own or rent accommodation   - Own outright = Reference - Own with a mortgage = 1.27 (1.09-1.46) - Rent = 1.32 (1.10-1.58)   Number in household 1.08 (1.03-1.15)  Number in household  Income   - less than 18 000 = 1.06 (0.87-1.28) - 18 000 to 30999 = Reference - 31 000 to 51 999 1.01 (0.82-1.25) - greater than 52 000 0.88 (0.71-1.09)   Occupation   - Employed (other) = Reference - Healthcare worker = 1.22 (1.00-1.49) - Unemployed = 1.05 (0.87-1.27) - Retired = 0.95 (0.81-1.11) | Socioeconomic determinants = OR (95% CI)  Ethnicity   - White = Reference - Black = 2.14 (1.57-2.93) - Other 1.68 (1.29-2.18)   Education   - High = Reference - Intermediate = 1.15 (1.05-1.26) - Low = 1.24 (1.12-1.37)   Type of accommodation   - House = Reference - Flat = 0.98 (0.90-1.06)   Own or rent accommodation   - Own outright = Reference - Own with a mortgage = 1.10 (1.00-1.22) - Rent = 1.01 (0.92-1.10)   Number in household 1.04 (0.96-1.13)  Income   - less than 18 000 1.04 (0.94-1.15) - 18 000 to 30999 = Reference - 31 000 to 51 999 1.00 (0.91-1.10) - greater than 52 000 0.94 (0.85-1.04)   Occupation   - Employed (other) = Reference - Healthcare worker = 1.08 (0.99-1.18) - Unemployed = 1.01 (0.93-1.10) - Retired = 1.02 (0.91-1.14) | Age, sex, ethnicity, education, type of accommodation, number in household, income, occupation |
| Elimian et al. 2020  (Nigeria) | N = 36 496  Community  February 27 2020 – June 8 2020 | | SARS-CoV-2 Infection | 10 517 COVID-19 cases (28.8%) | Socioeconomic determinants = OR (95% CI)  Occupation   - Pupil/ student = Reference - Child = 1.27 (1.00 to 1.60) - Housewife = 1.21 (0.98 to 1.50) - Trader/business = 1.19 (1.05 to 1.35) - Health worker = 0.97 (0.87 to 1.09) - Animal-related work N= 1.21 (0.73 to 2.00) - Farmer = 0.74 (0.59 to 0.93) - Religious / traditional leader = 1.54 (1.00 to 2.38) - Transporter = 0.82 (0.58 to 1.17) - Other = 1.23 (1.12 to 1.36) | Socioeconomic determinants = OR (95% CI)  Occupation   - Pupil/ student = Reference - Child = 1.29 (0.98 to 1.68) - Housewife = 0.93 (0.73 to 1.17) - Trader/business = 1.09 (0.93 to 1.27) - Health worker = 0.90 (0.78 to 1.03) - Animal-related work = 0.93 (0.53 to 1.63) - Farmer   = 0.59 (0.46 to 0.76)   - Religious / traditional leader = 1.13 (0.71 to 1.82) - Transporter = 0.83 (0.56 to 1.21) - Other = 1.11 (0.98 to 1.26) | Age, sex, geopolitical zone, travel history, clinical signs and symptoms |
| Raisi-Estabragh et al. 2020  (UK) | N = 7 099  In-hospital  March 16 2020 – June 14 2020 | | SARS-CoV-2 Infection | 1 439 COVID-19 cases (20.27%)  Ethnicity   - White N = 1 242 (86.3%) - BAME N = 185 (12.9%) | Socioeconomic determinants = OR (95% CI)  Ethnicity   - White = Reference - BAME 2.08 (1.72, 2.50) | Socioeconomic determinants = OR (95% CI)  Ethnicity   - White = Reference - BAME = 1.95 (1.60, 2.36) | Sex, age, BMI, comorbidities, medications and smoking |
| Shahbazi et al. 2020  (Iran) | N = 378  Hospitalised patients in Nahavand county  February 24 2020 – April 21 2020 | | SARS-CoV-2 Infection | 126 COVID-19 cases (33.33%)  Place of residence   - Urban N = 44 (34.92%) - Rural N = 82 (65.08%)   Marital status   - Single N = 18 (14.29%) - Married N = 108 (85.71%) | Socioeconomic determinants = OR (95% CI)  Place of residence   - Urban = 1.89 (1.04-2.51) - Rural = Reference   Marital status   - Single = Reference - Married = 3.69(2.11-6.46) | Socioeconomic determinants = OR (95% CI)  Place of residence   - Urban = 1.79 (1.10-2.91) - Rural = Reference   Marital status   - Single = Reference - Married = 2.08 (1.11-3.91) | Sex, age, marital status, travel history, history of contact with patient, comorbidities, pregnancy, place of residence |
| Guijarroa et al.  2021  (Spain) | N = 152 018  All adults registered in the official municipal population registry of the City Council of Alcorcón  February 1 2020 – April 25 2020 | | SARS-CoV-2 Infection | 1 036 COVID-19 cases (0.68%)   - Spain N = 856 (82.63%) - European Union N = 18 (1.74%) - Eastern Europe and Russia N = 9 (0.87%) - Asia N = 3 (0.29%) - North Africa N = 7(0.68%) - Sub Saharan Africa N = 9 (0.87%) - Latin America N = 121 (11.68%) - Caribbean N = 12 (1.16%) | NA | Socioeconomic determinants = RR (95% CI)   - Spain = Reference - European Union 0.66 (0.43−1.02) - Eastern Europe and Russia= 1.23 (0.49−3.06) - Asia = 0.64 (0.27−1.52) - North Africa = 1.11(0.37−3.31) - Sub Saharan Africa = 3.66(1.42−9.41) - Latin America = 6.92(4.49−10.67) - Caribbean= 6.35(3.83−10.55) | Age and sex |
|  |  |  | Hospitalisation for Covid-19 | 877 COVID-19 hospitalisations (0.58%) | NA | Socioeconomic determinants = RR (95% CI)   - Spain = Reference - European Union 0.64 (0.33−1.24) - Eastern Europe and Russia= 0.95 (0.35−2.54) - Asia = 0.57 (0.20−1.66) - North Africa = 1.46 (0.50−4.24) - Sub Saharan Africa = 3.32 (1.29−8.53) - Latin America = 7.65(4.99−11.72) - Caribbean= 6.39 (3.79−10.76) | Age and sex |
| Hanson et al.  2020    (USA) | N = 3 892  Patients at Indiana University Health or Eskenazi Health hospitals  January 1, 2020 – April 30 2020 | | SARS-CoV-2 Infection | 3 892 COVID-19 cases  African American N = 1 024 (33%)  Hispanic N = 507 (16%) |  | Socioeconomic determinants = OR (95% CI)  Race:   - African American = 4.58(4.25-4.94) - Non-African American = Reference   Ethnicity:   - Hispanic = 2.58(2.34-2.83)   Non-Hispanic = Reference | Ethnicity, race |
| Kolin et al.  2020  (UK) | N = 397 064  Participants from England  between the ages of40 and 69 recruited from 2006 to 2010  March 16 2020 – May 18 2020 | | SARS-CoV-2 Infection | 968 COVID-19 cases (0;24%)  Race   - Asian N = 52 (5.37%) - Black N = 61 (6.3%) - White N =839 (86.67%) | Socioeconomic determinants = RR (95% CI)  Race   - Asian = 2.28 (1.72-3.01) - Black = 3.66 (2.83-4.74) - White = Reference   Townsend deprivation index = 1.12 (1.10-1.14) | Socioeconomic determinants = RR ( CI 95%)  Race   - Asian = 2.13 (1.60-2.85) - Black = 2.53 (1.92-3.33) - White = Reference   Townsend deprivation index = 1.09 (1.07-1.12) | Age, sex, body-mass index, systolic blood pressure, race Townsend deprivation index |
| Mendez-Dominquez et al. 2020  (Mexico) | N = 17 763  Community  February 28, 2020 - April 21, 2020 | | SARS-CoV-2 Infection | 9 501 COVID-19 cases (53.49%) | NA | Socioeconomic determinants = IRR ( CI 95%)   - Migration = 1.05(1.05-1.06) - Indigenous ethnicity = 0.89(0.88-0.90) - Population affiliated to public health institutions = 0.81(0.79-0.81) - Urbanisation = 1.63(1.62-1.64)   Socioeconomic determinants = OR (95% CI)   - Interstate migrant = 0.69(0.61-0.77) | Population age and gender composition  Age, gender, and place of residence of patients |
|  |  |  | Hospitalisation for Covid-19 | 5 545 COVID-19 hospitalisations (31.22%) | NA | Socioeconomic determinants = IRR (95% CI)   - Migration = 0.65(0.58-0;74) - Indigenous ethnicity = 1.01(0.83-1.02) - Population affiliated to public health institutions = 1.27(1.25-1;29) - Urbanisation = 1.1(1.09-1.1)   Socioeconomic determinants = OR (95% CI)  Interstate migrant = 1.36(1.19-1.54) | Population age and gender composition  Age, gender, and place of residence of patients |
| Aung et al. 2021  (USA) | N = 36 041  World-wide mobile application-based cohort | | SARS-CoV-2 Infection | 484 infections (1.34%)  White N =403 (83.26%)  Black N = 7 (1.45%)  Hispanic N = 47 (9.7%)  Asian/Pacific Islander N = 10 (2.07%)  Other/multi-racial N = 9 (2%) | NA | Socioeconomic determinants = OR (95% CI)  Ethnicity:   - White = Reference - Black = 1.39 (0.61-3.17) - Hispanic = 1.31 (0.95-1.81) - Asian/Pacific Islander = 0.40 (0.21-0.76) - Other/multi-racial = 0.82 (0.40-1.65)   Education:   - Low level = Reference - High level = 0.90 (0.84-0.951) | Primary residence in USA , age, race/ethnicity, gender,health care worker, children living with you,any pets at home, flu shot within 1 year, Marijuana Use, immunodeficiency, health care worker, highest education level |
| Raisi-Estabragh  2020  (UK) | N = 4 510  General population  March 16 2020 - May 18 2020 | | SARS-CoV-2 Infection | 1 326 COVID-19 infections (29.4%)  BAME ethnicities N = 174 (13.1%) | Socioeconomic determinants = OR (95% CI)  BAME = 1.85 (1.51-2.25)  Townsend deprivation score = 1.01 (1.02-1.06)  Household size = 1.12(1.06-1.17) | Socioeconomic determinants = OR (95% CI)  BAME = 1.59(1.26-1.99)  Townsend deprivation score = 1.03(1.01-1.06)  Household size = 1.09(1.03-1.16) | Sex, age, ethnicity, BMI, Townsend deprivation score, household size |
| Rossi 2020  (Italy) | N = 2 635  General population in Northern Italy  March 3 2020 – March 26 2020 | | SARS-CoV-2 Infection | 1 564 COVID-19 infections  Italians N = 1 442 (54.72%)  Immigrants N = 122 (4.63%) | NA | Socioeconomic determinants = OR (95% CI)  Country of birth   - Italians = Reference - Immigrants = 0.99(0.82-1.2) | Age and sex |
| Zelner 2021  (USA) | N =49 701  General population  March 8 2020 - July 5 2020 | | SARS-CoV-2 Infection | SARS-CoV-2 Infection | NA | Socioeconomic determinants = MRR ( CI 95%)  Race   - Black = 5.5 (5.4- 5.6) - Latino = 3.1(3- 3.2) - Other = 3.9 (3.7- 4.1) - Asian/Pacific = 1.7 (1.6- 1.8) - White = Reference - Native American = 1 (0.8- 1.2) | Sex and age |
| Ho et al. 2020  (UK) | N = 235 928  In-hospital setting  March 16 2020 – May 3 2020 | | SARS-CoV-2 Infection | SARS-CoV-2 Infection | Socioeconomic determinants = RR (p-value)  Ethnicity   - White = Reference - Black = 2.88 (0.0001) - South Asian = 2.15(0.009) - Others = 1.99 (0.009)   Deprivation score = 1.3 (<0.0001) | Socioeconomic determinants = RR (p-value)  Ethnicity   - White = Reference - Black = 2 (0.02) - South Asian = 1.73 (0.08) - Others = 1.7 (0.053)   Deprivation score = 1.17(0.001) | Age, sex, ethnicity and deprivation, body mass index (BMI), forced expiratory volume in 1 s (FEV1), and walking pace |
| Atkins et al. 2020  (UK) | N = 268 563  In-hospital setting  March 16 2020 – April 26 2020 | | SARS-CoV-2 Infection | 507 SARS-CoV-2 Infection (0.2%) | NA | Socioeconomic determinants = RR (95% CI)  Ethnicity   - White = Reference - Black = 2.85 (1.71-4.74) - South Asian = 1.69 (1-2.85) - Others = 2 (1.11-3.61)   Education   - None = 2.06 (1.6 – 2.66) - School/College = 1.11 (0.85-1.45) - Professional qualification = 1.31 (0.96-1.79) - Degree = Reference | Age group, sex, ethnicity, education, baseline assessment center, and all comorbidities |
| Mathur et al. 2021  (UK) | N = 17 288 532  Adults (aged ≥18 years) registered with primary care practices in England   - Wave 1: February 1 2020 – August 3 2020 | | SARS-CoV-2 Infection | 71 246 SARS-CoV-2 Infection (0.4%) | Socioeconomic determinants = HR (95% CI)  Ethnicity   - White = Reference - South Asian = 2·38 (2·32–2·43) - Black = 1·82 (1·74–1·90) - Mixed = 1·37 (1·28–1·46) - Other = 1·06 (1·00–1·12) - Unknown = 0·97 (0·95–0·99) | Socioeconomic determinants = HR (95% CI)  Ethnicity   - White = Reference - South Asian = 1·99 (1·94–2·04) - Black = 1·69 (1·62–1·77) - Mixed = 1·49 (1·39–1·59) - Other = 1·20 (1·14–1·28) - Unknown = 1·06 (1·04–1·08) | Age, sex, deprivation, comorbidities, household size |
|  | N = 17 636 366   - Wave 2: September 1 2020 – December 31 2020 | | SARS-CoV-2 Infection | 506 773 SARS-CoV-2 Infection (2.9%) | Socioeconomic determinants = HR (95% CI)  Ethnicity   - White = Reference - South Asian = 1.67 (1.66 - 1.69) - Black = 1.01 (0.99 - 1.03) - Mixed = 1.15 (1.12 - 1.18) - Other = 0.79 (0.77 - 0.80) - Unknown = 1.06 (1.05 - 1.07) | Socioeconomic determinants = HR (95% CI)  Ethnicity   - White = Reference - South Asian = 1.32 (1.31 - 1.33) - Black = 0.85 (0.84 - 0.87) - Mixed = 0.98 (0.95 - 1.01) - Other = 0.72 (0.70 - 0.73) - Unknown = 1.04 (1.03 - 1.05) | Age, sex, deprivation, comorbidities, household size |
|  | N = 17 288 532  Adults (aged ≥18 years) registered with primary care practices in England   - Wave 1: February 1 2020 – August 3 2020 | | Hospitalisation for COVID-19 | 32 473 COVID-19 hospitalisations (0.2%) | Socioeconomic determinants = HR (95% CI)  Ethnicity   - White = Reference - South Asian = 1·26 (1·21–1·31) - Black = 1·39 (1·30–1·48) - Mixed = 0·86 (0·77–0·97) - Other = 0·77 (0·71–0·85) - Unknown = 0·91 (0·88–0·93) | Socioeconomic determinants = HR (95% CI)  Ethnicity   - White = Reference - South Asian = 1·48 (1·41–1·55) - Black = 1·78 (1·67–1·90) - Mixed = 1·63 (1·45–1·83) - Other = 1·54 (1·41–1·69) - Unknown = 1·06 (1·03–1·09) | Age, sex, deprivation, comorbidities, household size |
|  | N = 17 636 366   - Wave 2: September 1 2020 – December 31 2020 | | Hospitalisation for COVID-19 | 18 855 COVID-19 hospitalisations (<0.1%) | Socioeconomic determinants = HR (95% CI)  Ethnicity   - White = Reference - South Asian = 1.71 (1.62 - 1.79) - Black = 1 (0.90 - 1.11) - Mixed = 0.75 (0.63 - 0.89) - Other = 0.69 (0.60 - 0.79) - Unknown = 0.87 (0.84 - 0.91) | Socioeconomic determinants = HR (95% CI)  Ethnicity   - White = Reference - South Asian = 1.89 (1.79 - 2) - Black = 1.23 (1.11 - 1.37) - Mixed = 1.33 (1.12 - 1.58) - Other = 1.32 (1.16 - 1.51) - Unknown = 1.07 (1.03 - 1.11) | Age, sex, deprivation, comorbidities, household size |
|  | N = 17 288 532  Adults (aged ≥18 years) registered with primary care practices in England   - Wave 1: February 1 2020 – August 3 2020 | | ICU admission | 3 096 ICU admissions (<0.1%) | Socioeconomic determinants = HR (95% CI)  Ethnicity   - White = Reference - South Asian = 2·38 (2·12–2·67) - Black = 3·08 (2·63–3·60) - Mixed = 1·97 (1·51–2·57) - Other = 1·83 (1·48–2·25) - Unknown = 0·95 (0·87–1·04) | Socioeconomic determinants = HR (95% CI)  Ethnicity   - White = Reference - South Asian = 2·18 (1·92–2·48) - Black = 3·12 (2·65–3·67) - Mixed = 2·96 (2·26–3·87) - Other = 3·18 (2·58–3·93) - Unknown = 1·08 (0·99–1·19) | Age, sex, deprivation, comorbidities, household size |
|  | N = 17 636 366   - Wave 2: September 1 2020 – December 31 2020 | | ICU admission | 3 351 ICU admissions (<0.1%) | Socioeconomic determinants = HR (95% CI)  Ethnicity   - White = Reference - South Asian = 2.85 (2.57 - 3.16) - Black = 1.65 (1.35 - 2.02) - Mixed = 1.43 (1.06 - 1.93) - Other = 1.05 (0.81 - 1.37) - Unknown = 0.84 (0.77 - 0.92) | Socioeconomic determinants = HR (95% CI)  Ethnicity   - White = Reference - South Asian = 2.68 (2.39 - 3.01) - Black = 1.67 (1.37 - 2.05) - Mixed = 2.23 (1.65 - 3.02) - Other = 1.9 (1.46 - 2.46) - Unknown = 1.01 (0.92 - 1.11) | Age, sex, deprivation, comorbidities, household size |
|  | N = 17 288 532  Adults (aged ≥18 years) registered with primary care practices in England   - Wave 1: February 1 2020 – August 3 2020 | | Mortality | 11 469 COVID-19 deaths (<0.1%) | Socioeconomic determinants = HR (95% CI)  Ethnicity   - White = Reference - South Asian = 0·89 (0·83–0·97) - Black = 0·96 (0·85–1·09) - Mixed = 0·51 (0·40–0·65) - Other = 0·44 (0·37–0·54) - Unknown = 0·95 (0·91–0·99) | Socioeconomic determinants = HR (95% CI)  Ethnicity   - White = Reference - South Asian = 1·26 (1·15–1·37) - Black = 1.51 (1.33–1.71) - Mixed = 1·41 (1·11–1·81) - Other = 1·22 (1·00–1·48)   Unknown = 1·01 (0·97–1·06) | Age, sex, deprivation, comorbidities, household size |
|  | N = 17 636 366   - Wave 2: September 1 2020 – December 31 2020 | | Mortality | 7 366 COVID-19 deaths ((<0.1%) | Socioeconomic determinants = HR (95% CI)  Ethnicity   - White = Reference - South Asian = 1.17 (1.07 - 1.28) - Black = 0.52 (0.42 - 0.66) - Mixed = 0.38 (0.26 - 0.55) - Other = 0.28 (0.20 - 0.40)   Unknown = 1 (0.94 - 1.05) 1 | Socioeconomic determinants = HR (95% CI)  Ethnicity   - White = Reference - South Asian = 1.87 (1.68 - 2.07) - Black = 0.92 (0.73 - 1.16) - Mixed = 1.24 (0.85 - 1.83) - Other = 0.92 (0.66 - 1.29)   Unknown = 1.17 (1.11 - 1.24) | Age, sex, deprivation, comorbidities, household size |
| Clift et al. 2020  (UK) | N=6 083 102  Primary care patients  Inclusion on January 24 2020 | | Hospitalisation for COVID-19 | N= 10 776 (0.18%)  Ethnicity   - White N=6790 (63.01%) - Indian N=423 (3.93%) - Pakistani N=248 (2.30%) - Bangladeshi N=173 (1.61%) - Other Asian N=248 (2.30%) - Caribbean N=392 (3.64%) - Black African N=456 (4.23%) - Chinese N=45 (0.42%) - Other ethnic groups N=436 (4.05%) - Not recorder N=1565 (14.52%)   Accommodation   - Not in care home or homeless N=9895 (91.82%) - Lives in care home or homeless N=854 (7.93%) - Homeless according to GP records N=27 (0.25%)1   Townsend deprivation fifth   - 1 (most affluent) N=1799 (16.69%) - 2 N=1886 (17.50%) - 3 N=2114 (19.62%) - 4 N=2338 (21.70%) - 5 (most deprived) N=2612 (24.24%)   Not recorded N=27 (0.25%) | NA | Socioeconomic determinants = HR(95%CI)  Females:  Townsend material deprivation score (5 unit increase) = 1.52 (1.45 to 1.60)  Ethnicity   - White = reference - Indian = 1.89 (1.6-2.24) - Pakistani = 1.52 (1.21-1.89) - Bangladeshi = 1.41 (1.11-1.79) - Other Asian= 2.14 (1.74-2.64) - Caribbean= 2.01 (1.71-2.35) - Back African = 2.30 (1.97-2.68) - Chinese = 1.15 (0.71-1.85) - Other ethnic groups=1.90 (1.64-2.21)   Accommodation   - Not in care home or homeless=reference - Lives in care home or homeless= 1.84 (1.64-2.07) - Homeless according to GP records 1.23 (0.55-2.74)   Males:  Townsend material deprivation score (5 unit increase) = 1.46 (1.40 to 1.53)  Ethnicity   - White = reference - Indian 2.15 (1.89-2.44) - Pakistani = 2.01 (1.72-2.36) - Bangladeshi = 1.71 (1.41-2.08) - Other Asian= 2.29 (1.91-2.74) - Caribbean =2.29 (1.99-2.63) - Back African =2.59 (2.27-2.97) - Chinese= 1.51 (1.03-2.20) - Other ethnic groups=2.12 (1.83-2.46)   Accommodation   - Not in care home or homeless=reference - Lives in care home or homeless=2.52 (2.25-2.82) - Homeless according to GP records=1.5 (0.97-2.30) | Townsend score, BMI, age, comorbidities, ethnicity, domicile, treatments |
|  |  | | Mortality | N= 4384 (0.07%)  Ethnicity   - White N=2947 (67.22%) - Indian N=131 (2.99%) - Pakistani N=69 (1.57%) - Bangladeshi N=69 (1.57%) - Other Asian N=57 (1.30%) - Caribbean N=152 (3.47%) - Black African N=122 (2.78%) - Chinese N=18 (0.41%) - Other ethnic groups N=114 (2.60) - Not recorded N=705 (16.08%)   Accommodation   - Not in care home or homeless N=3345 (76.30%) - Lives in care home or homeless N=1033 (23.56%) - Homeless according to GP records N=6 (0.14%)   Townsend deprivation fifth   - 1 (most affluent) N=840 (19.16%) - 2 N=746 (17.02%) - 3 N=934 (21.30%) - 4 N=951 (21.69%) - 5 (most deprived) N=905 (20.64%)   Not recorded N=8 (0.18%) | NA | Socioeconomic determinants = HR(95%CI)  Females:  Townsend material deprivation score (5 unit increase) = 1.48 (1.37-1.61)  Ethnicity   - White = reference - Indian = 1.89 (1.43-2.51) - Pakistani = 1.40 (0.91-2.14) - Bangladeshi = 1.41 (0.88-2.26) - Other Asian= 1.19 (0.72-1.97) - Caribbean= 1.68 (1.29-2.20) - Back African = 1.98 (1.39-2.83) - Chinese = 1.21 (0.51-2.90) - Other ethnic groups=1.73 (1.28-2.35)   Accommodation   - Not in care home or homeless=reference - Lives in care home or homeless= 3.61 (3.18-4.10) - Homeless according to GP records=1.48 (0.21-10.52)   Males :  Townsend material deprivation score (5 unit increase) = 1.50 (1.40 to 1.61)  Ethnicity   - White = reference - Indian=1.59 (1.25-2.01) - Pakistani = 1.84 (1.39-2.44) - Bangladeshi = 2.27 (1.65-3.12) - Other Asian= 2.02 (1.49-2.74) - Caribbean =2.06 (1.65-2.57) - Back African =3.03 (2.42-3.80) - Chinese= 2.47 (1.49-4.09) - Other ethnic groups=2.04 (1.60-2.58)   Accommodation   - Not in care home or homeless=reference - Lives in care home or homeless=4.28 (3.80-4.83) - Homeless according to GP records=1.56 (0.65-3.76) | Townsend score, BMI, age, comorbidities, ethnicity, domicile, comorbidities and treatments |
| Izurieta et al.  2021  (USA) | N = 25 333 329  Medicare fee-for-service beneficiaries | | Hospitalisation for COVID-19 | 27,961 Hospitalizations (0.11%)  12 613 Deaths (0.05%) | NA | Ethnicity   - White = Reference - Black = 2.81(2.62-3.02) - Hispanic = 3.31(2.83-3.87) - North American Native = 4.22(2.9-6.16) - Asian = 1.5(1.23-1.82) - Other/Unknown = 1.16(1.01-1.32) | Age, sex, ethnicity, Covid-19 circulation rate, influenza vaccination status, presence of numerous medical conditions, Frailty index, hospital admission, area deprivation index, population density |
|  |  | | Mortality | 27 961 hospitalizations (0.11%)  12 613 deaths (0.05%) | NA | Socioeconomic determinants = OR (95% CI)  Ethnicity   - White = Reference - Black = 2.47 (2.17–2.81) - Hispanic = 3.11 (2.37–4.08) - North American Native = 5.82 (3.25–10.43) - Asian = 1.32 (.93–1.87) - Other/Unknown = 1.19 (.95–1.50) | Age, sex, ethnicity, Covid-19 circulation rate, influenza vaccination status, presence of numerous medical conditions, Frailty index, hospital admission, area deprivation index, population density |
| McKeique et al.  2020  (UK) | N = 41 220  ICU admission/ control  Inclusion on June 14 2020 | | Severe COVID-19 | 4 272 cases of severe COVID-19 (10.36%)  Scottish index of Multiple Deprivation: Indicator based on postal code:   - 1 - 2 N = 8 891 - 3 N = 7 556 - 4 N = 7 331 - 5 N = 7 733   Care Home Residence N= 4 829  Ethnicity   - White - South Asian N = 172 - Black N = 40 - Other N = 162 | Socioeconomic determinant = RR (95% CI)  Scottish index of Multiple Deprivation: Indicator based on postal code:   - 1 = Reference - 2 = 0.86 (0.78–0.95) - 3 = 0.88 (0.79–0.98) - 4 = 0.81 (0.72–0.90) - 5 = 0.54 (0.48–0.62)   Care Home Residence = 21.4 (19.1–23.9)  Ethnicity   - White = Reference - South Asian = 1.26 (0.81–1.97) - Black = 1.16 (0.44–3.04) - Other = 1.01 (0.63–1.65) | NA | NA |
| Mutambudzi et al. 2021  (UK) | N = 120 075  In-hospital setting  March 16 2020 – July 26 2020 | | Severe COVID-19 | Occupation   - Non-essential workers N = 112 (0.1%) - Healthcare workers N = 102 (0.9%) - Social and education workers N = 31 (0.2%) - Other essential workers N = 26 (0.2%) | NA | Socioeconomic determinant = RR (95% CI)  Occupation   - Non-essential workers = Reference - Healthcare workers = 7.69 (5.58 -10.60) - Social and education workers = 1.88 (1.21 -2.91) - Other essential workers= 1.15 (0.75 -1.77)   Socioeconomic deprivation   - Quartile 1 = Reference - Quartile 2 = 1.32 (0.87 -2.00) - Quartile 3 = 1.54 (1.04 -2.29) - Quartile 4 (least advantaged) = 1.77 (1.20 -2.63)   Ethnicity   - White British = Reference - White Irish = 1.59 (0.85 -2.98) - White Other = 1.24 (0.62 -2.46) - Mixed = 1.32 (0.42 -4.11) - South Asian = 4 (2.26 -7.07) - Black = 2.73 (1.61 -4.62) - Other = 1.36 (0.55 -3.36)   Education   - College or University degree = Reference - A levels/AS levels or equivalent = 1.00 (0.66 -1.54) - O levels/GCSEs/CSEs or equivalent = 1.42 (1.05 -1.93) - Other = 1.07 (0.70 -1.62) - None of the above = 1.74 (1.01 -3.01) | Sociodemographic factors, socioeconomic factors, work-related factors, comorbidities, disability, lifestyle-related factors |
| Bassett et al. 2020  (USA) | N = 306 800 000  Community | | Mortality | N = 128 720 deaths (0.04%)  White N = 68 377 (53.12%)  Black N = 29 476 (22.9%)  Hispanic N = 23 256 (18.07%)  Indian/Alaska N = 1143 (0.89%)  Asian N = 6468 (5.02%) | NA | Socioeconomic determinants = RR (95% CI)   - White = Reference - Black = 3.6 (3.5 - 3.7) - Hispanic = 2.8 (2.7 – 3) - Indian/Alaska = 2.2 (1.8 – 2.6)   Asian = 1.6 (1.4 – 1.7) | Age |
| AyoubKhani et al. 2020  (UK) | N = 47 872 412  Community (residents of England and Wales) | | Mortality | NA | NA | Socioeconomic determinants = HR (95% CI)  Male:   - White = Reference - Bangladeshi/Pakistani = 1.351 (1.219 – 1.497) - Black 1.773 (1.644–1.913) - Chinese = 1.025 (0.818– 1.284) - Indian = 1.305 (1.194–1.426) - Mixed = 1.09 (0.923– 1.286) - Other = 1.367 (1.237–1.104)   Female:   - White = Reference - Bangladeshi/Pakistani = 1.038 (0.897–1.201) - Black = 1.288 (1.170–1.417) - Chinese = 1.139 (0.865–1.5) - Indian = 0.902 (0.801–1.014) - Mixed = 1.006 (0.823–1.230) - Other =0.958 (0.831–1.104) | Age, population density, LAD, deprivation, SES, education, household composition, occupational exposure, self-reported health |
| Brandén et al. 2020  (Sweden) | N = 274 712  Community- All individuals living in Stockholm county  Inclusion on March 12 2020 | | Mortality | 1301 deaths (0.47%)  m² per individual in household   - 0 to < 20 N = 88 6.76% - 20 to <30 N = 183 (14.07%) - 30 to < 40 N = 325 (24.98%) - 40 to < 60 N = 320 (24.60%) - >=60 N = 383 (29.44%) - Missing < 5   Housing   - Multi-family housing N = 702 (53.96%)   - Single family housing N = 219 (16.83%)   - Care home N = 380 (29.21%)   Country of birth   - Sweden N = 928 (71.33%) - HIC N = 211 (16.22%) - LMIC MENA N = 83 (6.38%) - LMIC others N = 79 (6.07%)   Educational level   - Primary N = 415 (31.90%) - Secondary N = 506 (38.89%)   - Post-secondary N = 308 23.67%   - Missing N = 72 (5.53%)   Individual disposable income   - Lowest tertile N = 709 (54.50%) - Mid tertile N = 439 (33.74%) - Highest tertile N = 153 (11.76%) | NA | m² per individual in household   - 0 to < 20 = 2.10 (1.53–2.87) - 20 to <30 = 1.74 (1.39–2.18) - 30 to < 40 = 1.57 (1.30–1.90) - 40 to < 60 =1.13 (0.95–1.33) - >=60 = Reference   Housing   - Multi-family housing = Reference   - Single family housing = 1.03 (0.85–1.24)   - Care home = 4.13 (3.49–4.90)   Country of birth   - Sweden = Reference - HIC = 1.09 (0.93-1.27) - LMIC MENA= 2.02 (1.55-2.63) - LMIC others = 1.24 (0.97-1.6)   Educational level   - Primary = 1.17 (0.99-1.37) - Secondary = 1.18 (1.02-1.37)   - Post-secondary = Reference   - Missing = 1.13 (0.84-1.52)   Individual disposable income   - Lowest tertile = 1.38 (1.13-1.67) - Mid tertile = 1.18 (0.98-1.43) - Highest tertile = Reference | Age structure of hosuehold, housing incidence of COVID-19 per 10 000 habitants in borough, individuals per km2 in neighberhoods, country of birth, educational level, indiviudal disposable income, sex, m² per individual in household |
| Rostila et al. 2021  (Sweden) | N = 1 778 670  Community - Adult population aged 21+ living in Stockholm | | Mortality | NA | NA | Socioeconomic determinants = RR  Country of birth   - Sweden = Reference - Other Nordic countries = 1.25 (1.03 -1.52) - Europe = 0.91(0.74-1.12) - Middle East = 1.96(1.56-2.46) - Africa = 1.7 (1.17- 2.4) - Rest of the world = 0.84 (0.58-1.22)   Income  Divided into quintiles   - 1 (least) = 1.49 (1.18 -1.9) - 2 = 1.35 (1.07- 1.71) - 3 = 1.03 (0.77 - 1.37) - 4 (most) = Reference - NA = 0.42 (0.06 -3.21)   Education   - Primary = 1.24 (1.06- 1.45) - Secondary = 1.26 (1.09- 1.46) - Postsecondary = Reference - Missing data = 1.21 (0.91-1.61)   Employment status   - Employed = Reference - Unemployed = 2.27 (1.81- 2.84)   Housing type   - House or apartment = Reference - Special care = 5.93 (5.07- 6.94)   Working age in Household   - 0 people = Reference - 1-2 people = 1.61 (1.36- 1.9) - >=3 people = 2.32 (1.72- 3.12) | Age, sex, education level, employment, disposable income, HH type, number of working age individuals in household, population density |
| Drefahl et al.  2020  (Sweden) | N = 7 775 054  Community  March 12 2020 | | Mortality | 3126 deaths  Civil status   - Married N= 1032 (33%) - Never married N= 348   (11.1%)   - Divorced N = 598 (19.1%) - Widowed N = 1148 (36.7%)   Education   - Primary N =1224 (39.2%) - Secondary N = 1183(37.8%) - Post-secondary N =577 (18.5%) - Missing N = 1148(4.5%)   Individual net income   - Tertile 1 (low) N = 1916 (61.3%) - Tertile 2 N =894 (28.6%) - Tertile 3 (high) N =316 (10.1%)   Country of birth   - Sweden N =2406 (77%) - HIC N = 363(11.6%) - LMIC MENA N = 195 (6.2%) - LMIC other N = 162(5.2%)   County of residence   - Stockholm N =1557 (49.8%) - Other N = 1569 (50.2%) | Socioeconomic determinants = HR  UnAdjusted  Civil status   - Married = Reference - Never married = 1.61(1.42 -1.82) - Divorced = 1.54(1.39- 1.71) - Widowed = 1.11(1.01-1.22)   Education   - Primary = 1.15 (1.04 - 1.27) - Secondary = 1.22 (1.1-1.35) - Post-secondary = Reference   Individual net income   - Tertile 1 (low) = 1.22 (1.08-1.39) - Tertile 2 = 1.26 (1.11-1.44) - Tertile 3 = Reference   Country of birth   - Sweden = Reference - HIC = 1.52 (1.36 -1.7) - LMIC MENA = 5.06 (4.3 - 5.95) - LMIC other = 2.76 (2.38 - 3.19)   County of residence   - Stockholm = 4.5 (4.2 - 4.83) - Other = Refrence | Socioeconomic determinants = HR  Adjusted  Civil status   - Married = Reference - Never married = 1.66 (1.46-1.88) - Divorced = 1.57 (1.41-1.73) - Widowed = 1.48(1.34-1.63)   Education   - Primary = 1.34 (1.2-1.49 - Secondary = 1.3 (1.17-1.44) - Post-secondary = Reference   Individual net income   - Tertile 1 (low) = 1.6 (1.4-1.83) - Tertile 2= 1.32 (1.16-1.51) - Tertile 3 = Reference   Country of birth   - Sweden = Reference - HIC = 1.14(1.02-1.28) - LMIC MENA = 2.77 (2.31-3.32) - LMIC other = 1.9 (1.62-2.22)   County of residence   - Stockholm = 4.63 (4.3-4.99) - Other = Refrence | Sex, country of birth, and living in Stockholm, highest achieved educational degree and individual net income, marital status |
| Elliott et al.  2021  (UK) | N=473 550  Population based | | Mortality | N=459 (0.10%)  Education   - High N=87 (19.91%) - Intermediate N=188 (43.02%) - Low N=162 (37.07%)   Type of accommodation   - House N=369 (92.92%) - Flat N=76 (17.98%)   Own/rent   - Own outright N=258 (58.64%) - Own with a mortgage N=91 (20.68%) - Rent N=91 (20.68%)   Number in household N=443  Income   - 18000 to 30999 N=93 (25.48%) - Less than 18 000 N=171 (46.85%) - 31000 to 51999 N=62 (16.99%) - Greater than 52000 N=39 (10.68%)   Occupation   - Employed N=102 (22.47%) - Health care worker N=20 (4.41%) - Unemployed N=78 (17.18%) - Retired N=254 (55.95%)   Ethnicity   - White N=415 (91.41%) - Black N=23 (5.07%)   Other N=16 (3.52%) | Socioeconomic determinants =OR(95%CI)  Education   - High = reference - Intermediate=1.43(1.11-1.85) - Low =3.79 (2.92-4.91)   Type of accommodation   - House=reference - Flat=1.91 (1.49-2.44)   Own/rent   - Own outright=reference - Own with a mortgage=0.48 (0.38-0.61) - Rent=1.98 (1.56-2.52)   Number in household 0.63 (0.56-0.72)  Income   - 18000 to 30999=reference - Less than 18 000=2.14 (1.66-2.76) - 31000 to 51999=0.63 (0.46-0.87) - Greater than 52000=0.40 (0.27-0.58)   Occupation   - Employed=reference - Health care worker=1.66 (1.03-2.68) - Unemployed=2.82 (2.10-3.79) - Retired =4.31 (3.43-5.43)   Ethnicity   - White=reference - Black=3.17 (2.08-4.82) - Other=0.94(0.57-1.55) | Socioeconomic determinants=OR(95%CI)  Education   - High =reference - Intermediate=1.06 (0.90-1.25) - Low =1.11 (0.97-1.27)   Type of accommodation   - House=reference - Flat=1.09 (0.97-1.22)   Own/rent   - Own outright=reference - Own with a mortgage=1.06 (0.90-1.26) - Rent=1.08 (0.95-1.21)   Number in household=1.00 (0.86-1.16)  Income   - 18000 to 30999=reference - Less than 18 000=1.10 (0.96-1.27) - 31000 to 51999=1.09 (0.92-1.30) - Greater than 52000=1.11 (0.90-1.36)   Occupation   - Employed=reference - Health care worker=1.29 (1.12-1.48) - Unemployed=1.10 (0.95-1.28) - Retired =1.10 (0.93-1.30)   Ethnicity   - White=reference - Black=1.21 (1.12-1.48) - Other=0.96(0.83-1.11) | Sex, smoking status, alcohol drinker, BMI, cholesterol, HDL, triglycerides, vitamin D, cystatin, cancer, CVD, diabetes, respiratory, autoimmune, ACR inhibitors, angiotensin II RB, oral steroid, statin, environmental factors |
| Williamson et al. 2020  (UK) | N = 17 278 392  General population registered with a general practice  February 1 2020 – May 6 2020 | | Mortality | 10 926 COVID-19 deaths (0.06%)  Ethnicity   - White N = 7,119 (0.07%) - Mixed N = 62 (0.04%) - South Asian N= 608 (0.06%) - Black N = 250 (0.07%) - Other N= 110 (0.03%)   IMD quintile:   - 1 (least deprived) N= 1 908 (0.05%) - 2 N= 2 030 (0.06%) - 3 N= 2 114 (0.06%) - 4 N= 2 388 (0.07%) - 5 N= 2 486 (0.07%) | NA | Socioeconomic determinants = OR (95% CI)  Model 1:  Ethnicity   - White = Reference - Mixed = 1.62 (1.26–2.08) - South Asian = 1.69 (1.54–1.84) - Black = 1.88 (1.65–2.14) - Other = 1.37 (1.13–1.65)   IMD quintile:   - 1 (least deprived) = Reference - 2 = 1.16 (1.08–1.23) - 3 =1.31 (1.23–1.40) - 4 = 1.69 (1.59–1.79) - 5 = 2.11 (1.98–2.25)   Model 2:  Ethnicity   - White = Reference - Mixed = 1.43 (1.11–1.84) - South Asian = 1.45 (1.32–1.58) - Black = 1.48 (1.29–1.69) - Other = 1.33 (1.10–1.61)   IMD quintile:   - 1 (least deprived) = Reference - 2 = 1.12 (1.05–1.19) - 3 = 1.22 (1.15–1.30) - 4 = 1.51 (1.42–1.61) - 5 = 1.79 (1.68–1.91) | Model 1:  Age and sex  Model 2:  Age, sex, smoking, BMI, comorbidities and other health conditions, IMD quintile, ethnicity |
| Mak et al. 2021  (UK) | N = 410 199  Community (UK- Biobank)  March 16, 2020 - November 30, 2020 | | Mortality | 514 COVID-19 deaths (0.125%) | NA | Socioeconomic determinants = OR (95% CI)  Ethnicity   - White = Reference - Asian = 2.04(1.15-3.6) - Black = 3.93(2.38-6.5) - Others = 1.06(0.39-2.87)   Education   - High = Reference - Intermediate = 1.27(0.96-1.66) - Low = 1.49(1.09-2.02)   Income   - >£52,000 = Reference - £31,000–51,999 = 1.34(0.89-2.02) - 18,000–30,999= 1.29(0.86-1.92) - < £18,000 = 1.85(1.24-2.76)   Townsend deprivation quintile:   - 1 = Reference - 2 = 0.88(0.62-1.26) - 3 = 0.98(0.69-1.39) - 4 = 1.16(0.83-1.63)   5 = 1.62(1.17-2.24) | Frailty (concurrent HFRS), comorbidity (concurrent CCI), age (continuous), sex, smoking status, education, income and deprivation |
| Nafilyan et al. 2021  (UK) | N = 10 078 568  residents of England aged 65 years  March 2, 2020 - November 30, 2020 | | Mortality | 39,419 (0.39%) COVID-19 deaths |  | Socioeconomic determinants = HR (95% CI)  Living in household  Men:  Single = 1.132 (1.091–1.174)  Multi-generational household without children = 1.065 (1.009–1.125)  Multi-generational household with children = 1.174 (1.062–1.299)  3+ elderly adults = 0-995 (0.824–1.080)  Women  Single = 1.105 (1.059–1.152)  Multi-generational household without children = 1.156 (1.073–1.246)  Multi-generational household with children = 1.21 (1.060–1.380)  3+ elderly adults = 0.845 (0.658–1.086) | Geographical factors (region, population density, urban/rural classification), ethnicity, socioeconomic characteristics (IMD decile, household deprivation, educational attainment, social grade, household tenancy), health (self-reported health and disability from the Census , pre-existing conditions based on hospital contacts, number of hospital admissions, total days spent in hospital), a measure for overcrowding and property type. |
| **Ecological studies** | | | | | | | |
| Akanbi et al. 2021  (USA) | 70 zip codes  COVID-19 in 70 zip codes in Oakland County | | SARS-CoV-2 Infection | SARS-CoV-2 Infection | Socioeconomic determinants = IRR (95% CI)   - Ethnicity = 1.03 (1.02- 1.04) - Education level = 0.99 (0.98-1.01) - Median family income = 0.78 (0.65- 0.93) - Poverty level = 1.05 (01.02- 1.08) | Socioeconomic determinants = IRR (95% CI)   - Ethnicity = 1.02 (1.01; 1.03) - Education level = 1.03 (1; 1.05) - Median family income = 0.76 (0.5; 1.17) - Poverty level = 1.04 (0.99; 1.11) | Income, number of persons per household, mode of transportation, age, education level, poverty level |
| Adhikari et al. 2020  (USA) | 158 counties  158 counties that experienced at least one death attributed to confirmed COVID-19 | | SARS-CoV-2 Infection | SARS-CoV-2 Infection | NA | Socioeconomic determinants = RR (95% CI)  Less- poverty county   - Substantially White = Reference - Less diverse = 1.9 (1.4; 2.6) - More diverse = 3.2(2.3; 4.6) - Substantially non-White = 2.8(1.8; 4.4)   More- poverty county   - Substantially White = Reference - Less diverse = 3.8 (2.3; 6.1) - More diverse = 5.1(3.3; 8) - Substantially non-White = 7.1(5.1;12) | Income |
|  |  | | Mortality | NA | NA | Socioeconomic determinants = RR (95% CI)  Less- poverty county   - Substantially White = Reference - Less diverse = 1.8 (1.1- 3) - More diverse = 3.8(2.2- 6.7) - Substantially non-White = 2.6(1.1- 6.5)   More- poverty county   - Substantially White = Reference - Less diverse = 4.7 (2.2- 10) - More diverse = 4.9 (2.4- 9.7)   Substantially non-White = 9.3 (4.7-18.4) | Income |
| Alves et al.  2021  (Portugal) | 308 municipalities  Cross-sectional  Portuguese municipalities  Inclusionat four different points:  - April 1 2020 | | SARS-CoV-2 Infection | SARS-CoV-2 Infection | Socioeconomic determinants = OR (95% CI)  Unemployment:   - 1st tertile = 0.27 (0.12-0.59) - 2nd tertile = 0.36 (0.17-0.76) - 3rd tertile = Reference   Earning   - 1st tertile = 0.88 (0.44-1.76) - 2nd tertile = 0.52 (0.24-1.12) - 3rd tertile = Reference   Gini coefficient   - 1st tertile = 0.33 (0.15-0.73) - 2nd tertile = 0.63 (0.31-1.25) - 3rd tertile = Reference | Socioeconomic determinants = OR (95% CI)  Unemployment:   - 1st tertile = 0.36 (0.14-0.91) - 2nd tertile = 0.34 (0.15-0.81) - 3rd tertile = Reference   Earning   - 1st tertile = 2.40 (0.78-7.41) - 2nd tertile = 0.90 (0.34-2.41) - 3rd tertile = Reference   Gini coefficient   - 1st tertile = 0.50 (0.19-1.29) - 2nd tertile = 0.56 (0.25-1.24) - 3rd tertile = Reference | Population density, age, sex, socioeconomic variables (earning, GINI coefficient, unemployment), number of doctors |
|  | - May 1 2020 | | SARS-CoV-2 Infection | SARS-CoV-2 Infection | Unemployment:   - 1st tertile = 0.06 (0.01-0.44) - 2nd tertile =0.50 (0.20-1.23) - 3rd tertile = Reference   Earning   - 1st tertile = 0.35 (0.12-1.03) - 2nd tertile = 0.43 (0.16-1.19) - 3rd tertile = Reference   Gini coefficient   - 1st tertile = 0.72 (0.26-2.02) - 2nd tertile = 0.80 (0.30-2.18) - 3rd tertile = Reference | Unemployment:   - 1st tertile = 0.03 (0.00-0.41) - 2nd tertile = 0.44 (0.12-1.61) - 3rd tertile = Reference   Earning   - 1st tertile = 1.67 (0.29-9.76) - 2nd tertile = 0.84 (0.20-3.61) - 3rd tertile = Reference   Gini coefficient   - 1st tertile = 3.24 (0.36-28.84) - 2nd tertile = 2.78 (0.52-14.98) - 3rd tertile = Reference | Population density, age, sex, socioeconomic variables (earning, GINI coefficient, unemployment), number of doctors |
|  | - June 1 2020 | | SARS-CoV-2 Infection |  | Unemployment:   - 1st tertile = 0.71 (0.32-1.60) - 2nd tertile = 0.59 (0.25-1.37) - 3rd tertile = Reference   Earning   - 1st tertile = 0.32 (0.13-0.80) - 2nd tertile = 0.59 (0.27-1.29) - 3rd tertile = Reference   Gini coefficient   - 1st tertile = 0.76 (0.30-1.91) - 2nd tertile = 1.51 (0.67-3.41) - 3rd tertile = Reference | Unemployment:   - 1st tertile = 0.93 (0.31-2.80) - 2nd tertile = 0.58 (0.20-1.73) - 3rd tertile = Reference   Earning   - 1st tertile = 1.11 (0.33-3.73) - 2nd tertile = 1.96 (0.69-5.59) - 3rd tertile =   Gini coefficient   - 1st tertile = 1.17 (0.27-5.06) - 2nd tertile = 1.49 (0.51-4.37) - 3rd tertile = Reference | Population density, age, sex, socioeconomic variables (earning, GINI coefficient, unemployment), number of doctors |
|  | - July 1 2020 | | SARS-CoV-2 Infection |  | Unemployment:   - 1st tertile = 0.28 (0.13-0.61) - 2nd tertile = 0.55 (0.28-1.06 - 3rd tertile = Reference   Earning   - 1st tertile = 0.84 (0.43-1.63) - 2nd tertile = 0.57 (0.28-1.16) - 3rd tertile = Reference   Gini coefficient   - 1st tertile = 0.36 (0.17-0.77) - 2nd tertile = 0.72 (0.38-1.38) - 3rd tertile = Reference | Unemployment:   - 1st tertile = 0.40 (0.16-1.05) - 2nd tertile =0.65 (0.28-1.52) - 3rd tertile = Reference   Earning   - 1st tertile = 0.49 (0.18-1.30) - 2nd tertile =0.65 (0.29-1.47) - 3rd tertile = Reference   Gini coefficient   - 1st tertile = 4.71 (1.43-15.49) - 2nd tertile = 1.71 (0.61-4.78) - 3rd tertile = Reference | Population density, age, sex, socioeconomic variables (earning, GINI coefficient, unemployment), number of doctors |
| Li et al. 2020  (USA) | 661 counties  April 14 2020 | | SARS-CoV-2 Infection | NA | Socioeconomic determinants = OR (95% CI)  Race   - White = Reference - Black = 1.03 (1.02- 1.06) | Socioeconomic determinants = OR (95% CI)  Race   - White = Reference - Black = 1.22 (1.09- 1.40) | Macroeconomic and covid specific variables, county demographic and environmental factors, medical comorbidities and access to healthcare |
|  |  | | Mortality | NA | Regression coefficients  Race   - White = Reference   Black = -0.01(-0.63-0.61) | Socioeconomic determinants = Regression coefficients  Race   - White = Reference   Black = 0.35 (0.09-0.61) | Macroeconomic and covid specific variables, county demographic and environmental factors, medical comorbidities and access to healthcare |
| Temkin Greener et al. 2020  (USA) | N = 3994 ALs  Residents of Assisted Living (AL) communities  March 2020 - May 29 2020 | | SARS-CoV-2 Infection | 2 542 cases (96.0%) | NA | Socioeconomic determinants = OR (95% CI)   - Black and Hispanic = 0.94 (0.87- 1.03) - White = Reference | Assited Living Residens variables (age, sex, insurance, number of residents, comorbidities: dementia, COPD, asthma, CHF, obesity, HTA, diabetes), COVID-19 cases/1000 pop, state-level fixed effect |
|  |  | | Mortality | 675 deaths (86.9%) | NA | Socioeconomic determinants = OR   - Black and Hispanic = 0.948 (0.83- 1.08) - White = Reference | Assisted Living Residens variables (age, sex, insurance, number of residents, comorbidities: dementia, COPD, asthma, CHF, obesity, HTA, diabetes), COVID-19 deaths/1000 pop, state-level fixed effect |
| Abedi et al. 2020  (USA) | N = 102 178 117  Community-based – County level  April 9 2020 | | SARS-CoV-2 Infection | COVID-19 infection rate per one million mean (± SD), 912.20 ± 1034.26 | NA | Socioeconomic determinants = Regression coefficients (95% CI)  Ethnicity   - Black male = 0.354 (0.204-0.505) - White male = - 0.439 (0.584 - -0.295) - Asian male = 0.317 (0.202 - 0.435) - Hispanic male = 0.463 (0.307-0.618) - Black female = 0.471 (0.32 - 0.621) - White female = - 0.409 (-0.554- -0.264) - Asian female = 0.318 (0.201 - 0.435) - Hispanic female = 0.491 (0.341 - 0.642)   Median income = 0.364 (0.247-0.481)  High school = 0.076 (-0.046-0.198)  Bachelor degree = 0.299 (0.191-0.408)  Medicaid = -0.15(-0.261 - -0.04)  Poverty = -0.167(-0.284 - 0.05) | State variables (Poverty, education, income) |
|  |  | | Mortality | COVID-19-related death (mean, 4.13%± 2.70%- median, 3.40- IQR, 2.22–5.61) | NA | Socioeconomic determinants = Regression coefficients (95% CI)  Ethnicity   - Black male = -0.082(-0.269-0.104) - White male = 0.143(-0.027-0.313) - Asian male = -0.227(-0.365 – -0.088) - Hispanic male = -0.225(-0.402- -0.048) - Black female = -0.064(-0.236-0.018) - White female = 0.117(-0.051-0.285) - Asian female = -0.267(-0.41- -0.124) - Hispanic female = -0.21(-0.381- - 0.039)   Median income = -0.265(-0.408- - 0.122)  High school =0.134(-0.294-0.026)  Bachelor degree = -0.246(-0.388- -0.103)  Medicaid = 0.171(0.034-0.307)  Poverty = 0.148(-0.003-0.299) | State variables (Poverty, education, income) |
| Biggs et al. 2020  (USA) | N = 1105 census tracts  Louisiana census tracts.  March 9 2020 – August 24 2020 | | SARS-CoV-2 Infection | NA | Socioeconomic determinants = RR (95% CI)   - Overall Social Vulnerability Index = 1.52 (1.4 -1.65) - Theme 1 Socioeconomic = 1.32 (1.21-1.44) - Theme 2-House Composition & Disability = 1.27 (1.17-1.39) - Theme 3- Minority & Language = 1.6 (1.48-1.7) - Theme 4- Housing & Transportation = 1.35 (1.24-1.46) | Socioeconomic determinants = RR (95% CI)   - Overall Social Vulnerability Index = 1.52 (1.41-1.65) - Theme 1 Socioeconomic = 0.97 (0.87-1.09) - Theme 2-House Composition & Disability = 1.24 (1.12-1.36) - Theme 3- Minority & Language = 1.36 (1.24-1.49) - Theme 4- Housing & Transportation = 1.21 (1.1-1.32) | Model 1 (Over SVI): Population density  Model 2 (4 themes) : Population density, socioeconomic theme, House composition and disability, minority and language, housing and transportation |
| Chen et al. 2020  (USA) | 3142 counties (322 903 3030 population)  USA county  As of May 5 2020 | | SARS-CoV-2 Infection | 1 189 798 cases | NA | Socioeconomic determinants = RR (95% CI)  % poverty (categories)   - 0-4.9% = Reference - 5-9.9% = 1.36 (1.34 - 1.39) - 10-14.9% = 0.87 (0.85 - 0.88) - 15-19.9% = 0.89 (0.87 - 0.90) - 20-100% = 1.41 (1.39 - 1.44)   Index of Concentration at the Extremes (high income White households versus low income Black households)   - Q1: (-0.522,0.114) = 0.77 (0.76 - 0.77) - Q2: (0.114,0.159) = 0.54 (0.54- 0.55) - Q3: (0.159,0.205) = 0.41 (0.41 - 0.41) - Q4: (0.205 -0.283) = 0.53 (0.52 - 0.53) - Q5: (0.283,0.536) =Reference   % crowding (quintiles)   - Q1: (0%, 1.47%) = Reference - Q2: (1.47%, 2.12%) = 1.28 (1.27 - 1.29) - Q3: (2.12%, 3.06%) = 1.92 (1.91- 1.93) - Q4: (3.06%, 4.91%) = 1.83 (1.82- 1.84) - Q5: (4.91%, 49.3%) = 2.14 (2.12 - 2.15)   % percent population of color   - Q1: (0%, 17.2%) = Reference - Q2 (17.2%, 30.2%) = 2.05 (2.04 - 2.07) - Q3(30.2%, 44.3%) = 3.10 (3.07 - 3.12) - Q4(44.3%, 61%) = 3.23 (3.21-3.26) - Q5(61%, 100%) = 3.47 (3.44 - 3.49) | Age and sex |
|  |  | | Mortality | 68 656 Deaths | NA | Socioeconomic determinants = RR (95% CI)  % poverty (categories)   - 0-4.9%   = Reference   - 5-9.9% = 1.06 (0.99, 1.12) - 10-14.9% = 0.73 (0.69, 0.78) - 15-19.9% = 0.66 (0.62, 0.70) - 20-100% = 1.72 (1.61, 1.83)   Index of Concentration at the Extremes (high income White households versus low income Black households)   - Q1: (-0.522,0.114) = 1.04 (1.02, 1.06) - Q2: (0.114,0.159)   = 0.55 (0.53, 0.56)   - Q3: (0.159,0.205)   = 0.34 (0.33, 0.35)   - Q4: (0.205 -0.283)   = 0.52 (0.50, 0.53)   - Q5: (0.283,0.536) =   Reference  % crowding (quintiles)   - Q1: (0%, 1.47%) = Reference - Q2: (1.47%, 2.12%)   = 1.17 (1.14, 1.20)   - Q3: (2.12%, 3.06%) - = 1.66 (1.62, 1.71)Q4: (3.06%, 4.91%) = 1.36 (1.32, 1.40) - Q5: (4.91%, 49.3%) = 2.58 (2.52, 2.65)   % percent population of color   - Q1: (0%, 17.2%)   = Reference   - Q2 (17.2%, 30.2%)   = 2.20 (2.13, 2.28)   - Q3(30.2%, 44.3%)   = 3.07 (2.97, 3.17)   - Q4(44.3%, 61%)   = 3.34 (3.23, 3.45)  Q5(61%, 100%) = 4.94 (4.78, 5.09) | Age and sex |
| De Souza et al. 2020  (Brazil) | N=2496 municipalities  May 6 2020 | | SARS-CoV-2 Infection | N=125 186 (0.07%)  MHDI (Municipal Human Development Index,)   - Very low (0-0.499) N=289 (0.23%) - Low (0.500-0.599) N=4031(3.22%) - Medium (0.600-0.699) N=13021 (10.40%) - High (0.700-0.799) N=72544 (57.95%) - Very high (0.800-1) N=35301 (28.20%)   SVI (Social Vulnerability Index.)   - Very low (0-0.200) N=7428 (5.93%) - Low (0.200-0.300) N=54736 (43.72%) - Medium (0.300-0.400) N=49528 (39.56%) - High (0.400-0.500) N=8530 (6.81%) - Very high (0.500-1) N=4964 (3.97%)   Population size   - ≤10,000 inhabitants N=1296 (1.04%) - 10,001 to 20,000 N=2568 (2.05%) - 20,001 to 50,000 N=7075 (5.65%) - 50,001 to 100,000 N=6988 (5.58%) - >100,000 N=107259 (85.68%) |  | Socioeconomic determinants=correlation coefficient (P-value)  SVI=157.70 (<0.001)  MDHI=297.64 (<0.001) | SVI and MDHI variables |
| Demenech et al. 2020  (Brazil) | N= 27 Federative Units  April 21 2020-July 7 2020 | | SARS-CoV-2 Infection | NA | NA | Socioeconomic determinants= correlation coefficient (SE) (P-value)  Gini coefficient = 10406.8 (0.003)  Population density = -0.42 (0.682) | Population density |
|  |  | | Mortality | NA | NA | Socioeconomic determinants= correlation coefficient (p-value)  Gini coefficient = 319.122 (< 0.001)  Population density = -0.01 (0.916) | Population density |
| Sugg et al. 2021  (USA) | 13709 nursing homes  Nursing homes  Inclusiontill June 30 2020 | | SARS-CoV-2 Infection | Average of 11.32 confirmed cases per nursing home | NA | Socioeconomic determinants= RR (95% CI)  % Total population: African American = 1.27(1.27–1.27)  Average household size = 1.26 (1.26–1.26)  Per capita income = 2.48 (2.48–2.49)  % Civilian population in labor force 16 years and over Unemployed = 1.32 (1.32–1.32)  Population per sq. mile = 1.12(1.12–1.12) | Facility-based factors including: LPN staffing levels, total staff, number of fines for 2020  County-level factors including:  % Civilian population in labor force 16 years and over: unemployed, average household size, per capita income, population per sq. mile (2010), % Total population: African American,), COVID-19 rate (County) |
| Yang et al. 2021  (Hong Kong) | Tertiary Planning Unit Level  N = 4811 COVID-19 cases  Wave 1&2:  N = 1038  January 18 2020 – April 30 2020  Wave 3:  N = 3773  May 1 2020 – August 31 2020 | | SARS-CoV-2 Infection | N = 4811 COVID-19 cases  Wave 1&2:  N = 1038 (14%)  January 18 2020 – April 30 2020  Wave 3:  N = 3773 (86%)  May 1 2020 – August 31 2020 | NA | Socioeconomic determinants = IRR (95% CI)  Wave 1&2 :  Tertiary education or above (per 10% increase) = 1.44 (0.36-5.76)  Executives and professionals (per 10% increase) = 3.69 (1.02- 13.33)  Median monthly rent (per HKD 10 000 increase) = 1.09 (1.01- 1.18)  Wave 3:  Tertiary education or above (per 10% increase) = 0.60 (0.06-5.56)  Executives and professionals (per 10% increase) = .16 (0.03- 0.99)  Median monthly rent (per HKD 10 000 increase) = 0.96 (0.87-1.06) | Median monthly rent, proportion with tertiary education, proportion of executives and professionals, age |
| Figueroa et al. 2020  (USA) | 351 towns  Massachusetts cities  January 1 2020 – May 6 2020 | | SARS-CoV-2 Infection | NA | Socioeconomic determinants = Regression coefficients (95% CI)  Ethnicity   - Proportion of Black non-Latino population = 312.3 (241.9-383.0) - Proportion of Latino population = 258.2 (217- 300-3) - White non-Latino = Reference - Proportion of other non-Latino population = 86.5(7.2- 169.9)   Median income = -87.7 (−109.6- 64.6)  Mean household size = 669.4(344- 994.8)  Proportion with < HS education = 611.9 (524- 699.84)  Proportion of foreign-born noncitizens= 749.3 (658.8- 839.8) | Socioeconomic determinants = Regression coefficients (95% CI)  Ethnicity   - Proportion of Black non-Latino population = 307.2 (219.6-394.7) - Proportion of Latino population = 50.4 (−19.0-119.8) - White non-Latino = Reference - Proportion of other non-Latino population = -216.4(−321.4 - - 111.3)   Median income = -23.7 (-69.8- 22.7)  Mean household size = 886.2 (494.3-1278.1)  Proportion with < HS education = 57.7 (−115.1-230.5)  Proportion of foreign-born noncitizens= 791.7 (587.9- 851.6) | Proportion of people by race/ethnicity, foreign-born noncitizens, essential workers, older than age sixty, and with less than a high school education; number of people for household size; dollars for median income; and total number of people in the city or town |
| Figueroa et al. 2021  (USA) | USA counties  January 1 2020 – September 12 2020 | | SARS-CoV-2 Infection | NA | Socioeconomic determinants = Regression coefficients (95% CI)  Ethnicity   - Proportion of Black non-Latino population = 324.7 (298.0 - 351.4) - Proportion of Latino population = 293.5 (274.8 - 312.2) - White non-Latino = Reference - Proportion of other non-Latino population = -81(-125.7 - -36.2)   Median income = -108.6(-129.4 - -87.7)  Average household size = -600.3 (-673.8 - -526.9)  Proportion with < HS education = 990.8 (933.2- 1048.5)  Proportion of foreign-born noncitizens= 903.4 (843.4- 963.4) | Socioeconomic determinants = Regression coefficients (95% CI)  Ethnicity   - Proportion of Black non-Latino population = 278.1 (252.2 - 303.9) - Proportion of Latino population = 23.8 (-13.5 - 61.2) - White non-Latino = Reference - Proportion of other non-Latino population = -268.9 (-310.5 - -227.3)   Median income = -112.7 (-143.1 - -82.2)  Average household size = -1117.3 (934.4 - 1300.2)  Proportion with < HS education = 208.8 (106.5 - 311.1)  Proportion of foreign-born noncitizens= 803.7 (682.8 - 924.6) | Proportion of people by race/ethnicity, total population, average household size, median household income, the proportions of people who were age 60 years or older, proportion of adults who were employed as essential workers, proportion of adults who were foreign-born non-citizens, proportion of adults who completed less than a high school degree, and proportion of adults who commute Usingpublic transportation. |
|  |  | | Mortality | NA | Socioeconomic determinants = Regression coefficients (95% CI)  Ethnicity   - Proportion of Black non-Latino population = 14.5 (12.9 - 16.0) - Proportion of Latino population = 7.6 (6.5 - 8.8) - White non-Latino = Reference - Proportion of other non-Latino population = 5.7 (3.1 - 8.2)   Median income = 1.1(-0.1 - 2.3)  Average household size = -600.3 (22.3 - 37.6)  Proportion with < HS education = 31.5 (27.8 - 35.1)  Proportion of foreign-born noncitizens= 40.4 (36.8 - 44) | Socioeconomic determinants = Regression coefficients (95% CI)  Ethnicity   - Proportion of Black non-Latino population = 9.3 (8.0 - 10.6) - Proportion of Latino population = 0-4 (-1.5 - 2.4) - White non-Latino = Reference - Proportion of other non-Latino population = -11-9 (-14.0 - -9.7)   Median income = 1.6 (0.0 - 3.2)  Average household size = 56.4 (46.9 - 66.0  Proportion with < HS education = 13.8 (8.4 - 19.1)  Proportion of foreign-born noncitizens= 0.9 (-5.4 - 7.2) | Proportion of people by race/ethnicity, total population, average household size, median household income, the proportions of people who were age 60 years or older, proportion of adults who were employed as essential workers, proportion of adults who were foreign-born non-citizens, proportion of adults who completed less than a high school degree, and proportion of adults who commute Usingpublic transportation. |
| Garcias et al. 2021  (Spain) | 17 Spanish regions  Inclusionas of May 23 2020 | | SARS-CoV-2 Infection | NA | NA | Socioeconomic determinants = Regression coefficients (p-value)  Urban population = -0.0007 (< 0.1)  Log GDP per capita = 0.041 (<0.1) | Nursing homes beds,  % Nursing homes >100, urban population, aeroplane passengers, island region, log GDP per capita, % >65y population |
|  |  | | Mortality | NA | NA | Socioeconomic determinants = Regression coefficients (p-value)  Urban population = − 0.01 (< 0.1)  Log GDP per capita = 3.215 (<0.01) | Log Public Health Expenditure, nursing homes beds, % nursing homes >100,Urban population,  doctors, hospital Beds, aeroplane passengers, island region,  log GDP per capita |
| Hawkins D. et al. 2020  USA | Residents of Massachusetts (MA)  January 1 2020- June 10 2020 | | SARS-CoV-2 Infection | 87 256 COVID-19 cases  1 584.8 COVID-19 cases per 100 000 | NA | Socioeconomic determinants = RR(95% CI)  Percentage of residents in poverty   - First quartile = Reference - Second quartile= 1.26 (1.22- 1.30) - Third quartile = 1.39 (1.35- 1.43) - Fourth quartile = 1.96 (1.91- 2.02)   Median income   - Fourth quartile = Reference - Third quartile = 1.41 (1.37- 1.45) - Second quartile = 1.57 (1.53- 1.61) - First quartile = 2.19 (2.14- 2.25)   Percentage of residents who rented   - First quartile = Reference - Second quartile = 1.20 (1.16- 1.24) - Third quartile = 1.44 (1.40- 1.48) - Fourth quartile = 1.56 (1.52- 1.61)   Percentage of residents who were uninsured   - First quartile= Reference - Second quartile= 1.09 (1.05- 1.12) - Third quartile= 1.55 (1.50- 1.60) - Fourth quartile= 1.87 (1.82- 1.93)   Unemployment rate   - First quartile = Reference - Second quartile= 1.15 (1.12- 1.18) - Third quartile= 1.74 (1.70- 1.79) - Fourth quartile= 1.54 (1.50- 1.58) | Social variables (poverty, median income, employment in the health care and transportation industries and healthcare support and service occupations, rented accommodations, unemployment, and lack of insurance.);  median age of city |
| Li et al. 2020  (USA) | N = 12 576  Nursing homes nationally  May 25 2020 – May 31 2020 | | SARS-CoV-2 Infection | 0.9 ± 4.7 cOVID-19 confirmed cases among residents  Mean ± standard deviation   - Low proportion group = 0.4 ± 2.5 - Medium-proportion group = 0.7 ± 4.0) - Medium-high–proportion group = 0.9 ± 5.2 - High-proportion group = 1.5 ± 6.3 | NA | Socioeconomic determinants = RR(95% CI)  Proportions of racial/ethnic minority residents   - Low proportion group = Reference - Medium-proportion group = 1.25 (1.03–1.51) - Medium-high–proportion group = 1.44 (1.14-1.82) - High-proportion group = 1.76 (1.38-2.25) | Nursing home (i.e. NH with medium proportion of racial/ethnic minority residents / medium-high/ high, number of certified beds, type of ownership, % of Medicare and Medicaid residents …) and county-level covariates (i.e. county population size in 100k, county % of older population (65+), county median household income in $1000, county % of population with high school graduation, county-level competition for nursing home residents) and states covariates |
| Kolin et al.  2020  (UK) | N = 397 064  Census zone  March 16 2020 – May 18 2020 | | SARS-CoV-2 Infection | 968 COVID-19 cases (0;24%) | NA | Socioeconomic determinants = RR (95% CI)  Economic activity   - Individuals self-employed = 0.84 (0.78-0.90) - Individuals unemployed = 1.19 (1.12-1.25) - Individuals retired = 0.91 (0.85-0.975)   Ethnic group   - White Individuals = 0.90 (0.85-0.96) - Multiple Ethnicities Individuals = 1.14 (1.08-1.20) - Black Individuals = 1.12 (1.07-1.17)   Household composition   - Individuals Lone Parents = 1.18 (1.12-1.25) - Individuals in Other Household Type with Dependent Children = 1.08 (1.02-1.13)   Living arrangements   - Individuals Living Not in Couple = 1.16 (1.09-1.23) - Individuals Separated but Married = 1.15 (1.08-1.22)   Qualifications   - Individuals with No Qualification = 1.14 (1.07-1.32) - Individuals with Level 2 Qualification = 0.95 (0.90-1.02) - Individuals with Level 4 Qualification = 0.89 (0.83-0.95)   Tenure   - Individuals Own Home = 0.83 (0.78-0.88) - Individuals Socially Rent = 1.18 (1.12-1.24) - Individuals Rent from Other = 1.13 (1.07-1.19) | Age, sex, body-mass index, systolic blood pressure, and race |
| Hawkins R. et al. 2020  (USA) | 3143 counties  May 2, 2020 | | SARS-CoV-2 Infection | 1,089,999 cases | NA | - Housing vacancy rate   1.01 (1-1.02)   - Median income ratio 1.01(1.01-1.01) - Poverty rate 0.98(0.96-0.99) - % change in employment 1(1-1) - % change in establishment   1(0.99-1.01)   - % of adults not working 0.99(0.98-1) - % of adults without high school degree 1.1(1.09-1.11) - % population in distressed zipcodes 1(1-1) - % of population Black 1.03(1.02-1.04) - % of uninsured population aged younger than 65 1.01(1-1.02) | Age, comorbidities (chronic kidney disease, COPD, heart disease, obesity, diabetes), days since first case, Housing vacancy rate Median income ratio, Poverty rate % change in employment % change in establishments % of adults not working % of adults w/o a high school degree % population in distressed zip codes % of population aged older than 65 years % of population Black % of uninsured population aged younger than 65 years |
|  |  | | Mortality | 1,089,999 cases and 62,298 deaths in 3127 counties  Case fatality rate 5.7% |  | - Housing vacancy rate   0.98 (0.95-1.01)   - Median income ratio 1.01 (1-1.02) - Poverty rate 1 (0.97-1.02) - % change in employment 0.99 (0.99-1) - % change in establishment   1 (0.99-1.02)   - % of adults not working 0.98 (0.96-1) - % of adults without high school degree 1.08 (1.05-1.11) - % population in distressed zip codes   1(0.99-1)   - % of population Black 1.03 (1.02-1.05) - % of uninsured population aged younger than 65 0.99 (0.96-1.01) | Age, comorbidities (chronic kidney disease, COPD, heart disease, obesity, diabetes), days since first case, Housing vacancy rate Median income ratio, Poverty rate % change in employment % change in establishments % of adults not working % of adults w/o a high school degree % population in distressed zip codes % of population aged older than 65 years % of population Black % of uninsured population aged younger than 65 years |
| Karmakar et al. 2021  (USA) | 3137 counties  USA counties  20 January 2020- July 29 2020 | | SARS-CoV-2 Infection | 289 283 COVID-19 cases | NA | Socioeconomic determinants = IRR (95% CI)   - Poverty rate = 1.03 (1.03-1.04) - Unemployment rate (%) = 1.04 (1.03-1.06) - Per capita income = 0.98 (0.97-0.98) - Education = 1.07 (1.06-1.07) - Adults without health insurance (%) = 1.09 (1.08-1.10) - Gini income inequality index = 1.03 (1.02-1.04) - Racial/ethnic minorities and language, %   Any racial/ethnic minorityd African = 1.03 (1.02-1.03)  American = 1.02 (1.02-1.02)  Hispanic or Latinx = 1.02 (1.01-1.02)  American Indian or Alaskan Native = 1.01 (1.01-1.02)  Asian = 1.03 (1.01-1.05) | Population density, urbanicity, and COVID-19 testing rate |
|  |  | | Mortality | 147 074 COVID-19 deaths | NA | Socioeconomic determinants = IR (95% CI)   - Poverty rate = 1.05 (1.04-1.05) - Unemployment rate (%) = 1.07 (1.05-1.09) - Per capita income = 0.98 (0.97-0.99) - Education = 1.06 (1.05-1.07) - Adults without health insurance (%) = 1.07 (1.05-1.08) - Gini income inequality index = 1.05 (1.04-1.07) - Racial/ethnic minorities and language, %   Any racial/ethnic minority = 1.03 (1.02-1.03)  African = 1.02 (1.02-1.03)  American = 1.02 (1.02-1.02)  Hispanic or Latinx = 1.02 (1.01-1.02)  American Indian or Alaskan Native = 1.02 (1.02-1.03)  Asian = 1.01 (0.99-1.03 | Population density, urbanicity, and COVID-19 testing rate |
| Loomba et al. 2021  (USA) | Cross-sectional  Community State level  March 14, 2020 - April 30, 2020 | | SARS-CoV-2 Infection | 1 090 500 COVID-19 cases | Socioeconomic determinants = Coefficients (95% CI)  Race White = -4715.4(-11765.1-2334.2) Black =7312.70(-2051.3-16676.9) Native American = 23515(-54374.7-7342.9) Asian = 6237.80(10201.4-22677.1) Islander = -30422.10(-94071.5-33227.3) Other race = 38638.70(8221.4-69.056.1) Multiple race = -13133.50(-41037.2-14770.1)  Insurance Uninsured = -298.6(-544.2, -52.9)  Average household income 0.1(0.1-0.2)  High school grad percent = -82.5(-381.6- 216.5)  Per capita health care spending = 1(0.3-1.7)  Public transportation volume = 0.1(0.1-0.12  State popularity density = 8.9(6.5-11.2) | Socioeconomic determinants = Coefficients (p-value)  Race White = -0.05 (0.4) Black =-0.08(0.16) Native American = 0.02 ( 0.73) Asian = -0.11(0.14) Islander = -0.07 (0.24)  Other race = -0.06 (0.39) Multiple race = -0.08 ( 0.16)  Insurance Uninsured = 0.11 (0.13)  Average household income = -0.08 (0.28)  High school grad percent = -0.05(0.44)  Per capita health care spending = -0.04 (0.58)  Public transportation volume = 0.57 (<0.01)  State popularity density = 0.64 (<0.01) | Age, gender, comorbidities, obesity ,environmental covariates, drug use , Number of residents in nursing home facilities Number of inmates in prisons Tourism ranking 2018 (lower ranking means more tourists), race |
|  |  | | Mortality | 63 642 CVOID-19 deaths | Socioeconomic determinants = Coefficients (95% CI)  Race White = -1.6(-5.8-2.4) Black = 4.3(-1- 9.7) Native American = -16.6(-34.2-0.9) Asian = 1(-8.5- 10.6) Islander = -57.2(-57.2-16.1) Other race = 9.4(-9- 28) Multiple race = -4.6(-20.8- 11.6)  Insurance Uninsured -0.1(-0.3- -0.1)  Average household income 0.1(-0.1- 0.1)  High school grad percent -0.1(-0.2- 0.1)  Per capita health care spending 0.1(0.1-0.1)  Public transportation volume 0.1(0.1-0.2)  State popularity density  0.1(0.1-0.2) | Socioeconomic determinants = Coefficients (p-value)  Race White = -0.18 (0.18) Black =0.21 (0.13) Native American = -0.22 ( 0.09) Asian = 0.16 (0.28) Islander = -0.23 (0.09)  Other race = -0.02 (0.84) Multiple race = 0.1 ( 0.45)  Insurance Uninsured = -0.2 (0.2)  Average household income = 0.2 (0.16)  High school grad percent = -0.03 (0.81)  Per capita health care spending = 0.14 (0.37)  Public transportation volume = 0.11 (0.43)  State popularity density = 0.24 (0.07) | Age, gender, comorbidities, obesity, environmental covariates, drug use , Number of residents in nursing home facilities Number of inmates in prisons Tourism ranking 2018 (lower ranking means more tourists), race |
| Macchia et al. 2021  (Argentina) | N = 114 052  Community  January 31 – July 7 2020 | | SARS-CoV-2 Infection | 39 039 COVID-19 cases (34.22%) | Socioeconomic determinants = IRR (95% CI)  Socioeconomic status (SES) quintiles   - 1 = Reference - 2 = 1.358(1.259-1.465) - 3 = 1.611(1.492-1.741) - 4 = 1.865 (1.729-2.012) - 5 = 2.942(2.738-3.162)   Slums = 14.375(13.413 – 15.407) | NA | NA |
| Mehta et al. 2021  (USA) | Cohort  N = 482 323  Nursing home residents  April 1, 2020 - September 30, 2020 | | SARS-CoV-2 Infection | 131 119 COVID-19 infections (27.18%)   - Black N = 23 698 - Asian N = 3 899 - Hispanic or Latino N = 10 346 - Other N = 644 - White = 98 532 |  | Socioeconomic determinants = HR (95% CI)   - White = Reference - Black = 1.04(1.03-1.06) - Asian = 1.07(1.03-1.11) - Hispanic or Latino =1.07(1;05-1;1) - Other = 1.02(0.94-1.11) | Age, sex, Cognitive function, functional impairment, use of catheter, prognosis, comorbidities |
| Khazanchi et al. 2020  (USA) | 2754 counties  USA counties  April 19 2020 | | SARS-CoV-2 Infection |  |  | Socioeconomic determinants = IRR (95% CI)  Social vulnerability index =1.63(1.49-1.78)  Socioeconomic status = 1.42(1.26-1.6)  Household composition and disability = 0.85(0.73-0.99)  Minority status and language =4.94(3.91-6.24)  Housing type and transportation =1.52(1.35-1.72) | Socioeconomic status, household compositions and disability, minority status and language, Housing type and transportation, social vulnerability index |
|  |  | | Mortality |  |  | Socioeconomic determinants = IRR (95% CI)  Social vulnerability index =1.73(1.55-1.93)  Socioeconomic status = 1.71(1.47-1.98)  Household composition and disability = 1.1(0.93-1.3)  Minority status and language =4.74(3.55-6.32)  Housing type and transportation =1.32(1.14-1.53) | Socioeconomic status, household compositions and disability, minority status and language, Housing type and transportation, social vulnerability index |
| Palacio et al. 2021  (USA) | N = 2 834 352  Community  Inclusion on July 24 2020 | | SARS-CoV-2 Infection | 97 594 COVID-19 infections | NA | Socioeconomic determinants = IRR (95% CI)   - Quartile income 50 194-61 897$ = 1.16 (1.13-1.19) - Quartile income 41 541-49 239$ = 1.46 (1.43-1.5) - Quartile income 14 999-40 039$ = 1.89 (1.85-1.93) - Tertile of Self-reported Financial Strain (34- 52$) = 1.17(1.15-1.19) - Tertile of Selfreported Financial Strain = (53-100$) = 1.51(1.49-1.54) | Population and household size |
| Gaudart et al.  2021  (France) | 96 departments / in-hospital patients  March 19 2020 - May 11 2020 | | SARS-CoV-2 Infection | 100 988 COVID-19 cases  16 597 ICU admissions  17 062 deaths | Socioeconomic determinants = SIR (95% CI)  High median strandard of living = Reference  High rate of social assistance = 1.07 (0·87–1·32)  High poverty and unemployment ratio = 0.97 (0.68–1.38) | NA | NA |
| Lewis et al. 2020  (USA) | 99 Utah small statistical areas  General population (community  July 3 2020 – July 9 2020 | | SARS-CoV-2 Infection | 28 139 COVID-19 infections   - Very low N = 3 119 - Low N = 5 585 - Average N = 4 943 - High N = 6324 - Very high N = 8 177 | Socioeconomic determinants = OR (95% CI)  Level of deprivation   - Very low = Reference - Low = 1.23 (1.17–1.29) - Average = 1.49 (1.42–1.56) - High = 2.06 (1.98–2.15) - Very high = 3.00 (2.88–3.13) | Socioeconomic determinants = OR (95% CI)  Level of deprivation   - Very low = Reference - Low = 1.23 (1.19–1.29) - Average = 1.49 (1.43–1.56) - High = 2.08 (1.99–2.17) - Very high = 3.11 (2.98–3.24) | Age |
| Das et al. 2020  (India) | 155 Eelctoral wards  General population  May 15 2020 – May 21 2020 | | SARS-CoV-2 Infection | NA | NA | Socioeconomic determinants = Regression coefficients (p-value)  Poor Housing Condition = 0.322 (<0.001)  Lack of Household Amenities and Services = 0.26 (<0.03)  Low Asset Possession = 0.285 (<0.031) | Poor Housing Condition, Lack of Household Amenities and Services, Low Asset Possession, Poor WaSH Services, Gender Disparity |
| Li et al. 2020  (USA) | 215 Nursing homes with at least once case  Residents of nursing homes (NHs)  Inclusionas of April 4 2020 | | SARS-CoV-2 Infection | 1 918.8 infections (87–6,816) | NA | Socioeconomic determinants = OR (95% CI)  NHs concentrated by Medicaid residents = 1.16 (1.02-1.32)  NHs concentrated by racial/ethnic minority residents = 1.15(1.03-1.29) | Nursing home staffing, overall quality of care , concentration of Medicaid residents , and concentration of racial and ethnic minority residents |
| Alipio et al. 2020  (Philippine) | 17 regions  General population  Inclusionas of April 7 2020 | | SARS-CoV-2 Infection | NA | NA | Socioeconomic determinants = Regression coefficients (p-value)  Income = -3.99(<0.001) | Subsistence incidence |
| Ramirez-Aldana 2020  (Iran) | 31 provinces  General population  February 419 2020-March 18 2020 | | SARS-CoV-2 Infection | NA | NA | Socioeconomic determinants = Regression coefficients (p-value)  Urban population = 0.026 (0.008)  Literacy = -0.103 (<0.001) | Urban population, population aged>60 years old, literacy, average temperature, GDP, physician distribution, Transportation Efficiency Index |
| Ginsburgh et al. 2021  (France) | 94 French continental departments  Hospital setting  May 13 2020 – September 3 2020 | | SARS-CoV-2 Infection | Mean (range)  COVID-19 infections 1.78.79 per 100 000 (43.03-705.80 per 100 000) | NA | Socioeconomic determinants = Regression coefficients (p-value)  Income = -0.509 (0.319)  Household size = -0.093 (0.084)  Rural dummy = 0.451 (0.403)  Gini index = 0.039 (0.014) | Gini,log income, percent over 60, GPs density, rural dummy, household size, tests in population |
|  |  | | Mortality | Mean (range)  COVID-19 deaths 22.16 per 100 00 (0.66-114.19 per 100 000) | NA | Socioeconomic determinants = Regression coefficients (p-value)   - Income = -0.115 (0.89) - Household size = 0.67(0.78) - Rural dummy = -0.076(0.179)   Gini index = 0.08(0.028) | Gini,log income, percent over 60, GPs density, rural dummy, household size, tests in population |
| Mollalo et al. 2020  (USA) | General population / County-level  January 22 2020 – April 9 2020 | | SARS-CoV-2 Infection | NA | NA | Socioeconomic determinants = Regression coefficients (p-value)  Income inequality = 0.172 (0)  Median household inequality = 0.183 (0)  % of Black females = 0.064 (0) | Income inequality, median household income,% of nurse practitioners, % Black females |
| Ehlert et al. 2020  (Germany) | 401 administrative districts  General population  Inclusionuntil June 16 2020 | | SARS-CoV-2 Infection | NA | NA | Socioeconomic determinants = Regression coefficients (p-value)  Household income = 0.22 (> 0.1)  Unemployment rate = -22.35 (<0.05)  Rural regions = 28.28 (> 0.1)  Academics = 28.77 (<0.05) | Economic, demographic and regional characteristics |
|  |  | | Mortality | NA | NA | Socioeconomic determinants = Regression coefficients (p-value)   - Household income = 0(> 0.1) - Unemployment rate = -0.28 (>0.1) - Rural regions = 3.2 (> 0.1) - Academics = 1.13 (<0.1) | Economic, demographic and regional characteristics |
| Karaye et al. 2020  (USA) | 48 states  General population  Inclusionas of May 12 2020 | | SARS-CoV-2 Infection | 1 320 909 COVID-19 infections | NA | Socioeconomic determinants = Regression coefficients (p-value)  Overall SVI = 1.65(0.03)  Socioeconomic status (SES) = 0.61(0.05)  Household composition and disability= 0.72 (<0.001)  Minority status and language = 6.69 (<0.001)  Housing and transportation =0.69 (<0.001) | Population size, population density, number of people tested, average daily sunlight, precipitation, air temperature, heat index, and fine PM (PM2.5) |
| Khanijahani et al. 2020  (USA) | 3142 USA counties  General population  Inclusionuntil November 2 2020 | | SARS-CoV-2 Infection | NA | NA | Socioeconomic determinants = Regression coefficients (95%CI)  % Hispanic = 0.68 (0.48-0.87)  % Black = 0.69 (0.53-0.85)  Median household income = 0.06 (-0.14 – 0.260  Average household size = -4.55 (-12.83 – 4.51)  Unemployment rate = -2.37 (-3.83 – 0.87) | County-level socioeconomic variables |
|  |  | | Mortality | NA | NA | Socioeconomic determinants = Regression coefficients (95%CI)  % Hispanic = 0.65 (0.1-1.2)  % Black = 1.57 (1.11-2.03)  Median household income = 0.89 (0.35 – 1.42)  Average household size = 7.84 (-15.93– 38.48)  Unemployment rate = 1.22 (-0.15 – 2.61) | County-level socioeconomic variables |
| White et al. 2020  (USA) | 341 nursing facilities  Inclusionas of April 21 2020 | | SARS-CoV-2 Infection | NA | NA | Socioeconomic determinants = Marginal effect (95%CI)  % Black = 0.199-0.003 – 0.386) | Facility size, date of first county case, and whether the skilled nursing facilities (SNF) was universally tested |
| Zhang et al. 2020  (USA) | 1 624 counties  General population  Inclusionas of May 1 2020 | | SARS-CoV-2 Infection | NA | NA | Socioeconomic determinants = Regression coefficients (p-value)  % Population below poverty = -0.472 (>0.05)  % Minority population = 2.702 (>0.05)  % Uninsured population = 34.067 (>0.05) | The percent of population aged 65+, the percent of the population in poverty, the percent minority, and percent uninsured, population density and percent population tested |
|  |  | | Mortality | NA | NA | Socioeconomic determinants = Regression coefficients (p-value)  % Population below poverty = 4.107 (<0.05)  % Minority population = -1.801 (<0.01)  % Uninsured population = 3.94 (>0.05) | The percent of population aged 65+, the percent of the population in poverty, the percent minority, and percent uninsured, population density and percent population tested |
| Di Girolamo et al. 2020  (Italy) | N=4 427 364  Population based | | Mortality | N= 3301 (0.07%)  Index of deprivation   - Q1 N=666 (20.18%) - Q2 N=618 (18.72%) - Q3 N=563 (17.06%) - Q4 N=657 (19.90%) - Q5 N=784 (23.75%) - Missing N=13 (0.39%)   Household crowding   - Q1 N=704 (21.33%) - Q2 N=630 (19.09%) - Q3 N=673 (20.30%) - Q4 N=603 (18.27%) - Q5 N=683 (20.69%) - Missing N=8 (0.24%)   Proportion of foreign resident population   - Q1 N=618 (18.72%) - Q2 N=591 (17.90%) - Q3 N=664 (20.12%) - Q4 N=687 (20.81%) - Q5 N=737 (22.33%)   Missing N=4 (0.12%) | NA | Socioeconomic determinants = mortality rate ratio MRR (95%CI)  Males  Index of deprivation   - Q1=reference - Q2=1.10 (0.95-1.26) - Q3=1.03 (0.89-1.18) - Q4=1.21 (1.05-1.38) - Q5=1.39(1.21-1.59) - Missing=NA   Household crowding   - Q1=reference - Q2=1.03(0.9-1.19) - Q3=1.11(0.97-1.27) - Q4=1.09(0.95-1.26) - Q5=1.27(1.11-1.46) - Missing=NA   Proportion of foreign resident population   - Q1=reference - Q2=1.04(0.89-1.20) - Q3=1.23(1.07-1.42) - Q4=1.34(1.16-1.54) - Q5=1.57(1.37-1.80) - Missing=NA   Females  Index of deprivation   - Q1=reference - Q2=0.97(0.81-1.16) - Q3=0.96(0.80-1.15) - Q4=1.18(1.00-1.40) - Q5=1.55(1.33-1.82) - Missing=NA   Household crowding   - Q1=reference - Q2=1.02(0.86-1.22) - Q3=1.26(1.07-1.49) - Q4=1.15(0.96-1.36) - Q5=1.54(1.31-1.82) - Missing=NA   Proportion of foreign resident population   - Q1=reference - Q2=1.05(0.88-1.24) - Q3=1.10(0.93-1.30) - Q4=1.08 (0.91-1.28) - Q5=1.20(1.02-1.42)   Missing=NA | Age |
| Daras et al. 2021  (UK) | 6789 Middle Super Output Areas (MSOAs) (average 8242 people)  Residents of 6789 Middle Super Output Areas (MSOAs)  March 1 2020 – May 31 2020 | | Mortality | NA | NA | Socioeconomic determinants = IRR (95% CI)   - Overcrowding (%) = 1.11 (1.06 – 1.15) - Black, Asian and Minority Ethnic groups (%) = 1.08 (1.03 – 1.13) | Duration of epidemic, region and a linear spline for the care home variable above 3 SD |
| Rodriguez-Villamizar et al. 2021  (Columbia) | 772 municipalities  Colombian municipalities  Inclusionup to July 17 2020 | | Mortality | 6 288 COVID-19 deaths | Socioeconomic determinants= MRR (95% CI)   - % Urban population = 0.96 (0.94–0.9) - Population density = 0.99 (0.99–1.00)   Poverty index = 0.99 (0.97–1.02) | Socioeconomic determinants= MRR (95% CI)   - % Urban population = 1.02 (1.01–1.03) - Population density = 1 (0.99–1.00)   Poverty index = 1.03 (1.01–1.05) | Percentage of population 65 years or older, population density, poverty index, hospital beds capacity, percentage of urban population, number of COVID-19 tests at the department level, and prevalence (percentage) of hypertension, diabetes, and chronic renal failure. |
| Khanijahani et al. 2021  (USA) | 73056 tracts /3142 counties  USA counties  Jan 22 2020 - 21 July 2020 | | Mortality | Average of 20.4 per 100,000 population COVID-19 death | NA | Socioeconomic determinants = MRR (95% CI)  % residing in concentrated disadvantaged (10% increments) = 1.14(1.11-1.18)  % residing in Black concentration (10% increments) =1.11 (1.08 – 1.14) | Age, population density, ICU beds, hospital beds, days from the first case, core bases statistical area, region |
| Gorges et al. 2021  (USA) | 13 312 Nursing homes  Residents of nursing homes  January 1 2020 – September 13 2020 | | Mortality | 51 606COVID-19–associated deaths  Mean (SD) of 3.9 deaths per facility | Socioeconomic determinants = Marginal effects (SE)  White Race Q1 (Low) = 3.892 (0.203)  White Race Q2 = 3.518 (0.206)  % White Race Q3= 2.147 (0.170)  White Race Q4 = 1.564 (0-174)  p-value<0.01 | Socioeconomic determinants = Marginal effects (SE)  White Race Q1 (Low) = 0.963(0.226)  White Race Q2 = 1.056 (0.213)  % White Race Q3= 0.572 (0.197)  White Race Q4 = 0.848 (0.213)  p-value<0.01 | Ln certified beds, case-mix index, % resident hypertension, for-profit, govrnement owned, chain, %residents medicaid, % residents medicare, NHC star ratings (1;2;3;4), Adjusted nursing hours/ residency/day, COVID-19 cases/1000 population |
| Li et al. 2020  (USA) | 215 Nursing homes with at least once case  Residents of nursing homes (NHs)  Inclusionas of April 4 2020 | | Mortality | 121.1 deaths (1-406) | NA | Socioeconomic determinants = OR (95% CI)  NHs concentrated by Medicaid residents = 0.91 (0.7-1.18)  NHs concentrated by racial/ethnic minority residents = 0.89 (0.7-1.15) | Nursing home staffing, overall quality of care , concentration of Medicaid residents , and concentration of racial and ethnic minority residents |
| Gross et al. 2020  (USA) | 28 states  General population  Inclusionas of April 21 2020 | | Mortality | NA | Socioeconomic determinants = SMR Standardised mortality ratio (95%CI)  Race   - White = Reference - Black = 3.57 (2.84-4.48)   Ethnicity   - White = Reference - Latinx = 1.88 (1.61-2.19) | NA | NA |
| Fielding-Miller et al. 2020  (USA) | 3 024 counties  General population  Inclusionas of July 12 2020 | Mortality | | NA | NA | Socioeconomic determinants = Regression coefficients (p-value)  % Residents uninsured = 0.17 (0.25)  % Residents in poverty = 4.41 (<0.001) | Percentage farm worker, percentage non-English speaker, percentage uninsured, percentage residents in poverty, percentage residents over 65, residents per square meter, population (thoUSA nds) |

##### Table 3. Characteristics of studies reporting on socioeconomic determinants among population representatives samples – Prognostic role

| **First author (Country)** | **Study design Study setting Study period** | **Study quality score*** | **Follow-up period** | **Covid-19 diagnostics for population selection** | **Outcome measures** | **Study participants included in the analyses and available socio-demographic characteristics** | **List of socioeconomic determinants considered in the study** | **Socioeconomic determinants (definition, measurement, levles and the source of this information; prevalence where available)** | **Association metrics between socioeconomic determinants and the outcome; ; analyses Adjusted (yes/no/both)** | **Total N (%) of people for which the outcome occurred** |
| --- | --- | --- | --- | --- | --- | --- | --- | --- | --- | --- |
| **Individual-level studies** | | | | | | | | | | |
| Baqui et al.  2020  (Brazil) | Cross-sectional  Hospitalised patients  February 27 2020 – May 4 2020 | 8 | NA | RT-PCR | COVID-19 hospital mortality | N = 11 321  Black 6.45%; Asian 1.94%; White 58.7%; Mixed 32.6%; Indigenous 0.2% | Ethnicity | - White N = 4 333 (58.59%) - Black N = 476 ( 6.64%) - East Asian N = 143 (2.04%) - Indigenous N = 15 (0.23%) - Mixed (Pardo) N = 2 404 (32.5%) | HR (95% CI)  Adjusted | N = 3328 deaths (29.4%) |
| Azar et al.  2020  (USA) | Cohort  Patients at large integrated health system  January 1 2020 – April 8 2020 | 7 | 4 months | Positive test result in the electronic health records | Hospital admission, ICU admission, deaths | N = 1052  Mean age 53 (51.8;54.1);  Female 50.8%;  Black 5.8%;  Hispanic 25.8%;  Asian 11.8%;  White 40.7%;  Other 15.8% | Ethnicity  Median income | Ethnicity   - White N = 428 (40.68%) - Asian N = 124 (11.79%) - Black N = 61 (5.8%) - American Indian/Pacific Islander N = 2 (0.2%) - Hispanic N = 272 (25.86%) - Other/Unknown N = 165 (15.68%)   Median income   - Quartile 1 - Quartile 2 - Quartile 3 - Quartile 4 | OR (p-value)  Adjusted | N = 256 hospitalisation (24.33%)  N = 110 ICU admission (10;46%)  N = 51 deaths (4.85%) |
| Argoty-Pantoja et al. 2021  (Mexico) | Cohort  Community - Cases with a positive diagnosis for SARS-CoV2 infection  February 27 2020 – July 30 2020 | 6 | NA | Cases certified by the Institute of epidemiological diagnosis and reference:  - Outpatients  - Hospitalised | Fatality rate | N = 412 017  Female 46.81%;  Indigenous population 1.1% | Ethnicity | - Indigenous N = 4469 (1.08%) - Non-Indigenous N = 407548 (98.92%) | HR (95% CI)  Adjusted | N = 45 754 deaths (11.10%) |
| Bergman et al. 2021  (Sweden) | Case-control  Community - COVID-19 cases  January 30 2020 – September 27 2020 | 6 | NA | No info on the methods of testing  International Classification of Diseases, 10th Revision, Swedish Version, [ICD-10-SE) Code: U071 | COVID-19 diagnosis without hospitalisation, non-ICU hospitalisation with confirmed COVID-19, ICU hospitalization for confirmed COVID-19 | N = 518 739  Control = 434081  COVID-19 diagnosis without hospitalisation = 68 575  Non-ICU hospitalisation with confirmed COVID-19 = 13 589  ICU hospitalization for confirmed COVID-19 = 2494  Female 49.15% | Country of Birth  Education  Disposable family income in 2018 | Country of Birth   - Sweden N = 410092 (79.06%) - Not born in Sweden (20.94%)   Education   - Primary N = 83206 (16.04%) - Secondary N = 178629 (34.44%) - Post-secondary < 3y N = 58601 (11.3%) - Post-secondary> 3y N = 93212 (17.96%) - Missing N = 105 091 (20.26%)   Disposable family income in 2018   - Quintile 1 N = 85870 (16.55%) - Quintile 2 N = 84931 (16.37%) - Quintile 3 N = 83641 (16.12%) - Quintile 4 N = 86044 (16.6%) - Quintile 5 N = 87423 (16.85%) - Missing N = 90 830 (17.51%) | OR (95% CI)  Both | 68 575 infections (13.22%)  13 586 Non-ICO hospitalisations (2.62%)  2494 ICU hospitalisations (0.48%) |
| Elimian et al.  2020  (Nigeria) | Cohort  Community confirmed cases  February 27 2020 – June 8 2020 | 7 | NA | RT-PCR | COVID-19 mortality | N = 3 215 | Occupation  Residential setting | Occupation   - Pupil/ student - Child - Housewife - Trader/business - Health worker - Animal-related work - Farmer - Religious / traditional leader - Transporter - Other   Residential setting   - Rural - Urban | OR (95% CI)  Both | 295 COVID-19 deaths (9.2%) |
| Cifuentes et al. 2021  (Columbia) | Cohort  Nationwide confirmed cases of COVID-19  March 2 2020 – October 26 2020 | 7 | NA | RT-PCR and antigen-based validated tests | COVID-19 death among confirmed cases | N = 1 033 218  Female 49.63%;  White 93.99%;  Gipsy-Roman 36% ; Raizal 13%; Indigenous 2.21%; African Columbian 3.8% | Ethnicity  Area of residence  Type of health insurance regime  Socioeconomic strata | Ethnicity   - White/ Mestizo N = 971 078 (93.99) - African Columbian N = 39304 (3.80%) - Indigenous N = 22787 (2.21%) - Gipsy - Roman N = 36 (0.00%) - Raizal N = 13 (0.00%)   Area of residence   - Urban N = 888915 (86.03%) - Semirural (village) N = 45882 (4.44%) - Sparse rural N = 25993 (2.52%) - Unknown area N = 72428 (7.01%)   Type of health insurance regime   - Contributory regime N = 660085 (63.89%) - Subsidised N = 157540 (15.25%) - Special N = 16769 (1.62%) - Exception N = 48737 (4.72%) - Uninsured N = 18921 (1.83%) - Unknown or pending insurance N = 10384 (1.01%) - Non-registered insurance N = 120782 (11.6%)   Socioeconomic strata   - Very low N = 192501 (18.63%) - Low N = 378559 (36.64%) - Middle low N = 205489 (19.89%) - Middle N = 36257 (3.51%) - Middle high N = 12147 (1.18%) - High N = 6333 (0.61%) - Unkown N = 201932 (19.54%) | OR (95% CI)  Adjusted | 30565 deaths (2.96%) |
| Fabiani et al.  2021  (Italy) | Cohort  Community COVID-19 cases  February 20 2020 – July 19 2020 | 8 | NA | RT-PCR | Case fatality  COVID-19 hospitalisation rates | N = 210 732  Italians 92.51%;  Low-HDI countries 1.55%;  Medium-HDI countries 3.92%;  High-HDI countries 1.9% | Nationality 1  Nationality 2 | Nationality 1   - Italian N = 194 956 (92.51%) - Non- Italian N = 15 504 (7.36%)   Nationality 2   - Italian N = 194 956 (92.51%) - Low HDI countries N = 3255 (1.55%) - Medium HDI countries N = 8246 (3.92%) - High-HDI countries N = 4003 (1.9%) | RR (95% CI)  Both | 80 316 hospitalisations (38;11%)  28 226 deaths (13;39%) |
|  | Cohort  Hospitalised COVID-19 cases  February 20 2020 – July 19 2020 | 8 | NA | RT-PCR | ICU admission  Case fatality | N = 80 316  Italian 93.92%:  Low-HDI countries 1.15%;  Medium-HDI countries 3.7%  High-HDI countries 1.23% | Nationality 1  Nationality 2 | Nationality 1   - Italian N = 75 432 (93.92%) - Non- Italian N = 4884 (6.08%)   Nationality 2   - Italian N = 75 432 (93.92%) - Low HDI countries N = 925 (1.15%) - Medium HDI countries N = 2970 (3.7%) - High-HDI countries N = 989 (1.23%) | RR (95% CI)  Both | 9615 ICU admissions (11;97%)  21 150 deaths (26.33%) |
| Ferrando-Vivas et al. 2021  (UK) | Cohort  Hospitalized COVID-19 cases  March 1 2020-June 22 2020 | 7 | March 1 2020-June 22 2020 | No info on the diagnostic method | Time to death within 30 days of the start of critical care | N=9 990  Median(IQR) age 60(51-68); Female 29.6%; Black 9.8%; Asian 15.2%; White 66.4%; Other 8.6% | Ethnicity  Quintile of deprivation | Ethnicity   - White N=6384 (66.4%) - Asian N=1459 (15.2%) - Black N=940 (9.8%) - Other/mixed N=830 (8.6%)   Quintile of deprivation   - 1 (least deprived) N=1349 (14.6%) - 2 N=1483 (16%) - 3 N=1838 (19.8%) - 4 N=2225 (24%) - 5 (most deprived) N=2372 (25.6%) | HR (95%CI) | 3933 deaths (39.37%) |
| Peres et al.  2021  (Brazil) | Cohort  Hospitalised adult patients with COVID-19  February 16 2020 – August 8 2020 | 6 | NA | RT-PCR | In-hospital mortality | N = 228 196  Median age 61;  IQR 43-45;  Female 43%;  Black 35%;  Asian 11.2%;  White 35.4%;  Indigenous 0.2%;  ≥ High School 16.6% | Race  Level of education | Race   - Black/brown N= 79 914 (35%) - Asian N= 2 558 11.2%; - Indigenous N= 449 (0.2%) - White N= 80 853 (35.4%)   Level of education   - College / University N= 12,728 (5.6%) - High school N = 25,949 (11%) - Up to high shool N = 34,964 (15%) - Illiterate N = 4870 (2.1%) - Unknown N = 149,685 (66%) | OR (95% CI)  Adjusted | 77 967 ICU admission (34%)  85 171 in-hospital COVID-19 deaths (37.32%) |
| Rosenthal et al. 2020  (USA) | Cohort  In-hospital COVID-19 patients  April 1 2020 – May 31 2020 | 7 | NA | Principal or secondary discharge diagnosis of COVID-19 UsingInternational Classification of Diseases, 10th revision, Clinical Modification (ICD-10-CM) diagnosis code U07.1 | In-hospital mortality  Intensive care unit (ICU) admission  Use of invasive mechanical ventilation  Total hospital length of stay (LOS)  ICU LOS  Acute complications  Treatment pattern | N= 35 302  Mean age (± SD) 63.6 (± 17.7);  Median age 65;  IQR 52-77;  Female 46.2%;  Black 23.4;  Hispanic 17.9%;  White 38.9% | Race  Payer type  Ethnicity | Race   - White N = 13 725 (38.9%) - Black N = 8267 (23.4%) - Other/ unknown N = 13 310 (37.7%)   Payer type   - Commercial insurance N = 9022 (25.6%) - Medicaid N = 6371 (18%) - Medicare N = 17 593 (49.8%) - Self-pay N = 898 (2.5%) - Other or unknown N = 1418 (4%)   Ethnicity   - Hispanic N = 6331 (17.9%) - Non-Hispanic N = 20 784 (58.9%) - Other or unknown N = 8187 (23.2%) | OR (95% CI)  Both | 7164 COVID-19 in-hospital deaths (20.3%)  6849 ICU admission (19.4%) |
| Santos et al.  2021  (Brazil) | Cohort  Hospitalised older people due to COVID-19  February 28 2020 – May 18 2020 | 7 | NA | RT-PCR | Time of follow-up until death up to May 18, 2020 | N = 12 199 | Level of education  Race  Zone | Level of education   - Higher education N 3 738 - Intermediate / Elementary education N = 1 413 - Illiterate or low education N = 888   Race   - White or Asian N = 7 009 - Black or mixed N = 6 018   Zone   - Urban N = 17 809 - Rural N = 420 | HR (95% CI)  Both | 7 863 deaths (60.53%) |
| Santos et al. 2020  (Brazil) | Cohort  Patients admitted to Brazilian hospitals due to COVID-19  February 20 2020 – June 2 2020 | 7 | NA | RT-PCR | Survival functions | N = 46 285 | Level of education  Race  Zone | Level of education   - Higher education - Intermediate / Elementary education - Illiterate or low education   Race   - White or Asian - Black or mixed   Zone   - Urban - Rural | HR (95% CI)  Both | 21 408 deaths (46.25%) |
| Telle et al. 2021  (Norway) | Cohort  Community COVID-19 cases in Norway  January 1 2020 – June 6 2020 | 7 | 30 days | RT-PCR | Inpatient hospitalisation  Invasive ventilator treatment  Death within 30 days after the positive test | N = 8 569  Female 50%;  Place of birth:  Norway 71%, Africa Asia Latin-America 21%, Europe USA Oceania 7%, Unknown 2%: | Place of Birth | - Norway - Africa, Asia, Latin-America - Europe, USA , Canada, Oceania - Unknown | RR (95% CI)  Adjusted | 1 172 hospitalisations (14%)  146 mechanical ventilation (1.7%)  227 deaths (2.6%)  Among hospitalised patients, 12% received mechanical ventilation and 2.8% died  Among patients who received invasive mechanical ventilation, 18% died |
| Mendez-Dominquez et al. 2020  (Mexico) | Cross-sectional  Community  February 28, 2020 - April 21, 2020 | 7 | NA | RT-PCR (suspectedcases) | COVID-19 lethality (fatal cases) | N = 17 763  Mean age (± SD) 46.43 (0.55);  Female 55.31% | Migration  Indigenous ethnicity  Population affiliated to public health institutions  Urbanization  Interstate migrant | Migration. Refers to national and international migratory movements  Urbanization level: Percentage of the population living in communities with urban services and infrastructure. | IRR (95% CI)  OR (95% CI)  Adjusted | 968 COVID-19 deaths (5.55%) |
| Wollenstein- et al. 2020  (Brazil) | Cohort  Hospitalised COVID-19 patients in Brazil  March 1 2020 – June 30 2020 | 8 | NA | Positive tests from ARDS surveillance database repository | Mechanical ventilation  Mortality | N = 113 314  Female 43.4%;  Black 36.2%;  White 28.9%;  Yellow 1%;  Indigenous 0.3%;  ≥ High School 17.1% | Race  Schooling | Race   - Indigenous N = 366 (0.3%) - Yellow N = 1 115 (1%) - White N = 32 704 (28.9%) - Brown/Black N = 40 993 (36.2%)   Schooling   - Schooling No Education N = 2 799 - Schooling elementary 1-5 N = 9 374 (8.3%) - Schooling elementary 6-9 N = 6 727 (5.9%) - Schooling Medium 1-3 N = 12 629 (11.2%) - Schooling Superior N = 6 572 (5.8%) | OR (95% CI)  Adjusted | 50 387 deaths (44.47%) |
| Wu et al. 2020  (China) | Cohort  Community COVID-19 patients in Hubei province  December 10, 2019 - February 27 2020 | 7 | Until March 18 2020 | Four diagnosis:  - RT-PCR  - Clinically (symptoms+ pulmonary imaging)  - Clinical symptoms and exposure history | COVID-19 case fatality | N = 21 392  Female 48.12% | Residence  Occupation | Residence   - Local N = 17 828 (83.33%) - Migrant N = 3 564 (16;67%)   Occupation   - Medical-related N = 1 113 (5.20%) - Service-related N = 238 (1.11%) - Office worker N = 4 833 (22.59%) - Home worker N = 13 507 (63.14%) - Others N = 1 701 (7.95%) | HR (95% CI)  Both | 1 020 COVID-19 deaths (4.77%) |
| Mehta et al. 2021  (USA) | Cohort  Nursing home residents with COVID-19 infection  April 1, 2020 - September 30, 2020 | 7 | Until October 31,2020 | (ICD-10-CM) diagnosis code specific for COVID19 (U07.1) | Mortality within 30 days after diagnosis  Hospitalisation within 30 days after diagnosis | N = 137 119  Black 17.3%;  Hispanic 66%;  Asian 17.3%;  White 7.6%;  Other 2.8% | Ethnicity | - Black N = 23 698 - Asian N = 3 899 - Hispanic or Latino N = 10 346 - Other N = 644 - White = 98 532 | HR (95% CI)  Adjusted | 29204 COVID-19 hospitalisations (21.30%) |
| Hamadah et al. 2020  (Kuwait) | Cohort  Hospitalised COVID-19 cases  24 Feb 2020 - 20 April 2020 | 7 | 2  months | RT-PCR | ICU admission  Mortality | N = 1123  Mean age (SD) 41.9 (14.6);  Minimum – Maximum age (1-93);  Female 18.7% | Nationality | - Kuwaiti 26.17% - Non-Kuwaiti 73.83% | OR (95% CI)  Adjusted | 51 COVID-19 ICU admission (4.5%)  40 COVID-19 deaths (3.6%) |
| Kim et al. 2020  (South Korea) | Cohort  Confirmed COVID-19 cases (community)  Study end date till March 26, 2020 | 6 | NA | NA  Information from registry | Confirmed cases  Deaths  Crude death rate | N = 9148  Female 61% | District  Health insurance premium level  Self-employed  Employee | District   - Large urban - Small urban - Rural urban   Health insurance premium level   - Medicaid N = 727   Self-employed   - Lowest N = 661 - Low N =454 - Middle N = 469 - High N = 424 - Highest N=392   Employee   - Lowest N = 1518 - Low N = 1 442 - Middle N = 1 189 - High N = 934 - Employee with highest income level N = 938 | OR (95%CI)  Adjusted | 3 556 COVID-19 cases (38.87%)  67 deaths (73.24%) |
| Patel et al.  2020  (UK) | Cohort  Community cases  March 16,2020 - April 14,2020 | 7 | NA | Nose swab, throat swabs, lower respiratory sputum samples | Hospitalisation | N = 418 794  Age range (40-69):  Female 40%;  Black 69%;  Hispanic 55%;  Asian 18% | Race  Townsend Index  Average income | Race   - White N = 400 438 - Asian N = 10 614 - Black N = 7 714 | OR (95% CI)  Both | COVID-19 hospitalisations |
| Fisman et al.  2020  (Canada) | Cohort  Community COVID-19 cases  January 23 2020 – May 15 2020 | 7 | 4 months | RT-PCR and nucleid acid sequencing | COVID-19 mortality | N = 21 922  Median age 55;  Female 57%; | Low income  Non-health care worker  Non-homeless shelter  worker  Non-homeless | NA | OR (95%CI)  UnAdjusted | 13% COVID-19 Hospitalisation  2% Intubation or Mechanical ventilation  Case fatality 8% |
| Guo et al. 2020    (China) | Cross-sectional  Older persons with COVID-19 in Wuhan  December 12 2019 – March 17 2020 | 6 | 3 months | Epidemiological history and clinical manifestations,  RT-PCR  Viral gene identified by gene sequencing | COVID-19 death | N = 14 238  Female 49.59% | Location | Location   - Downtown N = 11 610 (81.54%) - Suburb N = 2 628 (18.46%) | Fatality ratio  UnAdjusted | Attributable fatality ratio = 222.57/ 100 000  Crude fatality ratio = 19.37% |
| Ibarra-Nava et al. 2021  (Mexico) | Cross-sectional  Adult patients with COVID-19  February 28 2020 – August 3 2020 | 7 | NA | laboratory confirmed | COVID-19 death  Hospitalisation  ICU admission | N = 416 546  Mean age (SD) 49.6(14.6);  Female 46.8%;  Non-Indigenous 98.9%  Indigenous 1.1% | Ethnic group | Ethnic group   - Non-Indigenous N = 412 368 (98.9%) - Indigenous N = 4 178 (1.1%) | OR (95% CI)  Adjusted | 113 853 COVID-19 hospitalisations (27;33%)  8 870 ICU admission (2.13%)  46 508 deaths (11.25%) |
|  | Cross-sectional  Hospitalised patients with COVID-19  February 28 2020 – August 3 2020 | 7 | NA |  | COVID-19 death | N = 113 853  Female 38.59%;  Non-Indigenous 98.6%  Indigenous 1.4% |  | Ethnic group   - Non-Indigenous N = 112 251 (98.6%) - Indigenous = 1 602 (1.4%) | OR (95% CI)  Adjusted | NA |
| Mak et al. 2021  (UK) | Cohort  COVID-19 in-patients  March 1 2020 - November 30 2020 | 6 | NA | RT-PCR  International Classification of Diseases 10th edition (ICD-10) | COVID-19 mortality | N = 2 812  Mean age (SD) 69.2 (8.7);  Female 45.1;  Black 3.3%;  Asian 4.2%;  White 90;6%;  Other 1.9% | Ethnicity  Income  Education  Townsend deprivation index | Ethnicity   - White N = 2 543 (90.6%) - Asian N = 117 (4.2%) - Black N = 93 (3;3%) - Others N = 54 (1.9%)   Education   - Low N = 834 (30.1%) - Intermediate N = 1 348 (48.6%) - High N = 592 (21.3%)   Income   - < £18,000 N = 852 (36.9%) - 18,000–30,999 N = 592 (25.6%) - £31,000–51,999 N = 497 (21.5%) - ≥£52,000 N = 371 (16.1%)   Townsend deprivation quintile:   - 1 (least deprived) N = 391 (13.9) - 2 N = 458 (16.3%) - 3 N = 506 (18%) - 4 N = 591 (21%) - 5 (most deprived) N= 866 (30.8%) | OR (95%CI)  Adjusted | 417 COVID-19 deaths (14.83%) |
| Navaratnam et al. 2021  (UK) | Cohort  Hospitalized COVID-19 patients  March 1 2020 – May 31 2020 | 7 | NA | International Classification of Diseases 10th edition (ICD-10) codes U071 and U072. | In-hospital death | N = 79 124  Female 51.43%; Black 6.3%, Asian 9%, White 81.67%; Other 4.2%; Mixed 0.94% | Ethnicity | Deprivation score  Ethnicity   - White N = 64 615 - Asian or Asian British N = 7117 - Black or Black British N = 4 983 - Mixed N = 746 - Other ethnic groups N = 3 291 | OR (95% CI)  Adjusted | 28 200 in-hospital deaths (30.8%) |
| Ortiz-Prado et al. 2021  (Ecuador) | Cross-sectional  Country-wide COVID-19 cases  Study started in 2020 until April 18 2020 | 8 | NA | RT-PCR | COVID-19 death | N = 9 468  Female 44.6%;  Mestizos 78%; Indigenous 0.79; CaucAsians 0.84%; Afro-Ecuadorians/Black 0.1%;  Single 39.3%;  Married/Cohabiting 51%;  Divorced / Separated 5.4%;  Widowed 1.3%;  Other 3% | Occupation | Occupation   - Politicians N = 18 - Registered Nurses N = 319 - Health Care professionals no physicians N = 587 - Medical doctors N = 890 - Dentists N = 33 - Civil Services N = 340 - Others N = 431 - Blue Collar N = 1 576 - Unemployed N = 367 - Retired N = 214 - Prisoners N = 4 - White collar jobs N = 18 | RR (95% CI)  Adjusted | 474 COVID-19 deaths (5%) |
| Poulson et al. 2020  (USA) | Cross-sectional  COVID-19 cases  April 5 2020 until May 18 2020 | 7 | NA | Laboratory confirmed cases | COVID-19 hospitalisation  COVID-19 ICU admission  COVID-19 mechanical ventilation  COVID-19 mortality | N = 124 780  Female 51.2%;  Back 61.3%;  Whit 38.7% | Ethnicity | Ethnicity   - Non-Hispanic White - Black | RR (95% CI)  Both | 38 199 Hospitalisations (16.19%)  5 996 ICU admissions (4.8%)  4 330 Mechanical ventilations (3.5%)  15 423 deaths (12.36%) |
| Zelner 2021  (USA) | Cohort  COVID-19 cases in the general population  March 8 2020 until July 5 2020 | 7 | NA | RT-PCR | COVID-19 mortality | N =49 701 | Race | Race   - Black N = 19 662 - Latino N = 3 657 - Other N = 1 612 - Asian/Pacific Islander N = 1 346 - White N = 23 301 - Native American N = 123 | MRR (95%IC)  Adjusted | 5 815 deaths (11.7%) |
| **Ecological studies** | | | | | | | | | | |
| Loomba et al. 2020  (USA) | Cohort  COVID-19 positive admissions (CPAs)  March 14, 2020 - April 14, 2020 | 7 | NA | Publicly available COVID-19 dashboard provided by the Virtual Pediatric System | COVID-19 ICU admission | N = 205 CPAs | Percent households below poverty line  Percent households with below high school as the highest adult education level  Percent paediatric who have health insurance  Percent paediatric who have a medical home  Percent Hispanic  Percent White  Percent Black  Percent other race  Population density | NA | Beta coefficient (p-value) | 2.8 COVID-19 positive admissions (CPAs) per million children |
| Millan-Guerrero et al. 2021  (Mexico) | Cohort  All indiviudals diagnosed with COVID-19  February 27, 2020 - July 1, 2020 | 7 | NA | RT-PCR | COVID-19 Survival | N = 231 772  Female 45.4%  Indigenous (1.1%), non-Indigenous (98.9%) | Poverty level | - No poverty - Extreme poverty level | HR (95% CI)  Both | NA |
| Ojinnaka et al. 2021  (USA) | Cohort  Community - County-level cases  March 4, 2020 - August 1, 2020 | 6 | NA | NA  Data obtained from Texas Department of State Health Services | COVID-19 Mortality | 254 Texas counties  Asian 6.26%;  White 35.35%;  Other 1.33%;  High school graduation rate 93.65% |  | - Percent Black 55.39% - Percent Non-Hispanic White 6.26% - Percen Asian 1.33% - Percent Hispanic 35.35% - Percent uninsured 21.21% - Primary case physican rate 40.5% - High school graduation rate 93.65% - Percent unemployed 3.87% - Black White segregation index 39.98%  1. Percent rural 55.52% | OR (95% CI)  Both | NA |
| Lewis et al. 2020  (USA) | Cross-sectional  General population COVID-19 cases  July 3 2020 – July 9 2020 | 7 | NA | RT-PCR | COVID-19 hospitalisation | 99 Utah small statistical areas | Level of deprivation | - Very low (least deprived) - Low - Average - High  1. Very high (most deprived) | OR (95%CI)  Both | 1 781 hospitalisations |
| Akanbi et al. 2020  (USA) | Cross-sectional  General population  April 3 2020 – May 16 2020 | 7 | NA |  | COVID-149 mortality | 70 Zip codes  Mean age 41.41 ; Age range 37.92-44.1 ; Black 13.6%; Asian 5.6%; White 77.3%; Other 1%; Native American 0.3%; Two or more races 2.2% | Ethnicity  Education level  Median family income  Poverty level | Ethnicity: % of Black people per ZIP code   2. | IRR (95%CI)  Both | Mean (SD)  12.77 deaths (15.21) |

##### Table 4. Association for socioeconomic determinants and outcomes among population representative samples – Prognostic role

| **First author (Country)** | **Sample** | **Outcome** | **N (%) of people for which the outcome occurred (total and per socioeconomic determinants level where available)** | **Association with socioeconomic determinants - Unadjusted** | **Association with socioeconomic determinants - Adjusted** | **Adjustment factors** |
| --- | --- | --- | --- | --- | --- | --- |
| **Individual-level studies** | | | | | | |
| Azar et al.  2020  (USA) | N = 1 052  Patients at large integrated health system  January 1 2020 – April 8 2020 | Hospitalisation COVID-19 | 256 hospitalisation (24.33%)  Ethnicity   - White N = 110 (43%) - Asian N = 22 (8.6%) - Black N = 32 (12.5%) - American Indian/Pacific Islander N = 0 - Hispanic N = 71 (27.74%) - Other/Unknown N = 21 (8.2%) | NA | Socioeconomic determinants = OR (p-value)  Ethnicity   - White = Reference - African American = 2.7 (p-value=0.007)   Median income   - Quartile 1 = Reference - Quartile 3 = 0.24 (p-value<0.05) - Quartile 4 = 0.55 (p-value<0.05) | Sex, comorbidities, age, insurance type, income, ethnicity, homeless status, smoking status |
| Patel et al.  2020  (UK) | N = 418 794  Community cases  March 16,2020 - April 14,2020 | Hospitalisation COVID-19 | NA | Socioeconomic determinants = OR (95% CI)  Race   - White = Reference - Asian = 2.16(1.47-3.16) - Black = 3.41(2.38-4.88)   Townsend deprivation index = 1.13 (1.1-1.16)  Average income=0.83(0.77-0.9) | Socioeconomic determinants = OR (95% CI)  Race   - White = Reference - Asian = 1.75(1.08-2.85) - Black = 2.38(1.52-3.74)   Townsend index = 1.09 (1.05-1.12)  Average income = 1.01(0.92-1.11) | Age, sex, geographical region, townsend deprivation index, income, comorbidities |
| Bergman, J. et al. 2021  (Sweden) | N = 518 739  Community  January 1 2020 – September 27 2020 | ICU admission | 2494 ICU hospitalisations (0.48%)  Country of Birth   - Sweden N = 1376 (55.2 %)   Education   - Primary N = 625 (26.4%) - Secondary N = 1075(45.4%) - Post-secondary < 3y N = 292 (12.3%) - Post-secondary> 3y N = 377 (15.9%)   Disposable family income in 2018   - Quintile 1 N = 575 (23.5%) - Quintile 2 N = 522(21.3%) - Quintile 3 N = 487(19.9%) - Quintile 4 N = 432 (17.6%) - Quintile 5 N = 435(17.7%) | Socioeconomic determinants = OR (95% CI)  Country of Birth   - Sweden 0.30 (0.28 - 0.32) - Not born in Sweden = Reference   Education   - Primary = Reference - Secondary = 0.79 (0.72-0.88) - Post-secondary < 3y = 0.65 (0.57-0.75) - Post-secondary> 3y = 0.54 (0.47-0.61)   Disposable family income in 2018   - Quintile 1 = Reference - Quintile 2 = 0.90 (0.80-1.02) - Quintile 3 = 0.84 (0.75-0.95) - Quintile 4 = 0.75 (0.66-0.85) - Quintile 5 = 0.75 (0.66-0.85) | Socioeconomic determinants = OR (95% CI)  Country of Birth   - Sweden = 0.33 (0.3-0.37) - Not born in Sweden = Reference   Education   - Primary = Reference - Secondary = 0.91 (0.82 -1.01) - Post-secondary < 3y = 0.8 (0.69-0.92) - Post-secondary> 3y = 0.73 (0.63 – 0.83)   Disposable family income in 2018   - Quintile 1 = Reference - Quintile 2 = 0.93 (0.81- 1.07) - Quintile 3 = 0.96 (0.83 -1.1) - Quintile 4 = 0.94 (0.81 -1.08) - Quintile 5 = 0.84 (0.72-0.97) | Sex, age, education, family disposable income, Stockholm residence, long-term care facility, born in Sweden |
| Baqui et al.  2020  (Brazil) | N = 11321  Hospitalised patients  February 27 2020 – May 4 2020 | Survival | N = 3328 (29.40%)   - White N = 1696 (51%) - Black N = 245 (7.36%) - East Asian N = 69 (2.1%) - Indigenous N= 8 (0.24%) - Mixed (Pardo) N = 1310 (39.36%) | NA | Socioeconomic determinants = HR (CI 95%)   - White = Reference - Black = 1.32(1.15-1.52) - East Asian = 1.12(0.88-1.44) - Mixed (Pardo) = 1.45(1.33-1.58) | Age, comorbidities, sex |
| Elimian et al. 2020  (Nigeria) | N = 3 215  Community confirmed cases  February 27 2020 – June 8 2020 | Mortality | 295 COVID-19 deaths (9.2%) | Socioeconomic determinants = IRR (CI 95%)  Occupation   - Pupil/ student = Reference - Child = Omitted - Housewife = 8.31 (2.08 to 33.17) - Trader/bUSA iness = 7.16 (2.11 to 24.28) - Health worker = 2.46 (0.69 to 8.82) - Animal-related work = Omitted - Farmer = 24.93 (6.83 to 90.94) - Religious / traditional leader = 22.56 (4.53 to 112.41) - Transporter = 11.28 (1.72 to 74.02) - Other = 7.63 (2.41 to 24.21) | Socioeconomic determinants = IRR (CI 95%)  Occupation   - Pupil/ student Reference - Child = Omitted - Housewife = 2.68 (0.54 to 13.23) - Trader/bUSA iness = 2.13 (0.54 to 8.50) - Health worker= 1.57 (0.38 to 6.54) - Animal-related work= Omitted - Farmer = 7.56 (1.70 to 33.53) - Religious / traditional leader = 1.24 (0.17 to 8.95) - Transporter = 5.46 (0.58 to 51.36) - Other = 3.74 (1.03 to 13.56) | Age, sex, geopolitical zone, travel history, clinical signs and symptoms |
| Fabiani et al. 2021  (Italy) | N = 210 732  Community COVID-19 cases  February 20 2020 – July 19 2020 | Hospitalisation for COVID-19 | Socioeconomic determinants = RR (95% CI)  80 316 hospitalisations (38;11%)  Nationality 1   - Italian N = 75 432 (94%) - Non- Italian N = 4884 (6%)   Nationality 2   - Italian N = 75 432 (94%) - Low HDI countries N = 925 (0.4%) - Medium HDI countries N = 2970 (1.41%) - High-HDI countries N = 989 (0.47%) | Socioeconomic determinants = RR (95% CI)  Nationality 1   - Italian = Reference - Non- Italian = 0.82 (0.79–0.84)   Nationality 2   - Italian = Reference - Low HDI countries = 0.74 (0.69–0.78) - Medium HDI countries = 0.93 (0.90–0.97 - High-HDI countries = 0.64 (0.60–0.68) | Socioeconomic determinants = RR (95% CI)  Nationality 1   - Italian = Reference - Non- Italian = 1.39 (1.33–1.44)   Nationality 2   - Italian = Reference - Low HDI countries = 1.59 (1.48–1.71) - Medium HDI countries = 1.46 (1.39–1.53) - High-HDI countries = 1.15 (1.07–1.23) | Sex, age group, geographical macro-area of diagnosis, level of urbanization of the place of residence, pre-existing comorbidities, calendar period of diagnosis and accounting for the random effect due regional contextual differences. |
|  | N = 80 316  Hospitalised COVID-19 cases  February 20 2020 – July 19 2020 | ICU admission | 9 615 ICU admissions (11.97%)  Nationality 1   - Italian N = 9 066 (94.2%) - Non- Italian N = 549 (5.71%)   Nationality 2   - Italian N = 9 066 (94.3%) - Low HDI countries N = 92 (0.96%) - Medium HDI countries N = 354 (3.69%) - High-HDI countries N = 103 (10.4%) | Nationality 1   - Italian = Reference - Non- Italian N = 0.94 (0.86–1.02)   Nationality 2   - Italian = Reference - Low HDI countries = 0.83 (0.67–1.02) - Medium HDI countries = 0.99 (0.89–1.10) - High-HDI countries = 0.87 (0.71–1.05) | Socioeconomic determinants = RR (95% CI)  Nationality 1   - Italian = Reference - Non- Italian N = 1.19 (1.07–1.32)   Nationality 2   - Italian = Reference - Low HDI countries = 1.16 (0.93–1.44) - Medium HDI countries = 1.25 (1.11–1.42) - High-HDI countries = 1.15 (0.88–1.32) | Sex, age group, geographical macro-area of diagnosis, level of urbanization of the place of residence, pre-existing comorbidities, calendar period of diagnosis and accounting for the random effect due regional contextual differences. |
|  | N = 80 316  Hospitalised COVID-19 cases  February 20 2020 – July 19 2020 | Mortality | 21 150 deaths (26.33%)  Nationality 1   - Italian N = 20 823 (98.45%) - Non- Italian N = 327 (1.55%)   Nationality 2   - Italian N = 20 823 (98.45%) - Low HDI countries N = 37 (0.17%) - Medium HDI countries N = 206 (0.97%)   High-HDI countries N = 84 (0.4%) | Nationality 1   - Italian = Reference - Non- Italian N = 0.24 (0.22–0.27)   Nationality 2   - Italian = Reference - Low HDI countries = 0.14 (0.10–0.20) - Medium HDI countries = 0.25 (0.22–0.29) - High-HDI countries = 0.31 (0.25–0.38) | Socioeconomic determinants = RR (95% CI)  Nationality 1   - Italian = Reference - Non- Italian N = 0.93 (0.83–1.04)   Nationality 2   - Italian = Reference - Low HDI countries = 0.93 (0.67–1.29) - Medium HDI countries = 0;93 (0.80–1.07) - High-HDI countries = 0.94 (0.76–1.17) | Sex, age group, geographical macro-area of diagnosis, level of urbanization of the place of residence, pre-existing comorbidities, calendar period of diagnosis and accounting for the random effect due regional contextual differences. |
|  | N = 210 732  Community COVID-19 cases  February 20 2020 – July 19 2020 | Mortality | 28 226 deaths (13;39%)  Nationality 1   - Italian N = 27 836 (98.62%) - Non- Italian N = 390 (1.38%)   Nationality 2   - Italian N = 27 836 (98.62%) - Low HDI countries N = 53 (0.19%) - Medium HDI countries N = 237 (0.84%) - High-HDI countries N = 100 (0.35%) | Socioeconomic determinants = RR (CI 95%)  Nationality 1   - Italian = Reference - Non- Italian 0.82 (0.79–0.84)   Nationality 2   - Italian = Reference - Low HDI countries = 0.74 (0.69–0.78) - Medium HDI countries = 0.93 (0.90–0.97 - High-HDI countries = 0.64 (0.60–0.68) | Socioeconomic determinants = RR (CI 95%)  Nationality 1   - Italian = Reference - Non- Italian = 1.39 (1.33–1.44)   Nationality 2   - Italian = Reference - Low HDI countries = 1.59 (1.48–1.71) - Medium HDI countries = 1.46 (1.39–1.53) - High-HDI countries = 1.15 (1.07–1.23) | Sex, age group, geographical macro-area of diagnosis, level of urbanization of the place of residence, pre-existing comorbidities, calendar period of diagnosis and accounting for the random effect due regional contextual differences. |
| Ferrando-Vivas et al. 2021  (UK) | N=9 990  Hospitalized COVID-19 cases  March 1 2020-June 22 2020 | Survival | N=3 933  Ethnicity   - White N=2530 (67.1%) - Asian N=591 (15.7%) - Black N=373 (9.9%) - Other/mixed N=278 (7.4%)   Quintile of deprivation   - 1 (least deprived) N=519 (14.5%) - 2 N=583 (16.3%) - 3 N=702 (19.6%) - 4 N=869 (24.2%) - 5 (most deprived) N=913 (25.5%) | NA | Socioeconomic determinants=HR (95%CI)  Ethnicity   - White=reference - Asian =1.270(1.154-1.397) - Black =1.053(0.933-1.190) - Other/mixed =0.991(0.872-1.127)   Quintile of deprivation   - 1 (least deprived)reference - 2=1.017(0.901-1.149) - 3=1.006(0.897-1.128) - 4=1.063(0.951-1.188) - 5 (most deprived) =1.137(1.011-1.279) | Age, BMI, sex, ethnicity, quintile of deprivation, patients' characteristics |
| Peres et al.  2021  (Brazil) | N = 228 196  Hospitalised adult patients with COVID-19  February 16 2020 – August 8 2020 | Mortality | 85 171 in-hospital COVID-19 deaths (37.32%)  Race   - Black/brown N= 33730 (39.60%) - Asian N= 99 (1.17%); - Indigenous N= 191 (0.22%) - White N= 29 033 (34.09%)   Level of education   - College / University N= 2 692 (3.16%) - High school N= 7 218 (8.47%) - Up to high school N= 15 429 (18.12%) - Illiterate N= 2 925 (3.43%) | NA | Socioeconomic determinants = OR (CI 95%)  Race   - Black/brown = 1.16 (1.13 - 1.18) - Asian = 0.96 (0.88 - 1.05) - Indigenous = 1.16 (0.94 - 1.42) - White = Reference   Level of education   - College / University = Reference - High school = 1.42 (1.34 - 1.50) - Up to high school = 1.73 (1.65 - 1.82) - Illiterate = 2.04 (1.89 - 2.21) | Age, sex, education, residence, comorbidities, ethnicity |
| Rosenthal et al. 2020  (USA) | N= 35 302  In-hospital COVID-19 patients  April 1 2020 – May 31 2020 | Mortality | 7164 COVID-19 in-hospital deaths (20.3%) | Socioeconomic determinants = OR (CI 95%)  Race   - White = Reference - Black = 0.78 (0.73-0.84) - Other/ unknown = 0.82 (0.77-0.87)   Payer type   - Commercial insurance = Reference - Medicaid =1.21 (1.10-1.34) - Medicare = 3.67 (3.40-3.96) - Self-pay =1.02 (0.85-1.23) - Other or unknown =0.89 (0.71-1.13) | Socioeconomic determinants = OR (CI 95%)  Race   - White = Reference - Black = 0.75 (0.69-0.82) - Other/ unknown = 0.95 (0.88-1.03)   Payer type   - Commercial insurance = Reference - Medicaid = 1.29 (1.14-1.45) - Medicare = 1.33 (1.20-1.48) - Self-pay = 1.33 (1.00-1.75) - Other or unknown = 1.12 (0.91-1.39) | Demographic characteristics: age, sex, race, payer type  Visit characteristics: admission point of origin)  Hospital characteristics: geographic region, teaching status  Clinical characteristics: comorbidities, complications  Medications: ACE inhibitors, statins, hydroxychloroquine and/or azithromycin Use, β blockers, calcium channel blockers), and supplements Usedduring hospitalization |
| Telle et al. 2021  (Norway) | N = 8 569  Community COVID-19 cases in Norway  January 1 2020 – June 6 2020 | Hospitalisation for COVID-19 | 1 172 hospitalisations (14%)  Place of Birth   - Norway 13% - Africa, Asia, Latin-America 19% - Europe, USA , Canada, Oceania 10%   Unknown 16% | NA | Socioeconomic determinants = RR (95% CI)  Place of Birth   - Norway = Reference - Africa, Asia, Latin-America = 2.1 (1.9-2.4) - Europe, USA , Canada, Oceania = 1 (0.8-1.3) - Unknown = 0.8 (0.6-1.2) | Age, sex, comorbidity proxy, nursing home resident |
|  |  | Mechanical ventilation | 146 mechanical ventilation (1.7%)  Place of Birth   - Norway 1% - Africa, Asia, Latin-America 3% - Europe, USA , Canada, Oceania 1%   Unknown 1% | NA | Socioeconomic determinants = RR (95% CI)  Place of Birth   - Norway = Reference - Africa, Asia, Latin-America = 2.7 (1.9–3.8) - Europe, USA , Canada, Oceania = 1.2 (0.6–2.4) - Unknown = 0.6 (0.2–2.3) | Age, sex, comorbidity proxy, nursing home resident |
|  |  | Mortality | 227 deaths (2.6%)  Place of Birth   - Norway 3% - Africa, Asia, Latin-America 1% - Europe, USA , Canada, Oceania 1%   Unknown 7% | NA | Socioeconomic determinants = RR (CI 95%)  Place of Birth   - Norway = Reference - Africa, Asia, Latin-America = 2.3 (1.5–3.5) - Europe, USA , Canada, Oceania = 0.9 (0.4–1.9) - Unknown = 0.7 (0.4–1.2) | Age, sex, comorbidity proxy, nursing home resident |
| Wollenstein- et al. 2020  (Brazil) | N = 113 314  Hospitalised COVID-19 patients in Brazil  March 1 2020 – June 30 2020 | Mechanical ventilation | NA | NA | Socioeconomic determinants = OR (95% CI)  Race   - White = 0.948 (0.906-0.991)   Schooling   - Schooling Medium = 0.768 (0.721-0.819) - Schooling Superior = 0.708 (0.645-0.778) | Age, region, gender, comorbidities, clinical symptoms, race, education, hospital (public / private) |
|  |  | Mortality | 50 387 deaths (44.47%) | NA | Socioeconomic determinants = OR (CI 95%)  Race   - Race Indigenous = 1.181 (0.871-1.603) - Race Yellow = 1.156 (0.978-1.366) - Race White = 0.94 (0.902-0.98)   Schooling   - Schooling No Education = 1.066 (0.952 -1.194) - Schooling elementary 1-5 = 0.984 (0.925-1.045 - Schooling Medium 1-3 = 0.858 (0.808-0.91) - Schooling Superior = 0.629 (0.577-0.686) | Age, region, gender, comorbidities, clinical symptoms, race, education, hospital (public / private) |
| Argoty-Pantoja et al. 2021  (Mexico) | N = 299 349  Community - Cases with a positive diagnosis for SARS-CoV2 infection:  - Outpatients  February 27 2020 – July 30 2020 | Survival | N = 5 110 deaths (1.71%)   - Indigenous N = 102 (2%) - Non-Indigenous N = 5008 (98%) | NA | Socioeconomic determinants = HR (CI 95%)   - Non-Indigenous = Reference - Indigenous = 1.63 (1.34 -1.98) | Sex, age, COPD, metabolic comorbidities, chronic kidney disease |
|  | N = 112 668  Community - Cases with a positive diagnosis for SARS-CoV2 infection:  - Hospitalised  February 27 2020 – July 30 2020 | Survival | N = 40 644 deaths (36.07%)   - Indigenous N = 666 (1.64%) - Non-Indigenous N = 39 978 (98.36%) | NA | Socioeconomic determinants = HR (CI 95%)   - Non-Indigenous = Reference - Indigenous = 1.01 (0.94 -1.09) | Sex, age, COPD, metabolic comorbidities, chronic kidney disease |
| Mendez-Dominquez et al. 2020  (Mexico) | N = 17 763  Community  February 28, 2020 - April 21, 2020 | Mortality | 968 COVID-19 deaths (5.55%) | NA | Socioeconomic determinants = IRR (CI 95%)   - Migration = 1.02(0.98-1.02) - Indigenous ethnicity = 0.82(0.88-1.01) - Population affiliated to public health institutions = 0.79(0.78-0.8) - Urbanisation = 1.58(1.55-1.62)   Socioeconomic determinants = OR (CI 95%)  Interstate migrant = 2.01(1.46-2.76) | Population age composition, gender  Age, gender, and place of residence of patients for the variable interstate migration |
| Wu et al. 2020  (China) | N = 21 392  Community COVID-19 patients in Hubei province  December 10, 2019 - February 27 2020 | Survival | 1 020 COVID-19 deaths (4.77%)  Residence   - Local N = 749 (73.43%) - Migrant N = 271 (26.57%)   Occupation   - Medical-related N = 12 (1;18%) - Service-related N = 7 (0;69%) - Office worker N = 100 (9.8%) - Home worker N = 810 (79.41%) - Others N = 91 (8;92%) | Socioeconomic determinants = HR (CI 95%)  Residence   - Local = Reference - Migrant = 1.75 (1.52, 2.01)   Occupation   - Medical-related = Reference - Service-related = 2.86 (1.12, 7.25) - Office worker = 1.94 (1.06, 3.52) - Home worker = 5.73 (3.24, 10.13) - Others = 5.07 (2.78, 9.26) | Socioeconomic determinants = HR (CI 95%)  Residence   - Local = Reference - Migrant = 0.86 (0.74, 1.01)   Occupation   - Medical-related = Reference - Service-related = 3.07 (1.20, 7.85) - Office worker = 1.92 (1.05, 3.50) - Home worker = 1.76 (0.98, 3.16) - Others = 2.13 (1.16, 3.92) | Sex, age, time between symptom onset and diagnosis occupation, residence, period, hospital level, area, severity |
| Hamadah et al. 2020  (Kuwait) | N = 1123  Hospitalised COVID-19 cases  24 Feb 2020 - 20 April 2020 | ICU admission | 51 COVID-19 ICU admission (4.5%)  Kuwaiti N = 11  Non-Kuwaiti N = 40 | NA | Kuwaiti = 1.93 (0.77-5.483)  Non-Kuwaiti | Age, smoking, comorbidities (HTA, diabetes, CVD, asthma, cancer), gender, BMI |
|  |  | Mortality | 40 COVID-19 deaths (3.6%)  Kuwaiti N = 10 (25%)  Non-Kuwaiti N = 30 (75%) | NA | Kuwaiti = Reference  Non-Kuwaiti = 1.627 (0.535-5.915) | age, smoking, comorbidities (HTA, diabetes, CVD, asthma, cancer), gender, BMI |
| Kim et al. 2020  (South Korea) | N = 9148  Confirmed COVID-19 cases (community)  Study end date till March 26, 2020 | Mortality | 67 deaths (0.73%)   - Medicaid N = 26 (38.81%) - Lowest N = 8 (11.94%) - Low N = 4 (5.97%) - Middle N =7 (10.45%) - High N = 6 (8.96%) - Highest N = 12 (17.91%) - Lowest N = 18 (26.87%) - Low N = 12 (17.91%) - Middle N = 13 (19.40%) - High N = 12 (17.91%) - Employee with highest income level N = 12 (17.91%) | NA | Socioeconomic determinants = OR (CI 95%)  District   - Large urban = Reference - Small urban Rural = 1.73 (0.91–3.30) - Rural = 1.90 (1.15–3.16)   Health insurance premium level   - Medicaid = 2.81 (1.35–5.83)   Self-employed   - Lowest = 1.19 (0.46–3.05) - Low = 1.25 (0.38–4.15) - Middle High = 1.63 (0.61–4.39) - Highest = 1.66 (0.59–4.69) - High = 2.31 (0.98–5.46)   Employee   - Employee Lowest = 1.60 (0.74–3.47) - Low = 1.33 (0.58–3.10) - Middle High = 1.34 (0.58–3.07) - High = 1.10 (0.48–2.56)   Highest = Reference | Sex, age group, high epidemic region, health insurance premium level |
| Fisman et al.  2020  (Canada) | Cohort  Community COVID-19 cases  January 23 2020 – May 15 2020 | Mortality | Case fatality 8% | Socioeconomic determinants = OR (CI 95%)  Low income = 1.04 (0.92–1.18)  Non-health care worker = 30.56 (15.77–59.22)  Non-homeless shelter  Worker = 5.79 (0.80–41.96)  Non-homeless = 2.31 (0.94–5.71) | NA | NA |
| Guo et al. 2020    (China) | N = 14 238  Older persons with COVID-19 in Wuhan  December 12 2019 – March 17 2020 | Mortality | N = 2 758 (19.37%)  Downtown N = 2366 (85.79%)  Suburb N = 392 (14.21%) | Socioeconomic determinants = Fatality ratio  Downtown = 20.38  Suburb = 14.92 | NA | NA |
| Ibarra-Nava et al. 2021  (Mexico) | N = 416 546  Adult patients with COVID-19  February 28 2020 – August 3 2020 | Mortality | 46 508 deaths (11.25%)   - Non-Indigenous N = 45 817 (98.51%) - Indigenous N = 691 (1.49%) | NA | Socioeconomic determinants = OR (CI 95%)  Ethnic group   - Non-Indigenous = Reference - Indigenous = 1.13 (1.03–1.24) | Age, sex, sector (private, public), comorbidities (diabetes, COPD, high blood pressure, chronic kidney disease), risk factors (obesity, smoking) |
|  | N = 113 853  Hospitalised patients with COVID-19  February 28 2020 – August 3 2020 | Mortality | 41 287 deaths (11.25%)   - Non-Indigenous N = 40 692 (98.51%) - Indigenous N = 595 (1.44 %) | NA | Socioeconomic determinants = OR (CI 95%)  Ethnic group   - Non-Indigenous = Reference - Indigenous = 0.92 (0.83–1.02) | Age, sex, sector (private, public), comorbidities (diabetes, COPD, high blood pressure, chronic kidney disease), risk factors (obesity, smoking) |
| Mak et al. 2021  (UK) | N = 2 812  COVID-19 in-patients  March 1 2020 - November 30 2020 | Mortality | 417 COVID-19 deaths (14.83%) | NA | Socioeconomic determinants = OR (CI 95%)  Ethnicity   - White = Reference - Asian = 1.12 (0.53–2.37) - Black = 4.35 (2.28–8.29) - Others = 0.68 (0.20–1.93)   Education   - High = Reference - Intermediate = 0.96 (0.68–1.36) - Low = 0.93 (0.63–1.38)   Income   - >£52,000 = Reference - £31,000–51,999 = 1.12 (0.68–1.85) - 18,000–30,999= 1.05 (0.64–1.72) - < £18,000 = 1.19 (0.72–1.96)   Townsend deprivation quintile:   - 1 ( least deprived) = Reference - 2 = 0.89 (0.57–1.40) - 3 = 0.71 (0.45–1.11) - 4 = 0.85 (0.55–1.31) - 5 = 1.10 (0.73–1.67) | Frailty (concurrent HFRS), comorbidity (concurrent CCI), age , sex, smoking status, education, income and deprivation |
| Navaratnam et al. 2021  (UK) | N = 79 124  Hospitalized COVID-19 patients  March 1 2020 – May 31 2020 | Mortality | 28 200 in-hospital deaths (30.8%)   - White N = 21 351 (33%) - Asian N = 1802 (25·3%) - Black N = 746 1266 (25·4%) - Mixed N = 165 (22·1%) - Other ethnic groups N = 689 (20.9%) | NA | Socioeconomic determinants = OR (CI 95%)  Deprivation score = 1·002 (1·001–1·003)  Ethnicity   - White = Reference - Asian = 1.211 (1·128-1·299) - Black or Black British = 1·015 (0·935–1·103) - Mixed = 1·317 (1·080–1·605) - Other ethnic groups = 0·989 (0·893–1·096) | Age, sex, ethnicity,, frailty, comorbidities, obesity, deprivation score,temporal trends |
| Ortiz-Prado et al. 2021  (Ecuador) | N = 9 468  Country-wide COVID-19 cases  Study started in 2020 until April 18 2020 | Mortality | 474 COVID-19 deaths (5%)  Occupation   - Politicians N = 0 - Registered Nurses N = 1 - Health Care professionals no physicians N = 4 - Medical doctors N = 14 - Dentists N = 1 - Civil Services N = 16 - Others N = 26 - Blue Collar N = 186 - Unemployed N = 53 - Retired N = 39 - Prisoners N = 1 - White collar jobs N = 18 | NA | Socioeconomic determinants = RR (CI 95%)  Occupation   - White collar jobs = Reference - Politicians =0.016(0.001-0.273) - Registered Nurses = 0.1(0.013-0.732) - Health Care professionals no physicians = 0.211(0.074-0.6) - Medical doctors = 0.487(0.257-0.923) - Dentists = 0.939(0.131-6.705) - Civil Services = 1.458(0.796-2.672) - Others = 1.87(1.1-3.164) - Blue Collar = 3.658(2.465-5.433) - Unemployed = 4.476(2.863-6.999) - Retired = 5.649(3.541-9.013) - Prisoners = 7.75(1.363-44.04) | Age, sex, comorbidities |
| Mehta et al. 2021  (USA) | N = 137 119  Nursing home residents  April 1, 2020 - September 30, 2020 | Hospitalisation for COVID-19 | 29 204 COVID-19 hospitalisations (21.30%)  • Black N = 7 293 (25%)  • Asian N = 1 067 (4%)  • Hispanic or Latino N= 2 969 (10%)  • Other N = 148 (1%)  • White = 17 727 (61%) | NA | Socioeconomic determinants = HR (95% CI)   - Ethnicity - White = Reference - Black = 1.28 (1.24-1.32) - Asian = 1.46 (1.36-1.57) - Hispanic or Latino = 1.20 (1.15-1.26) - Other = 1.09 (0.92-1.31) | Age, BMI, sex, comorbidities |
|  |  | Survival | 26 384 COVID-19 hospitalisations (19.24%)  • Black N = 4 690 (18%)  • Asian N = 853 (3%)  • Hispanic or Latino N= 2 133 (8%)  • Other N = 160 (1%)  • White N = 18 548 (70%) | NA | Socioeconomic determinants = HR (CI 95%)  Ethnicity   - White = Reference - Black = 0.99(0.95-1.02) - Asian = 1.19(1.1-1.28) - Hispanic or Latino = 0.97 (0.93-1.02) - Other = 1.35(1.15-1.59) | Age, BMI, sex, comorbidities |
| Poulson et al. 2020  (USA) | N = 124 780  COVID-19 cases  April 5 2020 until May 18 2020 | Hospitalisation for COVID-19 | 38 199 Hospitalisations (16.19%)   - Non-Hispanic White N = 19 990 (26.2%)   Black N = 18 209 (37.7%) | Socioeconomic determinants = RR (95% CI)  Ethnicity   - Non-Hispanic White = Reference - Black = 1.53(1.5-1.55) | Socioeconomic determinants = RR (95% CI)  Ethnicity   - Non-Hispanic White = Reference - Black = 1.42(1.4-1.44) | Sex, age group, and comorbidities |
|  |  | ICU admission | 5 996 ICU admissions (4.8%)   - Non-Hispanic White N = 3 225 (4.2%)   Black N = 2 771 (5.7%) | Socioeconomic determinants = RR (95% CI)  Ethnicity   - Non-Hispanic White = Reference - Black = 1.81(1.73-1.89) | Socioeconomic determinants = RR (95% CI)  Ethnicity   - Non-Hispanic White = Reference - Black = 1.68(1.6-1.77) | Sex, age group, and comorbidities |
|  |  | Mechanical ventilation | 4 330 Mechanical ventilations (3.5%)   - Non-Hispanic White N = 2 351 (3.1%)   Black N = 1 979 (4.1%) | Socioeconomic determinants = RR (95% CI)  Ethnicity   - Non-Hispanic White = Reference   Black = 1.81(1.71-1.91) | Socioeconomic determinants = RR (95% CI)  Ethnicity   - Non-Hispanic White = Reference - Black = 1.81(1.71-1.91) | Sex, age group, and comorbidities |
|  |  | Mortality | 15 423 deaths (12.36%)   - Non-Hispanic White N = 9 141 (12%) - Black N = 6 282 (13%) | Socioeconomic determinants = RR (CI 95%)  Ethnicity   - Non-Hispanic White = Reference   Black = 1.29(1.25-1.33) | Socioeconomic determinants = RR (CI 95%)  Ethnicity   - Non-Hispanic White = Reference - Black = 1.36(1.32-1.39) | Sex, age group, and comorbidities |
| Zelner 2021  (USA) | N = 49 701  COVID-19 cases in the general population  March 8 2020 until July 5 2020 | Mortality | 5 815 deaths (11.7%)   - Black N = 2 430 (41.79%) - Latino N = 133 (2.29%) - Other N = 104 (1.78%) - Asian/Pacific Islander N = 76 (1.3%) - White N = 3 064 (52.7%) - Native American N = 8 (0.14%) | NA | Socioeconomic determinants = MRR (CI 95%)  Race   - Black = 6.7 (6.4- 7.1) - Latino = 1.9 (1.6- 2.3) - Other = 3.4 (2.7- 4.1) - Asian/Pacific = 1.4 (1.1- 1.8) - White = Reference - Native American = 0.8 (0.3- 1.5) | Sex and age |
| Cifuentes et al. 2021  (Columbia) | N = 1 033 218  Nationwide confirmed cases of COVID-19  March 2 2020 – October 26 2020 | Mortality | 30565 deaths (2.96%)  Ethnicity   - White/ Mestizo N = 28 366 (2.92%) - African Columbian N = 1421 (3.62%) - Indigenous N = 22787 776 (3.41%) - Gipsy – Roman N= 2 (5.56%) - Raizal N = 0 (0.00%)   Area of residence   - Urban N = 27 844 (3.13%) - Semirural (village) N = 1296 (2.82%) - Sparse rural N = 991 (3.81%) - Unknown area N = 434 (0.60%)   Type of health insurance regime   - Contributory regime N = 13 415 (2.03%) - Subsidised N = 11 250 (7.14%) - Special N = 463 (2.76%) - Exception N = 1247 (2.56%) - Uninsured N = 474 (2.51%) - Unknown or pending insurance N = 10384 269 (2.59%) - Non-registered insurance N = 3447 (2.85%)   Socioeconomic strata   - Very low N = 8 644 (4.49%) - Low N = 12 113 (3.20%) - Middle low N = 5 276 (2.57%) - Middle N = 918 (2.53%) - Middle high N = 290 (2.39%) - High N = 207 (3.27%) - Unkown N = 3117 (1.54%) | NA | Socioeconomic determinants = OR (95% CI)  Ethnicity   - White/ Mestizo = Reference - African Columbian = 1.01 (0.96 to 1.08) - Indigenous = 1.27 (1.13 to 1.43) - Gipsy - Roman = 1.56 (0.39 to 6.25) - Raizal = 0.00 (0.00 to 3.44)   Area of residence   - Urban = Reference - Semirural (village) = 0.88 (0.82 to 0.93) - Sparse rural = 0.83 (0.76 to 0.91) - Unknown area = 0.14 (0.12 to 0.16)   Type of health insurance regime   - Contributory regime = Reference - Subsidised = 1.97 (1.89 to 2.04) - Special = 1.29 (1.17 to 1.41) - Exception = 1.37 (1.29 to 1.45) - Uninsured = 1.34 (1.21 to 1.48) - Unknown or pending insurance = 1.22 (1.07 to 1.39) - Non-registered insurance = 2.57 (2.41 to 2.73)   Socioeconomic strata   - Very low N = 1.73 (1.48 to 2.04) - Low = 1.61 (1.38 to 1.87) - Middle low = 1.34 (1.16 to 1.56) - Middle = 1.16 (0.99 to 1.36) - Middle high = 0.94 (0.79 to 1.13) - High = Reference   Unkown = 1.54 (1.30 to 1.83) | Age, sex, ethnicity, type of health insurance, area of residence and socioeconomic strata |
| Santos et al.  2021  (Brazil) | N = 12 199  Hospitalised older people due to COVID-19  February 28 2020 – May 18 2020 | Survival | 7 863 deaths (60.53%)  Level of education   - Higher education N = 246 (10.8%) - Intermediate / Elementary education N = 521 (22.8%) - Illiterate or low education N = 1515 (66.4%)   Race   - White or Asian N = 2 542 (50.5%) - Black or mixed N =2 490 (49;5%)   Zone   - Urban N = 6,754 (97.8%) - Rural N = 152 (2.2%) | Socioeconomic determinants = HR (95% CI)  Level of education   - Higher education = Reference - Intermediate / Elementary education = 1.66 (1.43-1.94) - Illiterate or low education = 2.05(1.79-2.35)   Race   - White or Asian = Reference - Black or mixed = 1.59 (1.51-1.69)   Zone   - Urban = Reference - Rural = 1;29 (1.10-1.52) | Socioeconomic determinants = HR (95% CI)  Level of education   - Higher education = Reference - Intermediate / Elementary education = 1.35 (1.13-1.62) - Illiterate or low education = 1.60 (1.35-1.89)   Race   - White or Asian = Reference - Black or mixed = 1.18 (1.05-1.33) | Age, region, hospitalisation date, race, time elapsed between first symptom to hospitalisation, municipality that provided assistance, ICU, level of education |
| Santos et al. 2020  (Brazil) | N = 46 285  Patients admitted to Brazilian hospitals due to COVID-19  February 20 2020 – June 2 2020 | Survival | 21 408 deaths (46.25%)  Level of education   - Higher education N = 704 (11.2%) - Intermediate / Elementary education N = 1 775 (28.1%) - Low education N = 1515 (66.4%) - Illiterate N = 633 (10%)   Race   - White or Asian 5 930 (42.9%) - Black or mixed N =7 893 (57.1%)   Zone   - Urban N = 18 315 (97.3%) - Rural N = 503 (2.7%) | Socioeconomic determinants = HR (95% CI)  Level of education   - Higher education = Reference - Intermediate / Elementary education = 1.49 (1.36-1.62) - Low education = 2.23(2.06-2.42) - Illiterate = 3;29 (2.95-3.66)   Race   - White or Asian = Reference - Black or mixed = 1.54 (1.49-1.59)   Zone   - Urban = Reference - Rural = 1.26 (1.16-1.38) | Socioeconomic determinants = HR (95% CI)  Race   - White or Asian = Reference - Black or mixed = 1.5 (1.43-1.58) | Age, level of education, race, urban/rural, Influenza-like outbreak, Hospital-acquired infection, Fever, Cough, Throat, Dyspnoea, Respiratory distress, O2 <95% saturation, Diarrhoea, Vomit, Cardiopathy, Down’s syndrome, Hepatic, Asthma, Diabetes, Neurological, Pneumopathy, Immunodepression, Kidney disease, Anti-viral, ICU, Ventilatory support, X-ray, ICU length of stay |
| **Ecological studies** | | | | | | |
| Lewis et al. 2020  (USA) | 99 Utah small statistical areas  General population COVID-19 cases  July 3 2020 – July 9 2020 | Hospitalisation for COVID-19 | 1 781 hospitalisations   - Very low N = 160 (8.98%) - Low N = 324 (18.19%) - Average N = 346 (19.43%) - High N = 375 (21.06%) - Very high N = 576 (32.34%) | Socioeconomic determinants = OR (95% CI)  Level of deprivation   - Very low = Reference - Low = 1.14 (0.94–1.38) - Average = 1.39 (1.15–1.69) - High = 1.16 (0.96–1.41) - Very high = 1.40 (1.17–1.68) | Socioeconomic determinants = OR (95% CI)  Level of deprivation   - Very low = Reference - Low = 1.22 (1–1.47) - Average = 1.52 (1.26–1.84) - High = 1.37 (1.14–2.65) - Very high = 1.64 (1.38–1.97) | Age |
| Loomba et al. 2020  (USA) | N = 205 CPAs  COVID-19 positive admissions  March 14, 2020 - April 14, 2020 | ICU admission | 2.8 COVID-19 positive admissions (CPAs) per million children | NA | Socioeconomic determinants = Beta coefficient (p-value)  Percent households below poverty line = 0.053 (0.612)  Percent households with below high school as the highest adult education level = 0.111 (0.502)  Percent pediatric who have health insurance = 0.24 (0.294)  Percent pediatric who have a medical home = -0.098 (0.449)  Percent Hispanic = 0.015 (0.725)  Percent White = -0.038 (0.256)  Percen Black = -0.093 (0.072)  Percent other race = -0.021 (0.638)  Population density = 0;008 (0.004) | Age, gender, comorbidities |
| Akanbi et al. 2020  (USA) | 70 zip codes  General population  April 3 2020 – May 16 2020 | Mortality | Mean (SD)  12.77 deaths (15.21) | Socioeconomic determinants = IRR (CI 95%)  Ethnicity = 1(1-1.01)  Education level = 1.01 (1-1.01)  Median family income = 1 (0.92-1.01)  Poverty level = 0.98 (0.91-1.04) | Socioeconomic determinants = IRR (CI 95%)  Ethnicity = 0.999 (0.99-1.01)  Education level = 1.02 (1-1.04)  Median family income = 0.77(0.54-1.09)  Poverty level = 1.01 (0.97-1.06) | Median family income, poverty level, number of persons per household, mode of transportation, age, education level, ethnicity |
| Ojinnaka et al. 2021  (USA) | 254 Texas counties  Community - County-level cases  March 4, 2020 - August 1, 2020 | Mortality |  | Socioeconomic determinants = OR (CI 95%)  Percent Black = 1.95 (1.03- 3.68)  Percent Non-Hispanic White= 1.61 (0.87- 2.99)  Percent Asian = 7.53 (1.99- 28.36)  Percent Hispanic = 1.65 (0.90- 3.02) | Socioeconomic determinants = OR (CI 95%)   - Percent Black = 0.83 (0.15- 4.49) - Percent Non-Hispanic White = 0.64 (0.10- 4.15) - Percen Asian = 0.71 (0.04- 12.45) - Percent Hispanic= 0.67 (0.11- 3.85) - Percent uninsured = 1.01 (0.61- 1.70) - Primary case physician rate= 0.94 (0.89- 1.00) - High school graduation rate = 0.84 (0.53- 1.32) - Percent unemployed = 0.97 (0.26- 3.62) - Black White segregation index = 1.00 (0.93- 1.08) - Percent rural = 0.92 (0.85- 1.00) | Health factors, healthcare access measures and other county-level demographic characteristic |
| Millan-Guerrero et al. 2021  (Mexico) | N = 231 772  All individuals diagnosed with COVID-19  February 27, 2020 - July 1, 2020 | Survival | NA | Socioeconomic determinants = HR (95% CI)  Poverty level   - No poverty = Reference - Poverty = 0.99 (0.95 – 1.04) - Extreme poverty = 1.13(1.1-1.16) | Socioeconomic determinants = HR (95% CI)  Poverty level   - No Poverty = Reference - Poverty = 0.98(0.94 – 1.03) - Extreme poverty = 1.09(1.06-1.12) | Sex, age and number of comorbidities |
