## Supplemental file 3 for "The role of socio-economic determinants in SARS-CoV-2 health outcomes: systematic review of population-based studies"

### Appendix

###### Table 1. Study quality assessment - for cohort studies (Individual-level)

| STUDY | SELECTION | | | | COMPARABILITY | OUTCOME | | | TOTAL |
| --- | --- | --- | --- | --- | --- | --- | --- | --- | --- |
|  | **Representativeness of Exposed Cohort** | **Selection of Non Exposed Cohort** | **Ascertainment of Exposure** | **Demonstration Outcome of Interest Not Present at Start of Study** | **Comparability of Cohorts on the Basis of Design or Analysis** | **Ascertainment of Outcome** | **Adequate Length of Follow Up** | **Adequacy of Follow Up** |  |
| Argoty-Panjota et al.  2021 | 1 | 1 | 0 | 1 | 0 | 1 | 1 | 1 | 6 |
| Ayoubkhani et al.  2020 | 1 | 1 | 1 | 1 | 0 | 1 | 1 | 1 | 7 |
| Azar et al.  2020 | 1 | 1 | 1 | 0 | 1 | 1 | 1 | 1 | 7 |
| Bailey et al. | 1 | 1 | 1 | 1 | 0 | 1 | 1 | 1 | 7 |
| Brandén et al.  2020 | 1 | 1 | 1 | 1 | 2 | 1 | 1 | 1 | 9 |
| Chadeau-Hyam et al.  2020 | 1 | 1 | 1 | 1 | 0 | 1 | 1 | 1 | 7 |
| Cifuentes et al.  2021 | 1 | 1 | 1 | 1 | 0 | 1 | 1 | 1 | 7 |
| Clift et al.  2020 | 1 | 1 | 1 | 1 | 0 | 1 | 1 | 1 | 7 |
| Drefahl et al.  2020 | 1 | 1 | 1 | 1 | 2 | 1 | 1 | 1 | 9 |
| Elimian et al.  2020 | 1 | 1 | 1 | 1 | 0 | 1 | 1 | 1 | 7 |
| Elliott et al.  2020 | 1 | 1 | 1 | 1 | 0 | 1 | 1 | 1 | 7 |
| Fabiani et al.  2021 | 1 | 1 | 1 | 1 | 1 | 1 | 1 | 1 | 8 |
| Ferrando-Vivas et al.  2021 | 1 | 1 | 1 | 1 | 0 | 1 | 1 | 1 | 7 |
| Fisman et al.  2020 | 1 | 1 | 1 | 1 | 0 | 1 | 1 | 1 | 7 |
| Guijarroa et al.  2021 | 1 | 1 | 1 | 1 | 0 | 1 | 1 | 1 | 7 |
| Hamadah et al. | 1 | 1 | 1 | 1 | 0 | 1 | 1 | 1 | 7 |
| Izurieta et al.  2021 | 1 | 1 | 1 | 1 | 0 | 1 | 1 | 1 | 7 |
| Kim et al.  2020 | 1 | 1 | 1 | 1 | 0 | 1 | 1 | 1 | 7 |
| Kolin et al.  2020 | 1 | 1 | 1 | 1 | 0 | 1 | 1 | 1 | 7 |
| Mak et al.  2021 | 1 | 1 | 1 | 1 | 0 | 1 | 1 | 1 | 7 |
| Mehta et al.  2021 | 1 | 1 | 1 | 1 | 0 | 1 | 1 | 1 | 7 |
| Nafilyan et al.  2021 | 1 | 1 | 1 | 1 | 1 | 1 | 1 | 1 | 8 |
| Navaratnam et al. | 1 | 1 | 1 | 1 | 0 | 1 | 1 | 1 | 7 |
| Patel et al.  2020 | 1 | 1 | 1 | 1 | 0 | 1 | 1 | 1 | 7 |
| Peres et al.  2021 | 1 | 1 | 0 | 1 | 0 | 1 | 1 | 1 | 6 |
| Raisi-Estabragh et al.  2020 | 1 | 1 | 1 | 1 | 0 | 1 | 1 | 1 | 7 |
| Rosenthal et al.  2020 | 1 | 1 | 1 | 1 | 0 | 1 | 1 | 1 | 7 |
| Rostila et al.  2021 | 1 | 1 | 1 | 1 | 0 | 1 | 1 | 1 | 7 |
| Santos et al. 2020 | 1 | 1 | 1 | 1 | 0 | 1 | 1 | 1 | 7 |
| Santos et al. 2021 | 1 | 1 | 1 | 1 | 0 | 1 | 1 | 1 | 7 |
| Telle et al.  2021 | 1 | 1 | 1 | 1 | 0 | 1 | 1 | 1 | 7 |
| Williamson et al.  2020 | 1 | 1 | 1 | 1 | 0 | 1 | 1 | 1 | 7 |
| Wollenstein-Betech et al.  2020 | 1 | 1 | 1 | 1 | 1 | 1 | 1 | 1 | 8 |
| Wu et al.  2020 | 1 | 1 | 1 | 1 | 0 | 1 | 1 | 1 | 7 |
| Ho et al.  2020 | 1 | 1 | 1 | 1 | 0 | 1 | 1 | 1 | 7 |
| Atkins et al.  2020 | 1 | 1 | 1 | 1 | 0 | 1 | 1 | 1 | 7 |
| Mutambudzi et al.  2021 | 1 | 1 | 1 | 1 | 0 | 1 | 1 | 1 | 7 |
| Mathur et al.  2021 | 1 | 1 | 1 | 1 | 0 | 1 | 1 | 1 | 7 |
| Zelner et al.  2021 | 1 | 1 | 1 | 1 | 0 | 1 | 1 | 1 | 7 |

###### Table 2. Study quality assessment - for cohort studies (Ecological studies)

| STUDY | SELECTION | | | | COMPARABILITY | OUTCOME | | | TOTAL |
| --- | --- | --- | --- | --- | --- | --- | --- | --- | --- |
|  | **Representativeness of Exposed Cohort** | **Selection of Non Exposed Cohort** | **Ascertainment of Exposure** | **Demonstration Outcome of Interest Not Present at Start of Study** | **Comparability of Cohorts on the Basis of Design or Analysis** | **Ascertainment of Outcome** | **Adequate Length of Follow Up** | **Adequacy of Follow Up** |  |
| Khanijahani et al.  2021 | 1 | 1 | 1 | 1 | 0 | 1 | 1 | 1 | 7 |
| Gaudart et al.  2021 | 1 | 1 | 1 | 1 | 0 | 1 | 1 | 1 | 7 |
| Macchia et al.  2021 | 1 | 1 | 1 | 0 | 0 | 1 | 1 | 1 | 6 |
| Loomba et al.  2020 | 1 | 1 | 1 | 0 | 0 | 1 | 1 | 1 | 6 |
| Millan-Guerrero et al.  2020 | 1 | 1 | 1 | 1 | 0 | 1 | 1 | 1 | 7 |
| Ojinnaka et al.  2021 | 1 | 1 | 1 | 0 | 0 | 1 | 1 | 1 | 6 |

###### Table 3. Study quality assessment - for cross-sectional studies (Individual-level)

| STUDY | SELECTION | | | | COMPARABILITY | OUTCOME | | TOTAL |
| --- | --- | --- | --- | --- | --- | --- | --- | --- |
|  | **Sample Representativeness** | **Sample Size** | **Non-respondents** | **Ascertainment of the Exposure (risk factor)** | **Comparability of Different Outcome Groups on the Basis of Design or Analysis** | **Assessment of Outcome** | **Statistical Test** |  |
| Baqui et al.  2020 | 1 | 1 | 1 | 2 | 0 | 2 | 1 | 8 |
| Bassett et al.  2020 | 1 | 1 | 1 | 2 | 1 | 2 | 1 | 9 |
| Aung et al.  2021 | 1 | 1 | 1 | 1 | 0 | 1 | 1 | 6 |
| Hanson et al.  2020 | 1 | 1 | 1 | 1 | 0 | 2 | 1 | 7 |
| Levin et al.  2021 | 1 | 1 | 1 | 2 | 0 | 2 | 1 | 8 |
| Guo et al.  2020 | 1 | 1 | 1 | 0 | 0 | 2 | 1 | 6 |
| Ibarra-Nava et al.  2021 | 1 | 1 | 1 | 1 | 0 | 2 | 1 | 7 |
| Mendez-Dominguez et al.  2020 | 1 | 1 | 1 | 1 | 0 | 2 | 1 | 7 |
| Ortiz-Prado et al.  2021 | 1 | 1 | 1 | 2 | 0 | 2 | 1 | 8 |
| Poulson et al. | 1 | 1 | 0 | 2 | 0 | 2 | 1 | 7 |
| Raisi-Estabragh et al.  2020 | 1 | 1 | 1 | 2 | 0 | 2 | 1 | 8 |
| Rossi et al.  2020 | 1 | 1 | 0 | 2 | 0 | 2 | 1 | 7 |

###### Table 4. Study quality assessment - for cross-sectional studies (Ecological)

| STUDY | SELECTION | | | | COMPARABILITY | OUTCOME | | TOTAL |
| --- | --- | --- | --- | --- | --- | --- | --- | --- |
|  | **Sample Representativeness** | **Sample Size** | **Non-respondents** | **Ascertainment of the Exposure (risk factor)** | **Comparability of Different Outcome Groups on the Basis of Design or Analysis** | **Assessment of Outcome** | **Statistical Test** |  |
| Adhikari et al.  2020 | 1 | 1 | 1 | 1 | 0 | 2 | 1 | 7 |
| Li et al. | 1 | 1 | 1 | 1 | 0 | 2 | 1 | 7 |
| Temkin-Greener et al.  2020 | 1 | 1 | 1 | 1 | 0 | 2 | 1 | 7 |
| Alves et al.  2021 | 1 | 1 | 1 | 2 | 0 | 2 | 1 | 8 |
| Abedi et al.  2020 | 1 | 1 | 1 | 2 | 0 | 2 | 1 | 8 |
| Biggs et al.  2020 | 1 | 1 | 1 | 2 | 0 | 2 | 1 | 8 |
| Chen et al.  2020 | 1 | 1 | 1 | 2 | 0 | 2 | 1 | 8 |
| Daras et al.  2021 | 1 | 1 | 1 | 2 | 0 | 2 | 1 | 8 |
| De Souza et al.  2020 | 1 | 1 | 1 | 1 | 0 | 2 | 1 | 7 |
| Demenech et al.  2020 | 1 | 1 | 1 | 1 | 0 | 0 | 1 | 5 |
| Di Girolamo et al.  2020 | 1 | 1 | 1 | 1 | 0 | 2 | 1 | 7 |
| Figueroa et al.  2020 | 1 | 1 | 1 | 2 | 0 | 2 | 1 | 8 |
| Figueroa et al.  2021 | 1 | 1 | 1 | 2 | 0 | 2 | 1 | 8 |
| Garcia et al.  2021 | 1 | 1 | 1 | 2 | 0 | 2 | 1 | 8 |
| Hawkins D. et al.  2020 | 1 | 1 | 1 | 2 | 0 | 2 | 1 | 8 |
| Hawkins R. et al.  2020 | 1 | 1 | 1 | 2 | 0 | 2 | 1 | 8 |
| Karmakar et al.  2021 | 1 | 1 | 1 | 1 | 0 | 2 | 1 | 7 |
| Khazanchi et al.  2020 | 1 | 1 | 1 | 2 | 0 | 2 | 1 | 8 |
| Georges et al.  2021 | 1 | 1 | 1 | 1 | 0 | 2 | 1 | 7 |
| Akanbi et al.  2020 | 1 | 1 | 1 | 1 | 0 | 2 | 1 | 7 |
| Li Y. et al.  2020 | 1 | 1 | 1 | 2 | 0 | 2 | 0 | 7 |
| Loomba et al.  2020 | 1 | 1 | 1 | 1 | 0 | 2 | 1 | 7 |
| Palacio et al.  2020 | 1 | 1 | 1 | 1 | 0 | 2 | 1 | 7 |
| Rodriguez-Villamizar et al.  2021 | 1 | 1 | 1 | 1 | 0 | 2 | 1 | 7 |
| Sugg et al.  2021 | 1 | 1 | 1 | 1 | 0 | 1 | 1 | 6 |
| Yang et al.  2021 | 1 | 1 | 1 | 1 | 0 | 2 | 1 | 7 |
| Lewis et al.  2020 | 1 | 1 | 1 | 1 | 0 | 2 | 1 | 7 |
| Das et al.  2020 | 1 | 1 | 1 | 1 | 0 | 0 | 1 | 5 |
| Li et al.  2020 | 1 | 1 | 1 | 1 | 0 | 2 | 1 | 7 |
| Alipio et al.  2020 | 1 | 1 | 1 | 0 | 0 | 0 | 1 | 4 |
| Ramirez-Aldana et al.  2020 | 1 | 1 | 1 | 2 | 0 | 0 | 1 | 6 |
| Ginsburgh et al.  2021 | 1 | 1 | 1 | 2 | 0 | 0 | 1 | 6 |
| Mollalo et al.  2020 | 1 | 1 | 1 | 2 | 0 | 2 | 1 | 8 |
| Ehlert et al.  2020 | 1 | 1 | 1 | 2 | 2 | 2 | 1 | 10 |
| Gross et al.  2020 | 1 | 1 | 1 | 2 | 0 | 2 | 0 | 7 |
| Karaye et al.  2020 | 1 | 1 | 1 | 2 | 0 | 2 | 1 | 8 |
| Khanijahani et al.  2020 | 1 | 1 | 1 | 2 | 0 | 2 | 1 | 8 |
| White et al.  2020 | 1 | 1 | 1 | 1 | 0 | 2 | 1 | 7 |
| Zhang et al.  2020 | 1 | 1 | 1 | 2 | 0 | 2 | 1 | 8 |
| Fielding-Miller et al. 2020 | 1 | 1 | 1 | 2 | 0 | 0 | 1 | 6 |

###### Table 5. Study quality assessment - for case-control studies (Individual-level)

| STUDY | SELECTION | | | | COMPARABILITY | EXPOSURE | | | TOTAL |
| --- | --- | --- | --- | --- | --- | --- | --- | --- | --- |
|  | **Case Definition** | **Representativeness of Cases** | **Selection of Controls** | **Definition of Controls** | **Comparability of Cases and Controls** | **Ascertainment of Exposure** | **Same Method Ascertainment Both Groups** | **Non Response Rate** |  |
| Bergman et al. | 0 | 1 | 1 | 1 | 0 | 1 | 1 | 1 | 6 |
| McKeique et al. | 1 | 1 | 1 | 1 | 0 | 1 | 1 | 1 | 7 |
| Shahbazi et al. | 1 | 1 | 1 | 1 | 0 | 1 | 1 | 1 | 7 |
