## Supplemental file 4 for "The role of socio-economic determinants in SARS-CoV-2 health outcomes: systematic review of population-based studies"

**Newcastle-Ottawa Scale adapted for quality assessment of cross-sectional studies for the systematic review** “Etiologic and prognostic roles of frailty, multimorbidity and socioeconomic characteristics in the development of Covid-19 related severe health outcomes in the general population: protocol for systematic literature review“

Adjusted from Herzog R, et al. Is Healthcare Workers’ Intention to Vaccinate Related to their Knowledge, Beliefs and Attitudes? A Systematic Review. *BMC Public Health* 2013 **13**:154 and Modesti PA, et al. Panethnic differences in blood pressure in Europe: a systematic review and meta-analysis.

**Selection:** (maximum 5 stars)

1) Representativeness of the sample:

a) Truly representative of the average in the target population. * (all subjects or random sampling)

b) Somewhat representative of the average in the target population. * (non-random sampling)

c) Selected group of users.

d) No description of the sampling strategy.

2) Sample size:

a) Justified and satisfactory. *

b) Not justified.

3) Non-respondents:

a) Comparability between respondents and non-respondents characteristics is established, and the response rate is satisfactory. *

b) The response rate is unsatisfactory, or the comparability between respondents and non-respondents is unsatisfactory.

c) No description of the response rate or the characteristics of the responders and the non-responders.

4) Ascertainment of the exposure (risk factor):

a) Validated measurement tool. **

b) Non-validated measurement tool, but the tool is available or described.*

c) No description of the measurement tool.

**Comparability:** (maximum 2 stars)

1) The subjects in different outcome groups are comparable, based on the study design or analysis. Confounding factors are controlled.

a) The study stratifies for age and sex **

b) The study stratifies for age *

c) The study doesn’t stratify for age

**Outcome:** (maximum 3 stars)

1) Assessment of the outcome:

a) Independent blind assessment. **

b) Record linkage. **

c) Self report. *

d) No description.

2) Statistical test:

a) The statistical test used to analyse the data is clearly described and appropriate, and the measurement of the association is presented, including confidence intervals and the probability level (p value). *

b) The statistical test is not appropriate, not described or incomplete.
